# Large-scale genomic surveillance reveals immunosuppression drives mutation dynamics in persistent SARS-CoV-2 infections

**DOI:** 10.1101/2025.02.10.25321987

**Authors:** Mark P. Khurana, Alexandros Katsiferis, Neil Scheidwasser, Mathilde Marie Brünnich Sloth, Jacob Curran-Sebastian, Christian Morgenstern, Man-Hung Eric Tang, Jannik Fonager, Morten Rasmussen, Marc Stegger, Charles Whittaker, The Danish COVID-19 Genome Consortium (DCGC), Sune Lehmann, Laust H. Mortensen, Pikka Jokelainen, Moritz U G Kraemer, Neil M Ferguson, Mahan Ghafari, Tyra G. Krause, David A. Duchêne, Samir Bhatt

**Affiliations:** Section for Health Data Science and AI, Department of Public Health, University of Copenhagen, Copenhagen, Denmark; MRC Centre for Global Infectious Disease Analysis, Department of Infectious Disease Epidemiology, School of Public Health, Faculty of Medicine, Imperial College London, London, United Kingdom; Department of Sequencing and Bioinformatics, Statens Serum Institut, Copenhagen, Denmark; Department of Virology and Microbiological Preparedness, Statens Serum Institut, Copenhagen, Denmark; Antimicrobial Resistance and Infectious Diseases Laboratory, Harry Butler Institute, Murdoch University, Murdoch, WA, Australia; Department of Applied Mathematics and Computer Science, Technical University of Denmark, Kongens Lyngby, Denmark; Statistics Denmark, Copenhagen, Denmark; Infectious Disease Preparedness and One Health, Statens Serum Institut, Copenhagen, Denmark; Department of Biology, University of Oxford, Oxford, United Kingdom; Pandemic Sciences Institute, University of Oxford, Oxford, United Kingdom; Epidemiological Infectious Disease Preparedness, Statens Serum Institut Copenhagen, Denmark

**Keywords:** persistent infection, SARS-CoV-2, viral evolution

## Abstract

Persistent SARS-CoV-2 infections have been hypothesized to play a key role in the emergence of variants of concern. However, the factors determining which individuals are at risk and their viral molecular signatures during infection remain poorly understood. Using the extensive COVID-19 surveillance database in Denmark, comprising over 700,000 genomes, we identified 303 persistent infections and, critically, linked them to health and sociodemographic data. Our analysis confirms the hypothesis that immunocompromised individuals are at the highest risk of experiencing persistent infections. Other disease groups associated with mortality, such as diabetes, showed no such associations. Among these persistent infections, the viral sequences exhibited signs of diversifying selection, with recurrent mutations linked to treatment resistance. Our findings suggest that immunosuppression plays a key role in the emergence of novelty in persistent infections.

## Introduction

Novel variants of concern are increasingly thought to arise in individuals with chronic infections, particularly those with compromised immune systems who are unable to clear an infectious episode [1–6]. In a persistent SARS-CoV-2 infection, defined as lasting 26 days or more [7], the virus is thought to avoid the tight bottlenecks upon transmission that often result in a loss of diversity after acute infections [6–8]. Chronically infected individuals are more likely to receive treatment during their infections, potentially intensifying selective pressure on the virus [6, 9, 10]. Persistent infections are also frequently found in individuals with compromised immune systems [2, 4, 6, 11], and are linked with selection for amino-acid changes that can define new virus lineages [7] and which occur at target sites for monoclonal antibodies [5, 7, 12–18]. Cryptic lineages found in wastewater samples are also likely shed by individuals with persistent infections, where unique mutational signatures have arisen [19–21]. Nonetheless, the medical diagnoses that are linked with persistent infections and the emergence of novel variants and mutations of SARS-CoV-2 remains incompletely understood [22–25].

Genomic surveillance is shifting how we detect and study pathogen spread and evolution [26, 27]. Research on within-host viral evolution is usually based on small sample sizes (e.g., case studies) [2, 3, 5], or on large cohort studies that lack individual-level data beyond sociodemographic characteristics [7, 18], such as patient history and health condition. When aiming to prevent the emergence of new variants via mitigation strategies at their source, it is critical to describe the scenarios of emergence and corresponding patient risk factors [7]. Extensive genomic surveillance coupled with detailed centralized registry data can provide unique insights into the settings linked with emergence of variants of concern.

Here, we leverage Denmark’s extensive genomic surveillance database, coupling it with national registry data to identify persistent SARS-CoV-2 infections and individual-level sociodemographic and diagnoses. Using consensus-level sequence data, we identify individuals with SARS-CoV-2 infection episodes spanning 26 days or more. We identify risk factors associated with persistent infections using a case-control study design. Further, we describe the mutational patterns associated with these infections, and identify characteristics and health conditions linked with accelerated selective genetic changes in the virus. By linking diagnostic health information directly with pathogen evolution across population-level data, our analyses assert new value to centralized health registries and epidemiological genomic surveillance efforts.

## Results

### Identifying persistent infections

To identify persistent SARS-CoV-2 infections, we analyzed all high-quality viral sequences collected by the Danish COVID-19 Genome Consortium (DCGC) from 2020 to 2024 (*n* = 738 944) (Supplementary Fig. S1). This dataset represents one of the largest and most representative sampling efforts conducted globally to date [28]. Only consensus sequences from the Alpha, Delta, and Omicron variants (BA.1, BA.2, and BA.5) were included due to a relative paucity of samples for the “wild-type” variant. For each individual, sequence pairs within the same variant were identified if sampled at least 26 days apart. Infection episodes were defined based on consensus sequences preceding and following each pair (see Methods). Consensus sequences were derived from reverse transcription (RT–PCR)-positive samples, using low cycle threshold (Ct) values as indicators of sequencing quality and viral load. Sequences with Ct ≤ 38 were retained, following DCGC guidelines for high-quality sequences included in the national sequencing database. This process yielded 455 potential persistent infections.

We then filtered the resulting subset of persistent infections to retain only those where consensus sequences shared the same rare single-nucleotide polymorphisms (SNPs) at one or more sites, relative to the population-level consensus of the major lineage, as previously described by Ghafari et al. [7]. Using our rare SNP cut-off, the false positive rate (i.e., the proportion of incorrectly identified persistent infections among randomly paired sequences) was estimated to be between 2.1% and 3.3%, resulting in 303 persistent infections (42 Alpha, 63 Delta, 27 BA.1, 132 BA.2, 39 BA.5) (Fig. 1, Supplementary Table S1). 20 infections lasted > 100 days, and five > 150 days, consistent with previously identified upper bounds for some persistent infections [6, 7]. Ct values, inversely proportional to sequencing quality and viral load, were higher at the final sequenced time point compared to the initial one (mean change in Ct = −5.28 [95% credible interval (CrI): −6.64, −3.94]) (Fig. 1f).

**Figure 1.**
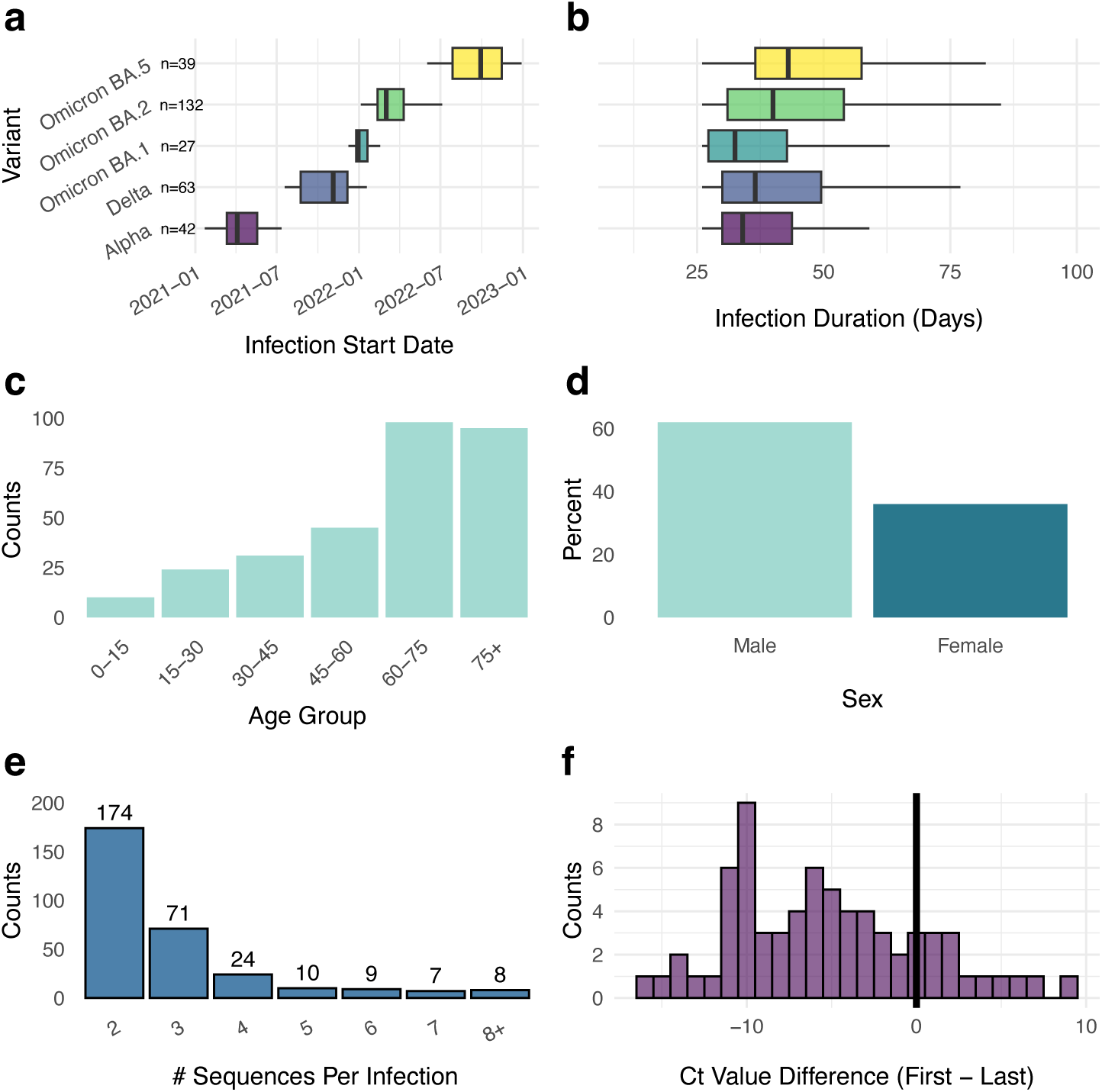
Characteristics of the study population (*n* = 303). a) Date of first sequence for each infection stratified by variant. b) Time since first and last sequence for each infected individual. c) Number of individuals in each age group. d) Number of individuals of each sex. e) Number of available sequences per infection. f) Difference in Ct values between first and last sequences for individuals with two or more available Ct values (*n* = 71). Values below zero indicate lower viral load (higher Ct) at last sequencing than at first.

### Risk factors for persistent infections

To identify risk factors for persistent SARS-CoV-2 infections, we conducted a matched case-control study. Each case of persistent infection was matched with controls who met the following criteria: a positive PCR test within ±1 day of the case’s infection start date and the same major variant, a negative test within 26 days of that positive result, and no additional positive tests within the subsequent 120 days. We identified a total of 146,904 controls across all strata, corresponding to 62,648 unique individuals. This corresponds to a median of 401 (interquartile range, IQR 162-522) controls per case. However, for 9 cases, we found fewer than 10 matches, resulting in a range of 0 to 8 matches for these cases. Using Bayesian uni- and multi-variable conditional logistic regression models [29], we assessed associations between persistent infection and key factors, including age, sex, vaccination status, Charlson Comorbidity Index (CCI) [30], and specific medical conditions (using ICD-10 codes): immunosuppression, autoimmune disorders, immune-mediated inflammatory diseases (IMID), and diabetes. To avoid temporal associations with vaccination, we only included pre-pandemic diagnoses (prior to 2020), but explored including contemporary diagnoses in supplementary analyses, finding similar results.

Compared to individuals aged 15-30, those in the 45-60, 60-75, and 75+ age groups had significantly higher odds of experiencing persistent SARS-CoV-2 infections (adjusted odds ratio, aOR 2.02 [CrI 1.28, 3.19], 5.90 [CrI 3.87, 9.11], and 7.63 [CrI 4.84, 12.01], respectively), while no significant differences were observed for the 0-15 and 30-45 age groups. Additionally, females were significantly less likely to experience persistent infection compared to males (aOR 0.57 [CrI 0.44, 0.70]) (Table 1). Compared to unvaccinated individuals, those with 2 or 3 vaccinations less than 18 weeks post-vaccination had a reduced odds of persistent infection by 46% (aOR 0.54 [CrI 0.35, 0.84]). However, individuals with one vaccination or 2–3 vaccinations ≥ 18 weeks post-vaccination showed no protection. Using a 10-year CCI lookback period resulted in similar estimates (Supplementary Table S2), as well as using contemporary 5-year CCI scores (Supplementary Table S4).

**Table 1.** Risk factors for persistent infection. Values are presented as median [IQR] for continuous variables and n (%) for categorical variables. Unadjusted and adjusted odds ratios (OR) with 95% credible intervals (CrI) are presented from the conditional logistic regression output, with adjustments for all other variables. CrI = credible interval; IQR = interquartile range. The Charlson Comorbidity Index was calculated with a 5-year lookback from January 1, 2020. Vaccination status combines the number of vaccinations at infection onset and the time since the last vaccination (*<* 18 weeks or *≥* 18 weeks) for those with 2 or 3 vaccinations. The total number of controls is *n* = 146,904, corresponding to *n* = 62,648 unique individuals.

| Variable |  | Cases (Median [IQR]) | Controls (Median [IQR]) | Unadjusted OR (95% CrI) | Adjusted* OR (95% CrI) |
| --- | --- | --- | --- | --- | --- |
| Age Group | 0-15 | 24 | 32667 | 0.61 (0.31-1.16) | 0.53 (0.26-0.99) |
|  | 15-30 | 10 | 24666 | Ref. | Ref. |
|  | 30-45 | 31 | 31606 | 1.21 (0.74-1.93) | 1.28 (0.78-2.06) |
|  | 45-60 | 46 | 29477 | 1.96 (1.27-3.08) | 2.02 (1.28-3.19) |
|  | 60-75 | 98 | 17615 | 6.88 (4.65-10.59) | 5.90 (3.87-9.11) |
|  | 75+ | 94 | 10873 | 10.47 (6.92-16.47) | 7.63 (4.84-12.01) |
| Sex | Male | 194 | 68774 | Ref. | Ref. |
|  | Female | 109 | 78130 | 0.48 (0.38-0.62) | 0.57 (0.44-0.70) |
| Vaccination status | No vaccination | 76 | 56497 | Ref. | Ref. |
|  | 1 Vaccination | 8 | 6520 | 0.94 (0.43-1.83) | 0.65 (0.30-1.27) |
|  | 2 or 3 Vaccinations, <18 weeks | 90 | 52046 | 1.27 (0.86-1.91) | 0.54 (0.35-0.84) |
|  | 2 or 3 Vaccinations, ≥ 18 weeks | 129 | 31841 | 2.17 (1.45-3.29) | 0.80 (0.53-1.24) |
| Charlson Comorbidity Index† | Per 1 Increase | 0 [0-2] | 0 [0-0] | 1.55 (1.47-1.64) | 1.35 (1.26-1.43) |

\*Adjusted for all other variables; CrI = Credible Interval; † 5-year lookback from 2020-01-01

Among individuals with specific medical conditions, immunosuppressed individuals had significantly higher odds of persistent infection (aOR 5.84 [CrI 4.74, 7.19]), consistent with findings from prior individual-level studies on chronic infections [1–6]. Those with autoimmune disorders had a marginally elevated risk of persistent infection (aOR 1.59 [CrI 1.02, 2.33]) (Table 2), although we found no evidence of increased risk in a multivariate model when adjusting for all other diagnoses (Supplementary Table S5). No increased risk was observed among individuals with diabetes or immune-mediated inflammatory diseases (aOR 1.10 [CrI 0.83, 1.42] and 1.13 [CrI 0.79, 1.55], respectively) (Table 2). We found no significant interactions between diagnoses and age or sex. We repeated the analysis using contemporary diagnoses (i.e., those registered up to 2021-12-31, for which diagnostic registry data is available), yielding similar results (Supplementary Table S3).

**Table 2.** Diagnoses associated with persistent infection. Values are presented as n for categorical variables. Unadjusted and adjusted odds ratios (OR) with 95% credible intervals (CrI) are presented from the conditional logistic regression output, adjusted for sex, age, and vaccination status. CrI = credible interval; IMID = immune-mediated inflammatory diseases. Diagnoses are based on pre-pandemic conditions (i.e., those prior to January 1, 2020). Vaccination status combines the number of vaccinations at infection onset and the time since the last vaccination (*<* 18 weeks or *≥* 18 weeks) for those with 2 or 3 vaccinations. The total number of controls is *n* = 146,904, corresponding to *n* = 62,648 unique individuals.

| Diagnosis Group† |  | Cases | Controls | Unadjusted OR (95% CrI) | Adjusted* OR (95% CrI) |
| --- | --- | --- | --- | --- | --- |
| Model 1: Immunosuppression | Yes | 76 | 1929 | 7.91 (6.47-9.73) | 5.84 (4.74-7.19) |
|  | No | 227 | 144975 | Ref. | Ref. |
| Model 2: Autoimmune | Yes | 13 | 1611 | 2.16 (1.38-3.24) | 1.59 (1.02-2.33) |
|  | No | 290 | 145293 | Ref. | Ref. |
| Model 3: Diabetes | Yes | 28 | 4717 | 1.81 (1.35-2.37) | 1.10 (0.83-1.42) |
|  | No | 275 | 142187 | Ref. | Ref. |
| Model 4: IMID | Yes | 18 | 4775 | 1.38 (0.98-1.89) | 1.13 (0.79-1.55) |
|  | No | 285 | 142129 | Ref. | Ref. |

\*Each model is also adjusted for sex, age, and vaccination status; CrI = Credible Interval.
† Using pre-pandemic diagnoses (i.e., those prior to 2020-01-01).

### Mutational landscape and frequent amino-acid changes

The majority of the within-host mutations observed between consecutive consensus sequences in each persistent infection occurred in the ORF1ab (open reading frame 1ab) and S (spike) coding regions of the SARS-CoV-2 genome (Fig. 2a). The highest mutation rates per site were found in the ORF7a, S, N, and E coding regions (Fig. 2c). A total of 148 infections (48.8%) showed no consensus-level mutations between any pairs of consecutive sequences in coding regions during their infections. Additionally, 175 infections (57.8%) showed no change between their first and last sequences in coding regions, and 143 infections (47.2%) showed no change when including non-coding regions. For those with at least one consensus-level change, the mean genome-wide rate was 1.03 × 10^−3^ substitutions per site per year (95% Confidence Interval, CI 2.50×10^−4^ to 2.78×10^−3^). Across all infections and pairs of consecutive sequences, we observed 679 non-synonymous (amino acid-changing) and 209 synonymous (silent) mutations in coding regions (Supplementary Fig. S2); comparing only the first and last sequences in each infection revealed 551 non-synonymous and 180 synonymous mutations (Fig. 2b).

**Figure 2.**
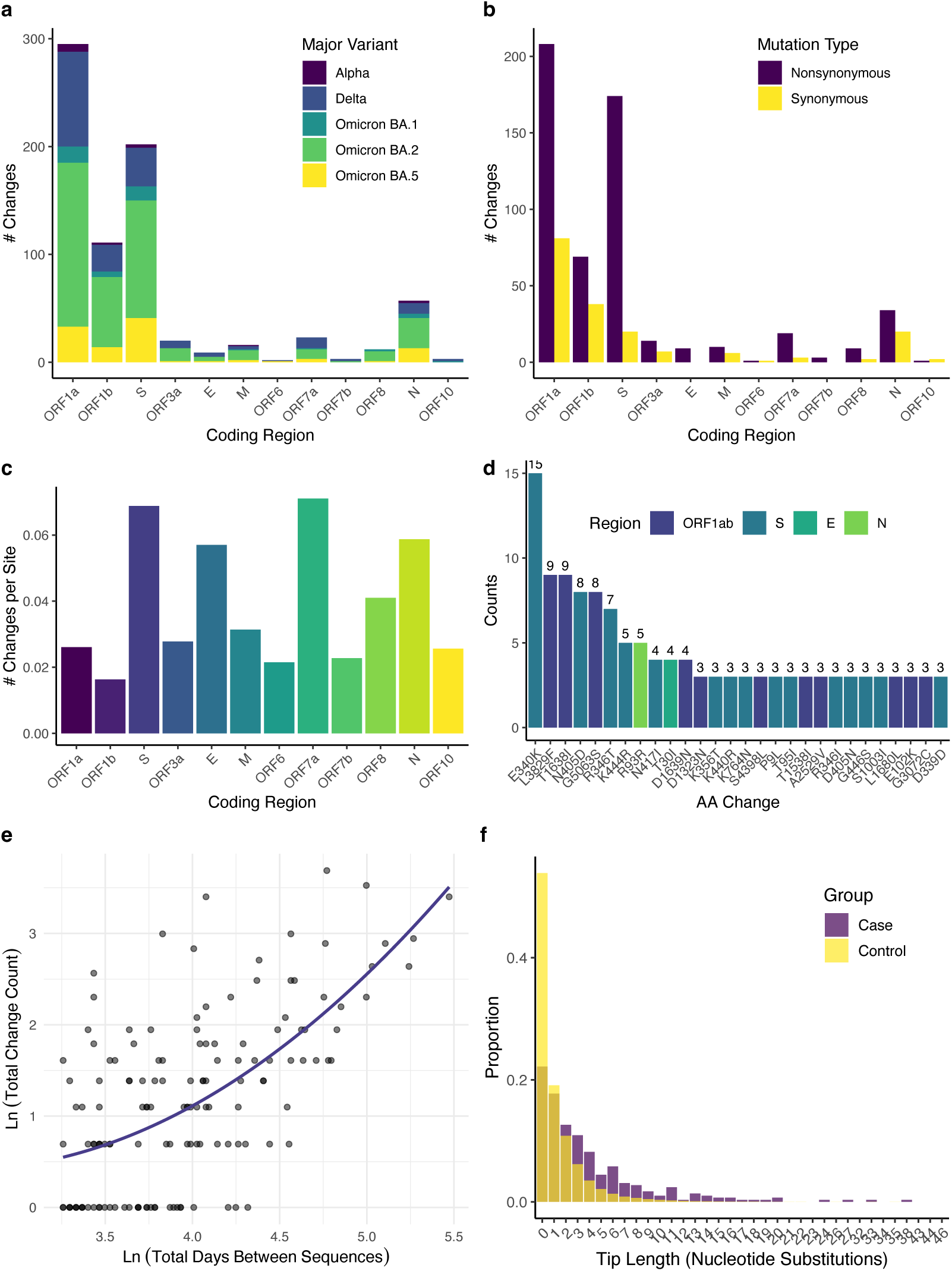
Comparison of coding region substitutions and evolutionary dynamics in persistent infections. a) Location of substitutions in each coding region. b) Location of (non)synonymous substitutions in each coding region. c) Number of nucleotide substitutions in each coding region normalized per site. d) Location of recurrent amino acid substitutions. e) Scatter plot illustrating the relationship between log-transformed total days between the first and last sequences and log-transformed total change count (including non-coding regions), excluding points without changes (*n* = 143). The Pearson correlation coefficient is 0.612. Two Bayesian models were fitted using the *brms* [33] package: a linear-log model and a quadratic-log model, with the line representing predictions from the quadratic model. Model comparison via Pareto Smoothed Importance Sampling Leave-One-Out-cross-validation (PSIS-LOOCV) showed that the quadratic model was a better fit to the data than the linear model (expected log pointwise predictive density (ELPD) difference: *−*0.8). f) Histogram comparing tip lengths of persistent infections to acute controls. Independent phylogenetic trees were constructed for each case-control set (from conditional logistic regression) with at least 10 controls (n=293).

While we did not observe evidence of genome-wide positive selection in persistent infections (Ka and Ks difference, *β* = −3.23 × 10^−3^, CrI: −1.99 × 10^−2^ to 1.47 × 10^−2^) (Fig. 3a), we found strong evidence of selection in the S coding region (*β* = 4.15 × 10^−4^, CrI: 2.70 × 10^−4^ to 5.64 × 10^−4^) (Fig. 3b). We also found evidence of positive selection in the ORF7a (*β* = 3.59 × 10^−3^, CrI: 3.98 × 10^−4^ to 6.65 × 10^−3^) and E (*β* = 6.59 × 10^−3^, CrI: 5.94 × 10^−3^ to 7.25 × 10^−3^) coding regions (Supplementary Fig. S4), although few observations were available. We found no evidence of positive or negative selection in other coding regions (Supplementary Table S14). In controls with multiple available sequences, we observed evidence of negative (purifying) selection in ORF1a (*β* = −1.59 × 10^−4^, CrI: −2.32 × 10^−4^ to −8.75 × 10^−5^) and ORF1b (*β* = −4.32 × 10^−4^, CrI: −5.64 × 10^−4^ to −3.03 × 10^−4^), with no evidence of genome-wide selection (Fig. 3a; Supplementary Table S15).

**Figure 3.**
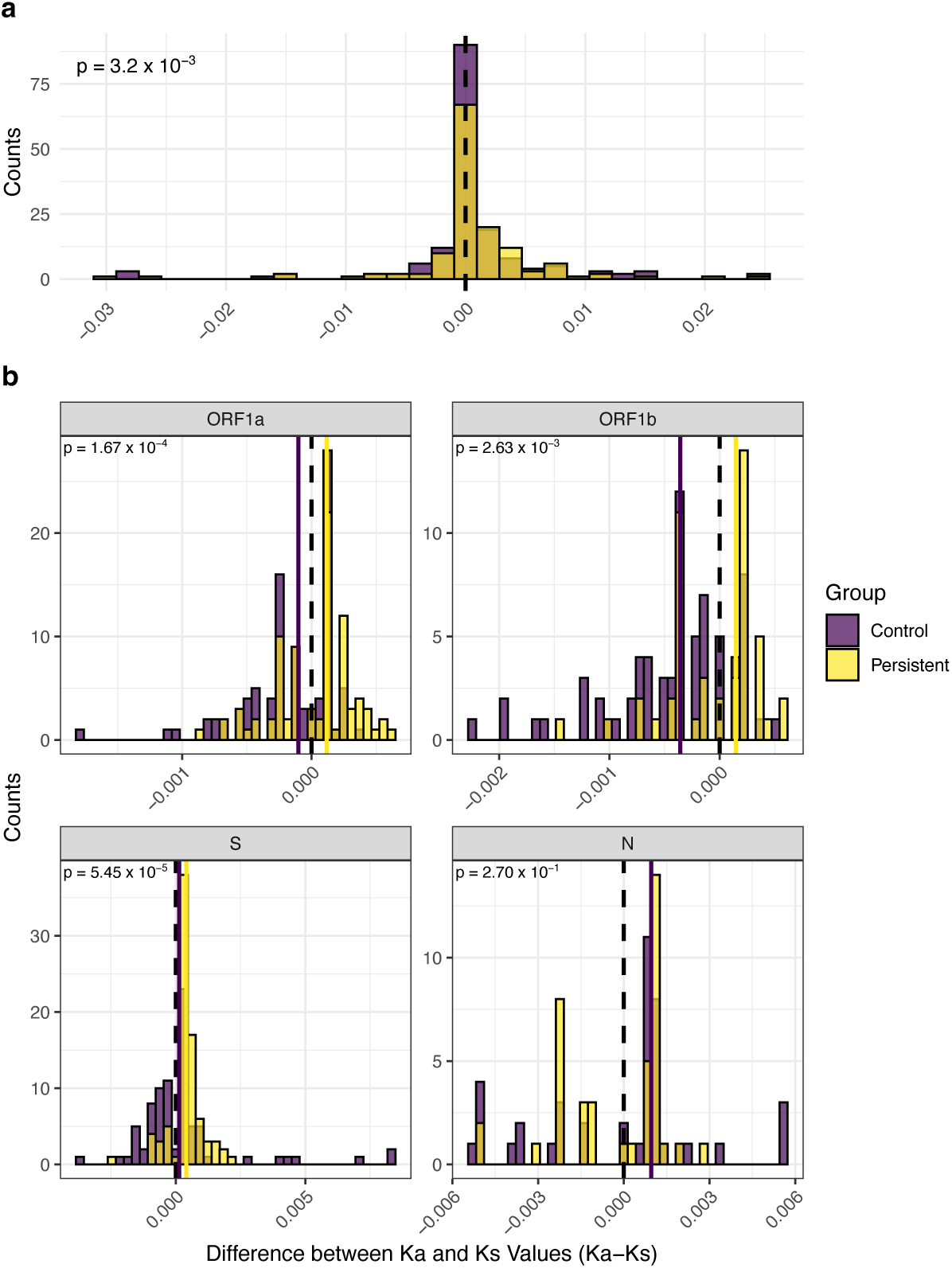
Differences in Ka and Ks values (Ka - Ks) across the genome and by coding region. (a) Difference in Ka and Ks values among individuals with persistent infections (*n* = 135) and among individuals without persistent infections (controls, *n* = 167) with two or more available sequences. Ka and Ks represent the substitution rates per non-synonymous and synonymous site, respectively. The dotted line indicates a value of 0, and the solid lines represent median values. The p-value denotes the p-value from the Kolmogorov-Smirnov (KS) test between the two distributions. (b) Difference in Ka and Ks values in each coding region among individuals with and without persistent infections, specifically those with at least one mutation in the region. P-values are from KS tests as above.

When comparing recurrent and single-appearing mutations in persistent infections, recurrent mutations exhibited significantly higher transmission fitness values than single-appearing mutations (Fig. 4). These fitness values measure the potential of mutations to spread between hosts (i.e., transmission-associated fitness) based on the ratio of observed to expected occurrences in a global SARS-CoV-2 phylogeny by Bloom et al. [31]. The approach assumes that non-deleterious mutations arise independently across the phylogenetic tree, with deviations from expectations reflecting their effects on transmission; higher values indicate mutations more likely to spread. We found that single-appearing mutations were associated with a mean fitness decrease of −0.40 (CrI −0.62, −0.18) relative to recurrent mutations, while other amino acids at matched sites showed a mean fitness decrease of −1.50 (CrI −1.86, −1.12) relative to recurrent mutations.

**Figure 4.**
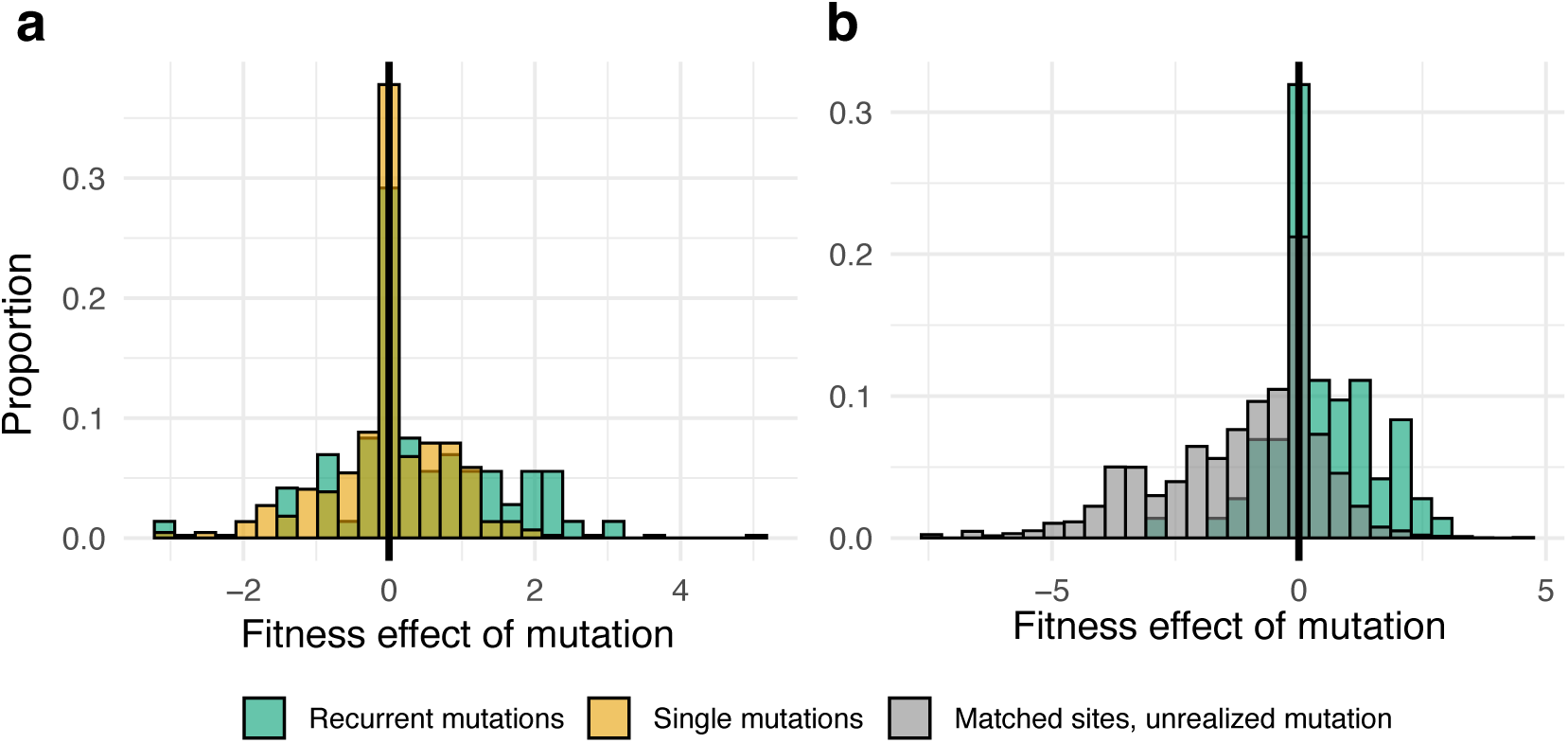
Distribution of fitness effects for recurrent and non-recurrent mutations. (a) Comparison of recurrent mutations versus single-appearing mutations in persistently infected individuals. (b) Comparison of recurrent mutations with other amino acids found at the same sites. Fitness values represent the potential for these mutations to spread between hosts, as determined from a globally representative phylogeny [31].

Similarly, we find that tip lengths from persistently infected individuals are significantly longer than those of their matched controls, suggesting greater molecular change compared to acutely infected individuals (*β̂* = −2.24 nucleotide substitutions for acute versus persistently infected individuals, [CrI −2.69, −1.79]) (Fig. 2f). We then compared the final sequence from each persistent infection, focusing on those with at least one nonsynonymous change relative to the initial sequence, to sequences from acute cases in the COVID-19 database sampled within 11 days (the upper limit of the virus’ generation time) [32]. Among the persistent cases, 10 matched exactly (0 differences) with 33 sequences in the database. 17 cases matched with one difference (248 sequences), and 27 matched with two differences (5239 sequences), suggesting that onward transmission from persistent SARS-CoV-2 infections is plausible, even in the presence of nonsynonymous changes.

The most common recurrent mutations observed between the first and last sequences in persistent infections were: S:E340K (nucleotide substitution G22580A; *n* = 15), ORF1a:T1638I (C5178T; *n* = 9), ORF1ab:L3829F (C11750T; *n* = 9), S:N405D (A22775G; *n* = 8), ORF1ab:G5063S (G15451A; *n* = 8), and S:R346T (G22599C, *n* = 7) (Fig. 3d) (Supplementary Table S6). These broadly correspond to those observed when comparing all pairs of consecutive sequences (Supplementary Table S7). Notably, S:N405D was also the most frequent recurrent mutation in another large persistent infection study, and forms part of the BA.2 spike protein. S:E340K has been linked with sotrovimab resistance, particularly in immunocompromised patients [5, 15]. We also noted several recurring amino acid changes that have previously been linked with monoclonal antibody resistance, including S:R346T, S:K444R (A22893G, *n* = 5), S:R346I (G22599T, *n* = 3), S:K356T (A22629C, *n* = 3) and S:S494P (T23042C, *n* = 2) [7, 10, 12–14, 16, 34–36]. ORF1ab:L3829F (C11750T) has also been associated with nirmatrelvir/ritonavir treatment [37], and ORF1ab:V5184I (G15814A, *n* = 4 when comparing consecutive sequences [Supplementary Table S7]) with resistance to remdesivir [38–40]. Similarly, we observed several amino acid changes associated with molnupiravir resistance, including S:K440R (A22881G, *n* = 3) and S:P9L (C21588T, *n* = 3) [41, 42]. ORF1ab:G5063S (G15451A) has also been associated with higher mortality in Delta patients [43]. Lastly, we also observed several other mutations previously identified in immunocompromised patients, including ORF1a:T1638I (C5178T), E:T30I (C26333T, *n* = 4), S:N417I (A22812T, *n* = 4) and S:T95I (C21846T, *n* = 3) [2, 6, 7, 11].

### Risk factors associated with within-host substitutions

To identify risk factors associated with different numbers of synonymous and nonsynonymous changes, we compared the first and last sequence for each persistent infection. A baseline model was developed using a zero-inflated negative binomial regression, with stepwise AIC (Akaike information criterion) employed to identify relevant characteristics. The model regressed these characteristics against the number of synonymous and nonsynonymous changes, adjusting for the time between samples as a covariate (Supplementary Table S9). For every additional day between samples, the number of absolute nonsynonymous changes across all coding region amino acid sites increased by 0.023 (Standard error (SE) = 0.002, *p <* 2 × 10^−16^); similarly, the number of synonymous changes increased by 0.019 (*SE* = 0.003, *p* = 1.27 × 10^−13^). Compared to the Alpha variant, genomes in individuals infected with Delta, Omicron BA.1, BA.2, and BA.5 showed significantly more nonsynonymous changes: Delta (*β̂* = 1.751, *SE* = 0.381, *p* = 4.25 × 10^−6^), BA.1 (*β̂* = 1.446, *SE* = 0.437, *p* = 9.39 × 10^−4^), BA.2 (*β̂* = 1.620, *SE* = 0.381, *p* = 2.15 × 10^−5^), and BA.5 (*β̂* = 2.100, *SE* = 0.400, *p* = 1.59 × 10^−7^). In contrast, rates of synonymous changes did not differ significantly between these variants. This difference likely reflects varying selection pressures as novel treatments, including monoclonal antibody and antiviral therapies, became more prevalent with later variants [44, 45]. While increasing vaccination rates could also increase selection pressure, we found no evidence that the number of vaccinations was associated with the amount of (non)synonymous change (Supplementary Table S12) or indeed the genome-wide substitution rate (Supplementary Table S13), resulting in their omission for the baseline model.

Notably, older age groups exhibited a smaller number of both nonsynonymous and synonymous changes in the viral genome compared to those in the 15-30 age group (Supplementary Table S9). Specifically, virus genomes in individuals aged 45-60 (nonsynonymous *β̂* = −0.842, *SE* = 0.301, *p* = 0.005; synonymous *β̂* = −1.063, *SE* = 0.388, *p* = 0.006), 60-75 (nonsynonymous *β̂* = −1.237, *SE* = 0.257, *p* = 2.15 × 10^−5^; synonymous *β̂* = −1.273, *SE* = 0.315, *p* = 5.26 × 10^−5^), and 75+ (nonsynonymous *β̂* = −1.127, *SE* = 0.273, *p* = 1.59 × 10^−5^; synonymous *β̂* = −1.451, *SE* = 0.342, *p* = 2.18 × 10^−5^) showed lower numbers of nonsynonymous and synonymous changes than in the 15-30 years group. Lastly, no association was found between CCI and the number of synonymous changes. However, there was evidence suggesting that higher CCI scores were linked to an increase in nonsynonymous changes (*β̂* = 0.145, *SE* = 0.046, *p* = 0.002). Similar results were found when only using pre-pandemic diagnoses (Supplementary Table S11).

We also examined the impact of various diagnoses on the number nonysynonymous and synonymous changes, adjusting for time between samples, age and major variant. Genomes in individuals with an immunosuppression diagnosis had significantly more nonsynonymous changes than those without (*β̂* = 0.564, *SE* = 0.099, *p* = 1.30 × 10^−8^), while showing similar numbers of synonymous changes (*β̂* = 0.027, *SE* = 0.152, *p* = 0.859) (Table 3). Other diagnoses, including autoimmune disorders (nonsynonymous *β̂* = −0.010, *SE* = 0.201, *p* = 0.960; synonymous *β̂* = −0.198, *SE* = 0.318, *p* = 0.533), diabetes (nonsynonymous *β̂* = −0.170, *SE* = 0.178, *p* = 0.338; synonymous *β̂* = −0.435, *SE* = 0.324, *p* = 0.180), and immune-mediated inflammatory diseases (IMID) (nonsynonymous *β̂* = −0.339, *SE* = 0.238, *p* = 0.155; synonymous *β̂* = −0.006, SE = 0.321, *p* = 0.986), were not associated with increased numbers of either nonsynonymous or synonymous changes. Results were similar when only using pre-pandemic diagnoses (Supplementary Table S10).

**Table 3.** Regression results for nonsynonymous and synonymous changes by diagnostic group. Coefficients (*β̂*), standard errors (SE), and p-values are from a zero-inflated negative binomial regression model, with each model adjusted for time between samples, major variant, and age group. Diagnoses are based on contemporary data (up to December 31, 2021). Estimates denote the absolute number of (non)synonymous changes across all coding regions relative to the reference group.

| Diagnosis Group<sup>†</sup> | Nonsynonymous Changes |  |  | Synonymous Changes |  |  |
| --- | --- | --- | --- | --- | --- | --- |
| | $\hat{\beta}$ | SE | p-value | $\hat{\beta}$ | SE | p-value |
| **Model 1: Immunosuppression** | 0.564 | 0.099 | $1.30 \times 10^{-8}$ | 0.027 | 0.152 | 0.859 |
| **Model 2: Autoimmune** | -0.010 | 0.201 | 0.960 | -0.198 | 0.318 | 0.533 |
| **Model 3: Diabetes** | -0.170 | 0.178 | 0.338 | -0.435 | 0.324 | 0.180 |
| **Model 4: IMID** | -0.339 | 0.238 | 0.155 | -0.006 | 0.321 | 0.986 |
| Adjusted for time between samples, major variant and age group. |  |  |  |  |  |  |
| IMID = Immune-mediated inflammatory diseases. SE = Standard Error. |  |  |  |  |  |  |
| <sup>†</sup> Using contemporary diagnoses i.e., those registered up to 2021-12-31, for which diagnostic data is available |  |  |  |  |  |  |

Among individuals with amino acid substitutions in the viral genome associated with monoclonal antibody resistance, the majority also had an immunosuppression diagnosis (Supplementary Table S8). Similarly, for S:K440R (3*/*3, 100%) and S:P9L (3*/*3, 100%), both associated with molnupiravir resistance, all individuals had an immunosuppression diagnosis. For substitutions associated with nirmatrelvir/ritonavir resistance, such as ORF1ab:L3829F, 4*/*7 (57.1%) of individuals had an immunosuppression diagnosis (Supplementary Table S8). 58.3% (7*/*12) of individuals with a monoclonal antibody resistance-linked substitution—specifically among those with infections ending prior to mid-2022, out of a total of 34 individuals (including those with infections ending beyond this point), which is the cut-off for available hospitalization data—were hospitalized at some point during their infection.

Additionally, the overall rebound rate—defined as individuals with a negative RT-PCR test followed by a subsequent positive test during their infection—was 18.2% (55*/*303) across all persistent infections. Among non-immunosuppressed individuals, 20.4% (38*/*186) experienced rebound, compared to 14.5% (17*/*117) of immunosuppressed individuals. Using a Bayesian logistic regression model, we found no evidence of a difference in the likelihood of rebounding between immunosuppressed and non-immunosuppressed individuals (*β* = −0.28, CrI: −0.72 to 0.15) (Supplementary Table S16), even after adjusting for infection duration (*β* = −0.29, CrI: −0.75 to 0.15) (Supplementary Table S17). Additionally, rebounding was not associated with the substitution rate (Supplementary Table S18).

## Discussion

In this study, we identified risk factors associated with persistent SARS-CoV-2 infections by analyzing a comprehensive dataset from the Danish national sequencing database, including detailed associated metadata. Our results indicate that older age groups, particularly individuals aged 45 years and above, are markedly more likely to develop persistent infections compared to younger individuals. Additionally, we find that individuals with an immunosuppression diagnosis face over five times the risk of experiencing persistent infections, underscoring the heightened vulnerability of this population. Our analysis also reveals several mutations associated with antibody and antiviral resistance, and significant selective pressures in several coding regions not observed in acute infections, suggesting ongoing viral adaptation. Notably, immunosuppression appears to contribute to an increased accumulation of nonsynonymous mutations, whereas these dynamics were not associated with other disease groups.

Our findings on sociodemographic characteristics are consistent with those of other studies, which also report that older age groups and males are at a higher risk of developing persistent infections upon testing positive [18]. Importantly, our findings also provide evidence that full vaccination, prior to immune waning, reduces the likelihood of an infection becoming persistent. This builds on previous findings suggesting that vaccination reduces viral diversity during infection, limiting the exploration of protein sequence variations [46–48]. This could lower the likelihood of novel variants and immune escape, thereby helping to prevent infections from becoming persistent.

We found that immunosuppression and autoimmune disorders were risk factors for persistent infections, whereas no association was observed between diabetes, IMID, and persistent infections. This aligns with other research suggesting that adaptive SARS-CoV-2 evolution frequently occurs in individuals with immunosuppression, particularly those with HIV or cancer [1, 49–54]. In the case of HIV, these findings underscore the critical importance of initiating antiretroviral therapy to restore CD4+ T cell responses, which are essential for neutralizing antibody activity and the clearance of SARS-CoV-2 infection [49, 50]. While the increased risk associated with autoimmune disorders attenuated when adjusting for all other diagnostic groups, we cannot rule out an association, which could plausibly be due to the use of immunosuppressant medications in these individuals, inhibiting their ability to clear the virus.

Notably, genomes of individuals with immunosuppression harbored significantly more nonsynonymous changes upon infection, while other diagnoses showed no association with these changes. Many of these recurrent nonsynonymous changes have previously been linked to resistance against monoclonal antibody [7, 10, 12–14, 16, 34–36] and antiviral treatments, including nirmatrelvir/ritonavir [37], remdesivir [38–40] and molnupiravir [41, 42], and have a higher between-host fitness on average compared to single-occurring and unrealized mutations at the same sites. This also highlights the need to vaccinate individuals with immunosuppression and take measures to limit onward transmission until viral shedding has been conclusively resolved [51].

However, due to limited data on medication administration, we cannot definitively determine whether this adaptive evolution is driven by compromised immune systems that allow for tolerance of novel viral trajectories or increased selection pressure from more aggressive treatment strategies. Supporting a role for treatment-induced selection, most individuals with monoclonal antibody resistance-linked substitutions had an immunosuppression diagnosis, further reinforced by the similar numbers of synonymous changes. Nevertheless, among those with infections ending prior to mid-2022, less than sixty percent of these cases involved hospitalized individuals, suggesting that observed viral evolution is not solely attributable to in-hospital treatment. Furthermore, we found no evidence of differing rebound rates between individuals with immunosuppression and without immunosuppression, suggesting that immunosuppression alone is unlikely to drive the emergence of adaptive drug-resistant mutations that could facilitate infection rebound. Further evidence is therefore required to disentangle the extent to which these dynamics in individuals with immunosuppression are due to specific treatment regimes or tolerance due to compromised immune systems.

Our analysis also revealed notable patterns of selection pressure across the SARS-CoV-2 genome. While no evidence of genome-wide positive selection was found, we observed significant selective forces in specific coding regions, such as the spike (S) region, which was not observed in acute infections. Additionally, we detected purifying selection in acute infections, particularly in the ORF1ab region. These findings indicate that adaptive evolution in certain genomic regions may be shaped and constrained by selective pressures imposed by the host immune response and antiviral treatments. Importantly, the low genomic distances between sequences from persistent infections and those circulating at the same time suggest that onward transmission from persistent SARS-CoV-2 infections is plausible. Comparisons of exact matches [55, 56], which leverage clusters of identical sequences to infer transmission patterns, further highlight the role of persistent infections in both generating new variants and seeding their spread.

Although irregular testing intervals prevented us from estimating population-wide rates of persistent infections, their prevalence is likely higher than previously thought. Conservative estimates from the United Kingdom suggest that 0.1–0.5% of infections may become persistent [7], underscoring their potential as sources of new variants. Given the strict criteria used to define persistent infections in this study and the non-random testing strategy, the number of cases identified likely represents a significant underestimate of the true burden in the Danish population. Nearly one in five individuals in our cohort experienced viral rebound during their persistent infection. These dynamic patterns in viral load, as others have noted, further suggest that population-level reinfection rates may be overestimated when defined solely by the presence of a negative test [7]; this highlights the challenges of tracking potentially novel mutations arising from these infections, particularly given their frequency and duration.

This study has several limitations. First, unmeasured confounding factors could obscure the relationship between sociodemographic factors and the risk of persistent infection. Additionally, the absence of medication data prevents us from establishing causal links between treatment and the emergence of specific nonsynonymous changes. Second, our findings may primarily reflect individuals who undergo frequent testing, potentially due to specific occupations, socioeconomic factors, or hospitalization, which leads to more frequent testing. This is particularly relevant for infections occurring from 2022 onwards, when systematic community testing was scaled back. As a result, the study population may not accurately represent the true distribution of persistent infections in the broader population. Finally, while the case-control design was well-suited for studying the rare outcome of persistent infections, it may still be subject to selection biases that could affect the robustness of the results. To mitigate this, we employed a matching procedure that ensured comparability between cases and controls in terms of both testing behaviors and variant.

Overall, our study highlights the potential role of age and health conditions, particularly immunosuppression, as risk factors for persistent SARS-CoV-2 infections and contributors to viral mutation during such infections. These results emphasize the importance of closely monitoring the evolutionary trajectories of persistently infected individuals, especially those who are immunosuppressed and receiving ongoing treatment, to track the emergence of potentially novel variants.

## Methods

### Testing, genomic data and sequencing in Denmark

Large-scale sequencing of SARS-CoV-2 was a key strategy in response to COVID-19 in Denmark. The Danish COVID-19 Genome Consortium (DCGC) - formed in March 2020 - undertook sequencing efforts to monitor the evolution of SARS-CoV-2. The DCGC included all genomic sequences that met the inclusion criteria based on a cycle threshold (Ct) value, which ranged from 30 to 38 during the study period. Whole genome sequencing (WGS) was conducted by multiple institutions, including Aalborg University (AAU), *Statens Serum Institut* (SSI), and regional clinical microbiology laboratories. Sequencing workflows commonly employed whole genome amplification using a modified version of the ARTIC tiled PCR approach [57] (https://artic.network), generating overlapping amplicons ranging from approximately 1000 to 1500 base pairs in length. A custom 2-step PCR method facilitated the barcoding of amplicon libraries, which were then normalized, pooled, and prepared for sequencing using Oxford Nanopore’s SQK-LSK109 ligation kit. Sequencing was primarily performed on the MinION platform with Oxford Nanopore R.9.4.1 flow cells [58]. Raw data was base-called using `Guppy` (https://nanoporetech.com) and demultiplexed via a custom `cutadapt` v.2.10 wrapper [59]. Consensus sequences were generated with the `artic minion` function, following ARTIC network protocol defaults (v.1.1.0) and incorporating `medaka` [60] for consensus calling.

Additional sequencing workflows utilized the ARTIC v3 amplicon sequencing panel [61], which consisted of 98 overlapping amplicons of approximately 300 nucleotides, sometimes including custom spike-ins to maintain consistent coverage [62]. This sequencing approach was executed on Illumina platforms, such as the NextSeq or NovaSeq, with paired read lengths ranging from 51 to 150 nucleotides (most often 74 nucleotides). Sequencing reads underwent trimming with `trim-galore` v.0.6.10 (https://github.com/FelixKrueger/TrimGalore), with consensus sequences generated using `iVar` [63] (v.1.4.3) or a combination of `iVar` and custom `BCFtools` v.1.18 commands. A PHRED score threshold of 20 was applied during read and primer trimming. Sequences from regional labs employing either Nanopore or Illumina sequencing were also integrated into national sequence databases, provided they met quality criteria: fewer than 3000 ambiguous bases (N’s), ≤ 5 ambiguous base calls, high yield relative to controls, and passing QC standards that detected contamination. Sequence integrity was verified against SARS-CoV-2 reference models, with tools such as VADR [64], prior to being uploaded to sequence data repositories. Metadata related to sequencing included information on sampling and sequencing dates. The sequences were aligned to the Wuhan WIV04 (MN996528.1) reference genome [65] using `MAFFT` (v.7.520) [66]. Following the alignment, problematic regions identified by De Maio et al. [67] were masked using the *augur mask* tool from the *Nextstrain* pipeline [68] (Augur version v.22.3.0). This masking process included regions at both the 5’ end (positions 1-55) and the 3’ end (from site 28804 onwards).

### Defining and identifying persistent infections

To identify possible persistent infections, we adopted the method used by Ghafari et al. [7] In short, possible persistent infections were defined as infections spanning 26 days or more (i.e., two or more positive tests within the same major variant, ≥ 26 days and *<* 120 days apart, identified using Pangolin v4.3.1). The original justification for the 26 day cut-off by Ghafari et al. [7] was based on the observation that most individuals with acute infection shed the virus for less than 20 days and no longer than 30 days in the respiratory tract [69, 70]. Infections were defined as all positive tests within this period, from the first to the last positive test for a given variant. Only infections that began before January 1, 2023, were included to ensure that vaccination data were available for all individuals.

Additionally, the primary lineages (and respective sub-lineages) we analyzed included Alpha (B.1.1.7), Delta, Omicron BA.1, Omicron BA.2, and Omicron BA.5. We assessed whether all sequences from each individual-level infection were from the same infection by checking for shared rare single-nucleotide polymorphisms (SNPs) at consecutive time points relative to the population for the given major variant. SNPs were classified as rare if their frequency in the population-level dataset fell below a specified threshold. To establish a threshold that maximized the identification of persistent infections while minimizing false positives, we generated a dataset of 1,000 randomly paired sequences from different individuals with the same major variant in the population dataset, ensuring they were sampled 26 days apart. We then assessed the proportion of incorrectly identified persistent infections at different thresholds for defining rare SNPs. Although raising the threshold resulted in a higher number of identified persistent infections, it also increased the false-positive rate among the randomly paired sequences at excessively high thresholds. Given the strict definitions for persistent infections, we expect the number of identified persistent infections to be an under-estimate of the true number. By testing a wide range of thresholds using logarithmic spacing for 100 different values (Supplementary Fig. S3), we established the threshold at 10.9% (i.e., the frequency of the given SNP is below the threshold among all other population-level sequences within the same major variant). This resulted in false-positive rates of 3.1% (Alpha), 3.0% (Delta), 3.3% (Omicron BA.1), 2.1% (Omicron BA.2), and 2.2% (Omicron BA.5).

### Registry Linkage

To associate sequences with Danish registry data, pseudo-anonymized Danish civil registration numbers (CPR numbers) were employed to connect sequences to the relevant registry information maintained by Statistics Denmark. This registry data included sex, age, test date, and vaccination status. The following vaccinations were administered as part of the Danish vaccination program: Comirnaty (Pfizer/BioNTech), Spikevax (Moderna), Vaxzevria (AstraZeneca), and COVID-19 Vaccine Janssen (Johnson & Johnson) [28, 71]. Individuals were considered vaccinated at the time of infection onset if they had received one, two, or three doses, with at least seven days having passed since their most recent dose.

For diagnostic codes to identify health conditions, we linked CPR numbers to the National Patient Register [72] to obtain relevant in- and out-patient ICD-10 codes for immunosuppression, autoimmune disorders, Immune-Mediated Inflammatory Diseases (IMID) and diabetes using previously defined codelists. A full list of ICD-10 codes by health condition can be found in Supplementary Tables S19, S20, S21 and S22. The National Patient Register includes data on individuals who have been admitted to somatic hospital departments since 1977 and ambulatory care from 1995. Individuals were considered part of a disease group (i.e. binary) if they had an ICD-10 code in the relevant disease group. Charlson Comorbidity Index (CCI) [30] scores were calculated using the `heaven` package [73], with the relevant ICD-10 codes and weightings defined by the Danish Multidisciplinary Cancer Group (DMCG) [74, 75]. CCI values were computed using both 5-year and 10-year look-back periods from the start of the infection. Hospitalization was defined as an admission lasting 12 hours or longer.

### Case-control design

To identify risk factors for persistent infections, we designed a case-control study in which individuals with a persistent infection served as cases, and other individuals within the Danish testing database were considered potential controls. Controls were identified by matching each case to individuals with a positive PCR result for the same major variant, within ±1 day of the case’s infection start date. This matching was crucial to ensure comparability between cases and controls in terms of testing behaviors as well as variant. Controls were defined as individuals with a negative PCR test within 26 days and no positive tests during the 120 days following their positive test date, confirming that they were not persistently infected.

To investigate risk factors for persistent infections, we used Bayesian multivariable conditional logistic regression [29], accounting for the matched design by including strata for each case-control set. We developed five key models: baseline, immunosuppression, autoimmune disorders, IMID, and diabetes. A weakly informative prior N(0, 1) was placed on all regression coefficients; the model was fit using four Markov chains with 2,000 iterations each, including a warm-up phase of 1,000 iterations. Posterior summaries were generated, including point estimates and 95% credible intervals based on the 2.5th and 97.5th percentiles of the posterior distributions. Models were run using `brms` [33] (v.2.20.3). The baseline model included covariates age group (0–15 [0 ≤ age *<* 15], 15–30 [15 ≤ age *<* 30], 30–45 [30 ≤ age *<* 45], 45–60 [45 ≤ age *<* 60], 60–75 [60 ≤ age *<* 75], 75+ [age ≥ 75]), sex (female, male), vaccination status (which combines the number of vaccinations at infection onset and the time since the last vaccination *<* 18 weeks or ≥ 18 weeks) for those with two or three vaccinations), and the Charlson Comorbidity Index (CCI). The 18-week cut-off was chosen to account for immunity, which typically wanes between three and four months post-vaccination, followed by a stabilization period [76–78]. Two and three vaccinations were grouped due to policy changes during the pandemic, during which both were considered fully vaccinated at different stages.

In the models for immunosuppression, autoimmune disorders, IMID, and diabetes, the same covariates were used, but CCI was replaced by the relevant condition for each model. Major variant was excluded in the models due to matching individuals by test date and major variant. For baseline models, we used pre-pandemic diagnoses (i.e., those prior to 2020-01-01) to clarify the temporal relationship between vaccinations and relevant diagnoses, including Charlson Comorbidity Index (CCI) values. Sensitivity analyses examined the impact of using contemporary diagnoses (i.e., those registered up to 2021-12-31, for which diagnostic data is available). Sensitivity analyses further explored interactions between covariates to understand their combined influence on persistent infections. CCI values were also tested using both 5-year and 10-year look-back periods from the start of infection.

### Characterizing mutations and viral load trajectories

For consecutive within-host consensus sequences, we used a custom code to identify mutation sites in `R` version 4.2.3. To assess changes in viral load during persistent infections, we compared Ct values from the initial sequence collection to those at the final available sequencing time point. The distribution of Ct differences was approximately normal (Shapiro-Wilk test [*W* = 0.976, *p* = 0.192)]) [79]. We modeled the difference in Ct values using a Bayesian approach implemented in `brms`, specifying a weakly informative normal prior, N(0, 1), on the mean Ct difference.

To assess the between-host fitness effects of individual amino acid substitutions, we used a model developed by Bloom et al. [31], which estimates the fitness effects of specific amino acid changes based on a global phylogeny. These fitness effects are inferred by comparing observed versus expected mutation counts. Data for specific amino acid changes is available here: https://github.com/jbloomlab/SARS2-mut-fitness/blob/main/results/aa_fitness/aa_fitness.csv. We then compared (a) the fitness of recurrent mutations (those occurring two or more times in our dataset) versus single-occurrence mutations observed in persistently infected individuals, and (b) the fitness of recurrent mutations against other amino acids observed at the same sites (i.e., comparing each recurrent mutation to other amino acids not observed at the same position). We constructed a Bayesian linear model to evaluate fitness differences between recurrent mutations and their respective comparison groups, specifying a weakly informative prior (*N* (0, 1)) for the regression coefficients, with model fit using `brms`.

To compare tip lengths between persistent and non-persistent infections, we constructed a phylogenetic tree for each set of cases and controls (from the conditional logistic regression) with at least 10 controls (n=293). For each case-control set, we performed phylogenetic analysis using IQ-TREE (v1.6.12) with a GTR+F substitution model. The Wuhan WIV04 (MN996528.1) reference sequence [65] was used as the outgroup.

For Ka and Ks difference calculations, we used the `kaks` function from the `seqinr` package (v4.2.23), analyzing sequences from both persistent and non-persistent infections. For persistent infections, we selected the first and last sequence from each infection episode. For non-persistent infections, sequences from control individuals identified through conditional logistic regression analysis were used. Individuals with two or more sequences (starting from their matched infection date) were included, and we analyzed the first and last sequence within the infection episode, defined as sequences collected within 26 days of the infection start date. Alignments were split by coding region to calculate region-specific Ka and Ks values. To compare the differences in Ka and Ks values, we assumed a normal distribution and conducted a Bayesian analysis. The model was implemented using the `brms` package, with a weakly informative prior (*N* (0, 0.01)) specified for the intercept. Model fitting was performed using four chains, each with 2,000 iterations, including a warm-up phase of 1,000 iterations.

### Characteristics associated with number of (non)synonymous mutations

To investigate factors associated with the number of synonymous and nonsynonymous mutations per infection, we analyzed the initial and final viral sequences from each case, calculating the number of synonymous and nonsynonymous substitutions over the infection period. We used a zero-inflated negative binomial regression model to account for excess zeros and overdispersion, regressing mutation counts on host characteristics, including age and major variant, while adjusting for the time interval between sample dates. Variable selection was performed using stepwise AIC with the *MASS* [80] package (v7.3.58.2) in *R*.

We further examined the influence of specific diagnoses, such as immunosuppression, autoimmune disorders, IMID, and diabetes, by incorporating these conditions as covariates in separate models; in these, we adjusted for age, sampling interval, and major variant. Sensitivity analyses were conducted using pre-pandemic diagnoses (registered before 2020) for both individual diagnosis models and for CCI scoring in the baseline model. To identify episodes of viral rebound, we used the national SARS-CoV-2 testing database to find negative test results during the individual’s infection. For Bayesian logistic regression models comparing the likelihood of rebounding, a weakly informative prior N(0, 1) was placed on regression coefficients. The model was fit using four Markov chains with 2,000 iterations each (1,000 warm-up), implemented in `brms`.

## Data availability

The data utilized in this study is accessible under restricted conditions under Danish data protection laws, since the information pertains to sensitive individual-level data. Researchers can request access to the data, including SARS-CoV-2 consensus genomes, from The Danish Health Data Authority and Statens Serum Institut, complying with Danish data protection regulations and any necessary permissions. No data collection or sequencing was conducted specifically for this study.

## Code availability

All code relevant to reproducing the experiments is available online: https://github.com/MLGlobalHealth/sars-cov2-persistent.

## Data Availability

https://github.com/MLGlobalHealth/sars-cov2-persistent

## Author contributions

MPK, AK, DD and SB conceived and designed the study. MPK, AK, NS, MMBS and JCS performed the analyses. PJ, MHET, LAEH, SL, MR, MS, TGK, and LHM were responsible for data curation and management. NMF, CM and MG helped with data analysis. All authors contributed to editing the original draft.

## Competing interests

The authors declare no competing interests.

## Ethical statement

This study was conducted using data from national registers only. According to Danish law, ethics approval is not needed for this type of research. All data management and analyses were carried out on Statistics Denmark’s secure research servers. The study only contains aggregated results and no personal data.

## Funding

SB, CM, CW and NMF acknowledge funding from the MRC Centre for Global Infectious Disease Analysis (reference MR/X020258/1), funded by the UK Medical Research Council (MRC). This UK-funded award is carried out in the frame of the Global Health EDCTP3 Joint Undertaking. SB, CM, and NFM are funded by the National Institute for Health and Care Research (NIHR) Health Protection Research Unit in Modelling and Health Economics, a partnership between the UK Health Security Agency, Imperial College London and LSHTM (grant code NIHR200908). Disclaimer: “The views expressed are those of the author(s) and not necessarily those of the NIHR, UK Health Security Agency or the Department of Health and Social Care.”. SB acknowledges support from the Novo Nordisk Foundation via The Novo Nordisk Young Investigator Award (NNF20OC0059309) which also supports FPL. SB acknowledges the Danish National Research Foundation (DNRF160) through the chair grant which also supports MPK, JCS, and NS. SB acknowledges support from The Eric and Wendy Schmidt Fund For Strategic Innovation via the Schmidt Polymath Award (G-22-63345), who also support CM. DAD acknowledges funding from the Novo Nordisk Foundation Emerging Data Science Investigator Award (NNF23OC0084647). PJ and TGV acknowledge funding from EU under Grant Agreements 101094685 LEAPS and 101102733 DURABLE; views and opinions expressed do not necessarily reflect those of the EU or the granting authorities, neither the EU nor the granting authorities can be held responsible for them. M.U.G.K. acknowledges funding from The Rockefeller Foundation, Google.org, the Oxford Martin School Pandemic Genomics programme, European Union’s Horizon Europe programme projects MOOD (grant code 874850) and E4Warning (grant code 101086640), the John Fell Fund, a Branco Weiss Fellowship and Wellcome Trust grants 225288/Z/22/Z, 226052/Z/22/Z and 228186/Z/23/Z, United Kingdom Research and Innovation (grant code APP8583) and the Medical Research Foundation (MRF-RG-ICCH-2022-100069).

## Acknowledgements

The authors express gratitude to Statens Serum Institut and The Danish Health Data Authority for their efforts in gathering and granting access to the data. Additionally, appreciation is extended to the Danish Covid-19 Genome Consortium for conducting the sequencing of SARS-CoV-2 samples; the full list of members and their affiliations is listed in the Appendix.

## Appendix

**Figure S1.**
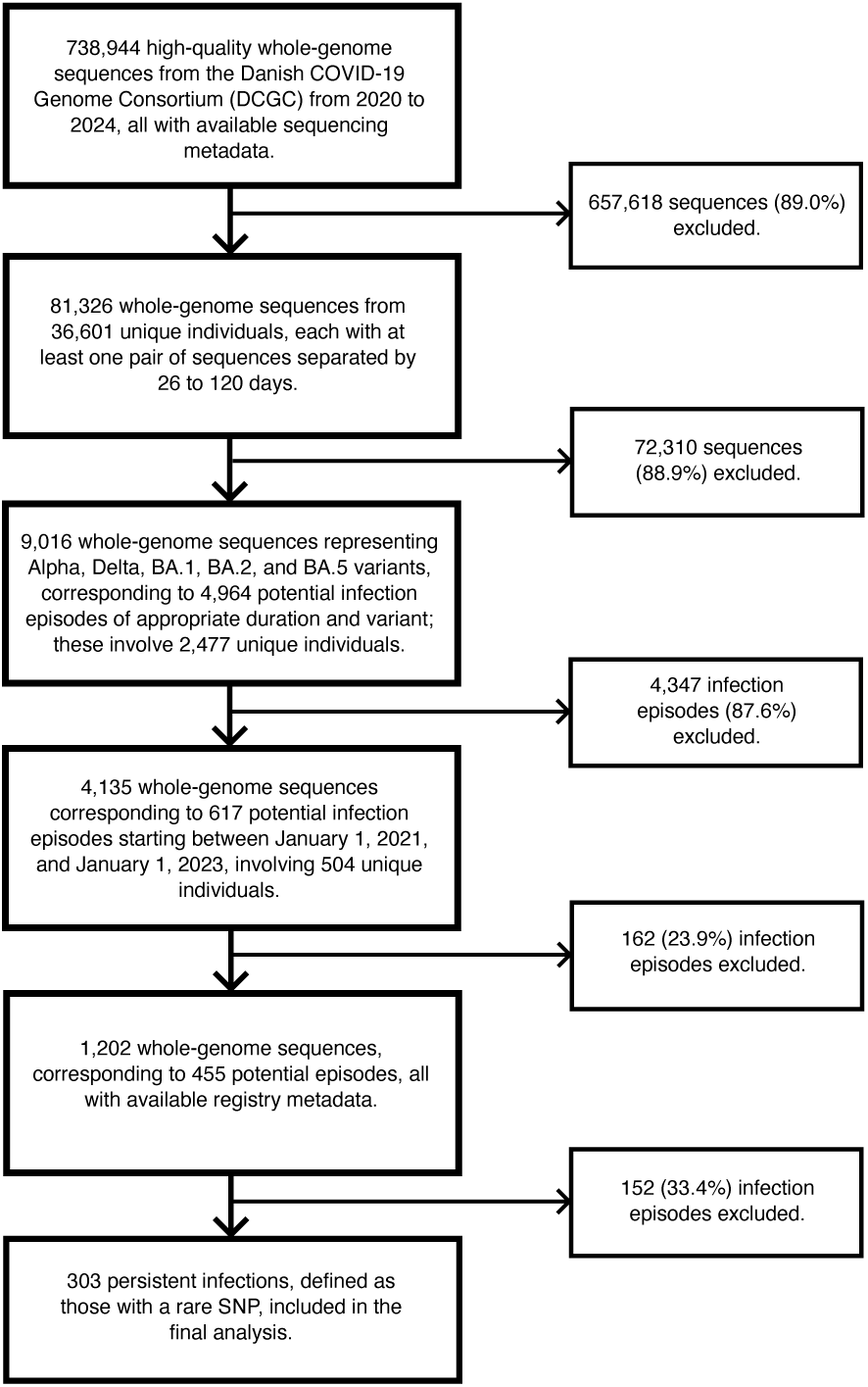
Flowchart for finding relevant sequences and defining persistent infections in this study.

**Figure S2.**
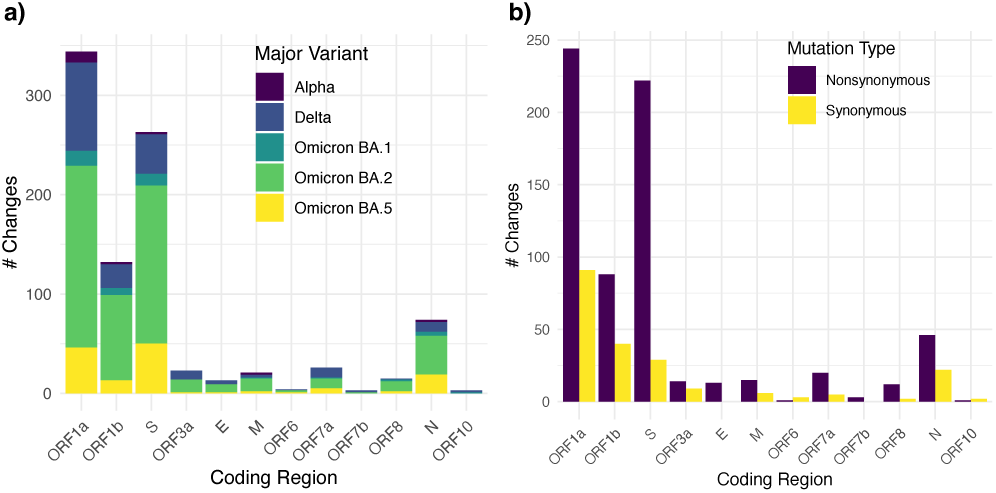
a) Number of mutations and b) (non)synonymous changes by coding region across all pairs of consecutive sequences.

**Figure S3.**
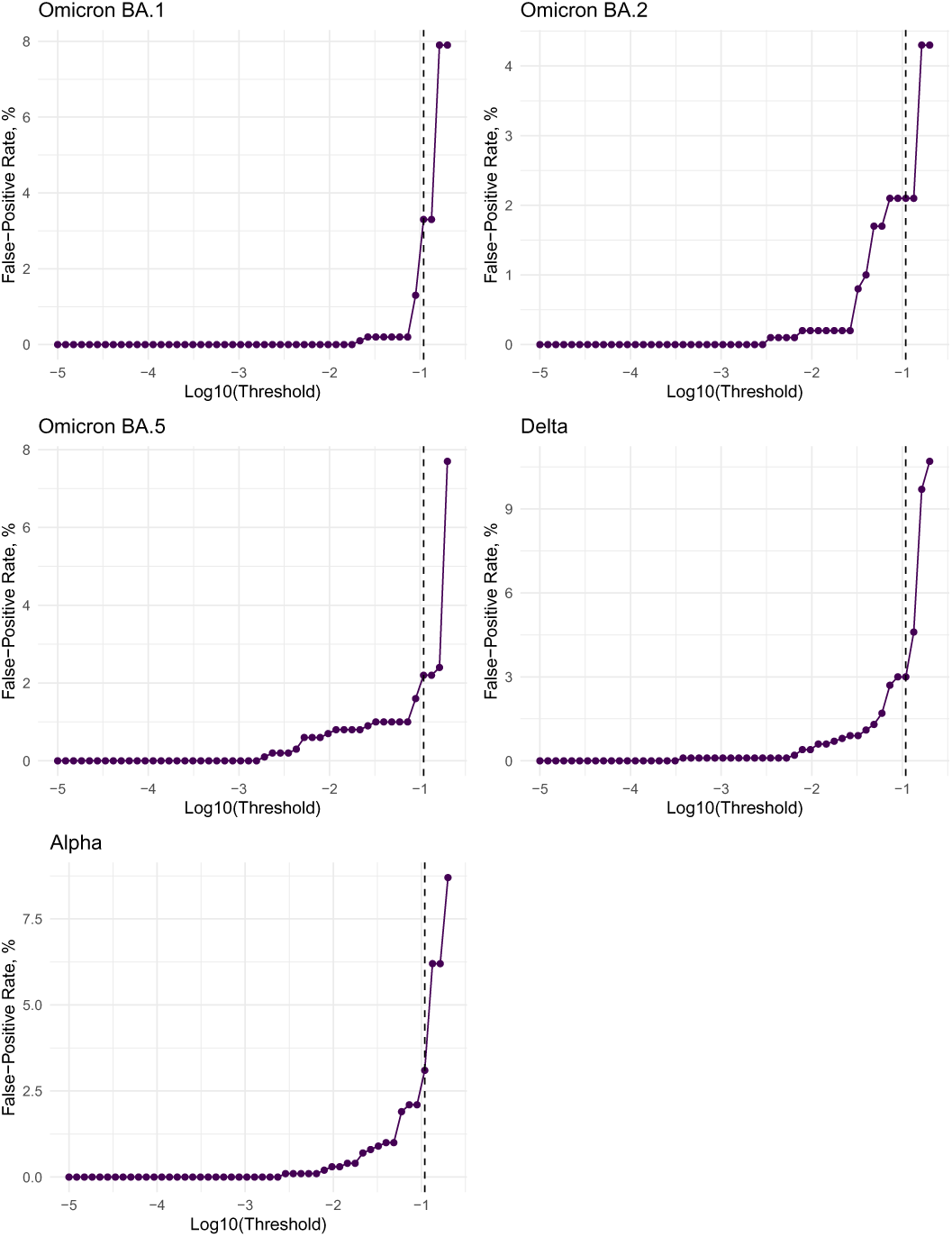
Plots with false-positive rates at different thresholds for each major variant.

**Figure S4.**
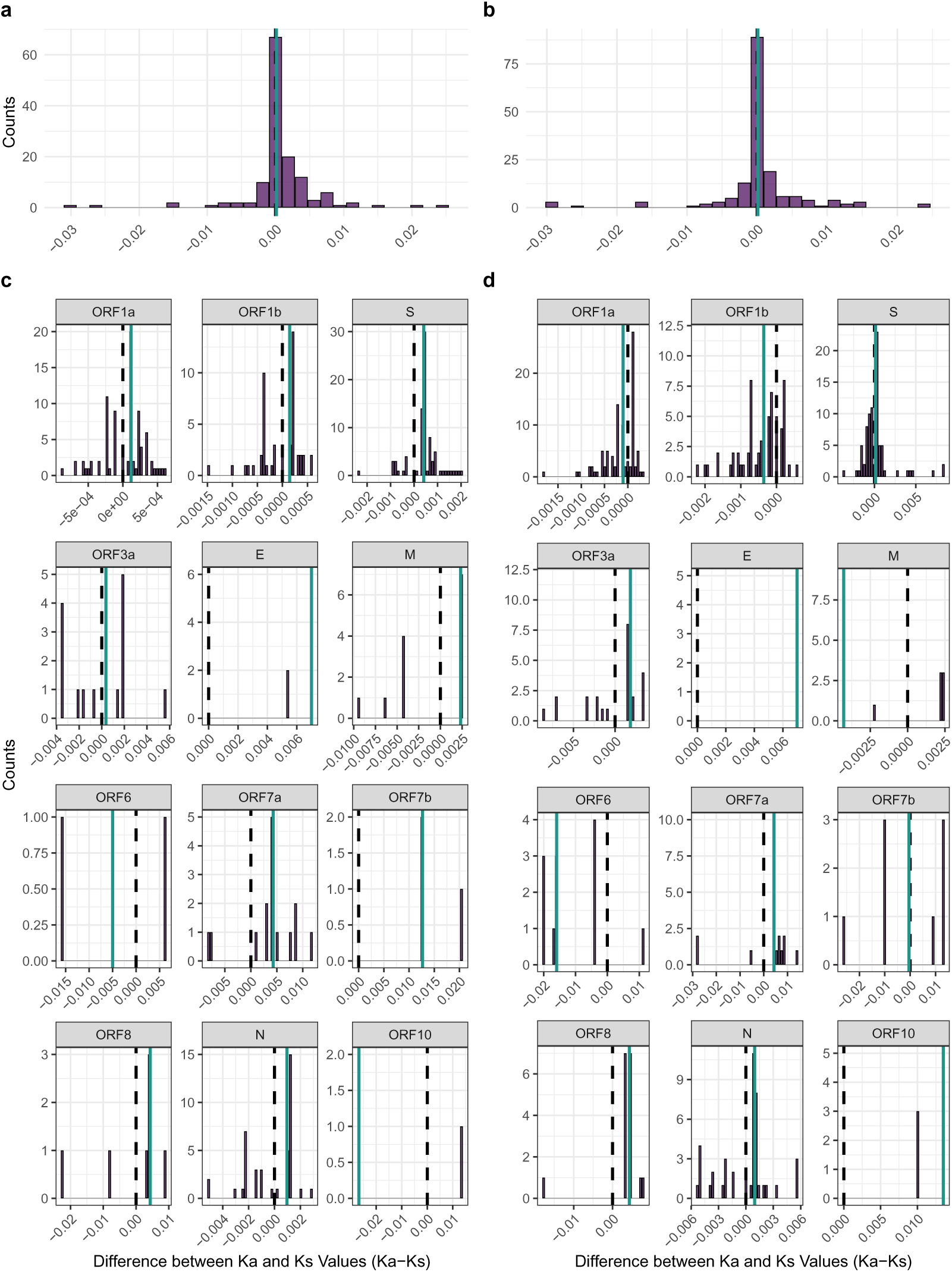
Differences in Ka and Ks values (Ka - Ks) across the genome and by coding region. (a) Difference in Ka and Ks values among individuals with persistent infections (*n* = 135). Ka and Ks represent the substitution rates per non-synonymous and synonymous site, respectively. The dotted line indicates a value of 0, and the green line represents the median. (b) Difference in Ka and Ks values among individuals without persistent infections (controls, *n* = 167) with two or more available sequences. (c) Difference in Ka and Ks values in each coding region among individuals with persistent infections, specifically those with at least one mutation in the region. (d) Difference in Ka and Ks values in each coding region among individuals without persistent infections, specifically those with at least one mutation in the region.

**Table S1.** Distribution of Variants, Age Groups, Sex, and Number of Vaccinations (n=303)

| Major Variant | Count (Percentage) |
| --- | --- |
| Alpha | 42 (13.9%) |
| Delta | 63 (20.8%) |
| Omicron BA.1 | 27 (8.9%) |
| Omicron BA.2 | 132 (43.6%) |
| Omicron BA.5 | 39 (12.9%) |

| Age Group | Count (Percentage) |
| --- | --- |
| 0-15 | 10 (3.3%) |
| 15-30 | 24 (7.9%) |
| 30-45 | 31 (10.2%) |
| 45-60 | 45 (14.9%) |
| 60-75 | 98 (32.3%) |
| 75+ | 95 (31.3%) |

| Sex | Count (Percentage) |
| --- | --- |
| Male | 194 (64.0%) |
| Female | 109 (36.0%) |

| Number of vaccinations at infection | Count (Percentage) |
| --- | --- |
| 0 | 76 (25.1%) |
| 1 | 8 (2.6%) |
| 2 | 44 (14.5%) |
| 3 | 175 (57.8%) |

**Table S2.**
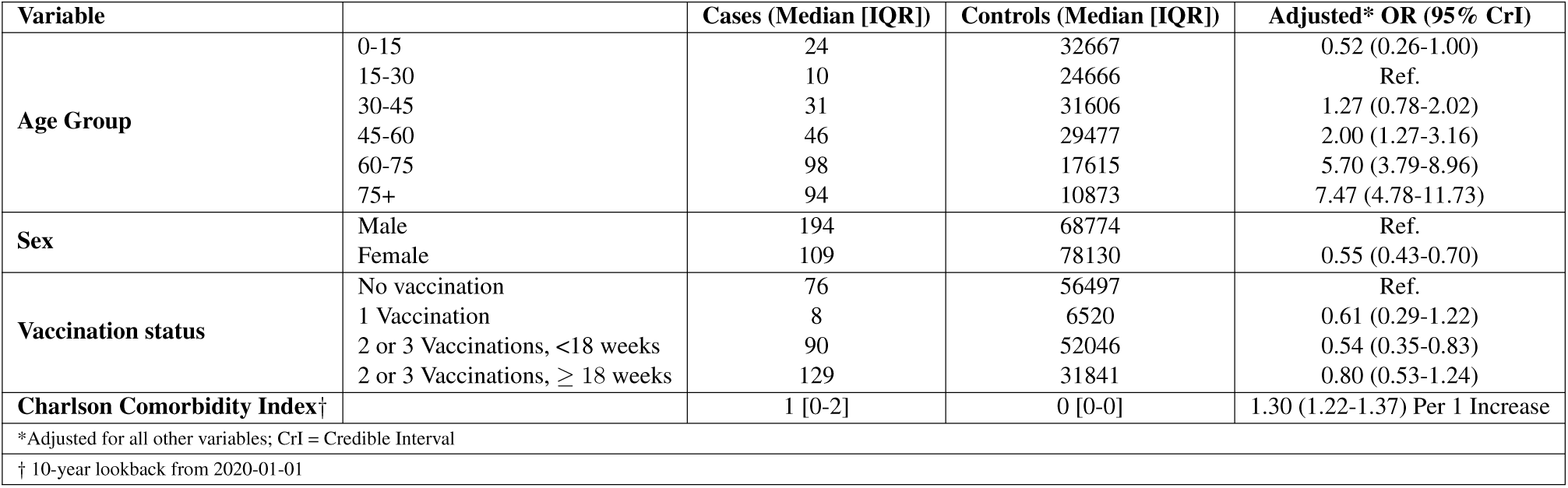
Risk factors for persistent infection. Values are presented as median [IQR] for continuous variables and n (%) for categorical variables. Adjusted odds ratios (OR) with 95% credible intervals (CrI) are provided from the conditional logistic regression output, with adjustments for all other variables. CrI = credible interval. The Charlson Comorbidity Index was calculated with a 10-year lookback period from January 1, 2020. Vaccination status combines the number of vaccinations at infection onset and the time since the last vaccination (*<* 18 weeks or *≥* 18 weeks) for those with 2 or 3 vaccinations. The total number of controls is *n* = 146,904, corresponding to 62,648 unique individuals.

**Table S3.** Diagnoses associated with persistent infection. Unadjusted and adjusted odds ratios (OR) with 95% confidence intervals (CI) are provided, with adjustments for sex, age, and number of vaccinations. IMID = Immune-mediated inflammatory diseases. The diagnoses were recorded up to December 31, 2021, for which diagnostic data is available. The total number of controls is *n* = 146,904, corresponding to 62,648 unique individuals.

| Variable |  | # Cases | # Controls | Unadjusted OR (95% CI) | Adjusted* OR (95% CI) |
| --- | --- | --- | --- | --- | --- |
| **Model 1: Immunosuppression Diagnosis** | Yes | 117 | 2477 | 10.65 (8.90-12.78) | 8.05 (6.68-9.64) |
|  | No | 186 | 144427 | Ref. | Ref. |
| **Model 2: Autoimmune Diagnosis** | Yes | 16 | 1825 | 2.34 (1.60-3.35) | 1.71 (1.15-2.41) |
|  | No | 287 | 145079 | Ref. | Ref. |
| **Model 3: Diabetes Diagnosis** | Yes | 30 | 4959 | 1.87 (1.42-2.45) | 1.14 (0.84-1.49) |
|  | No | 273 | 141945 | Ref. | Ref. |
| **Model 4: IMID Diagnosis** | Yes | 19 | 5110 | 1.36 (0.95-1.88) | 1.14 (0.80-1.56) |
|  | No | 284 | 141794 | Ref. | Ref. |
| *All adjusted for sex, age, and vaccination status. |  |  |  |  |  |

**Table S4.**
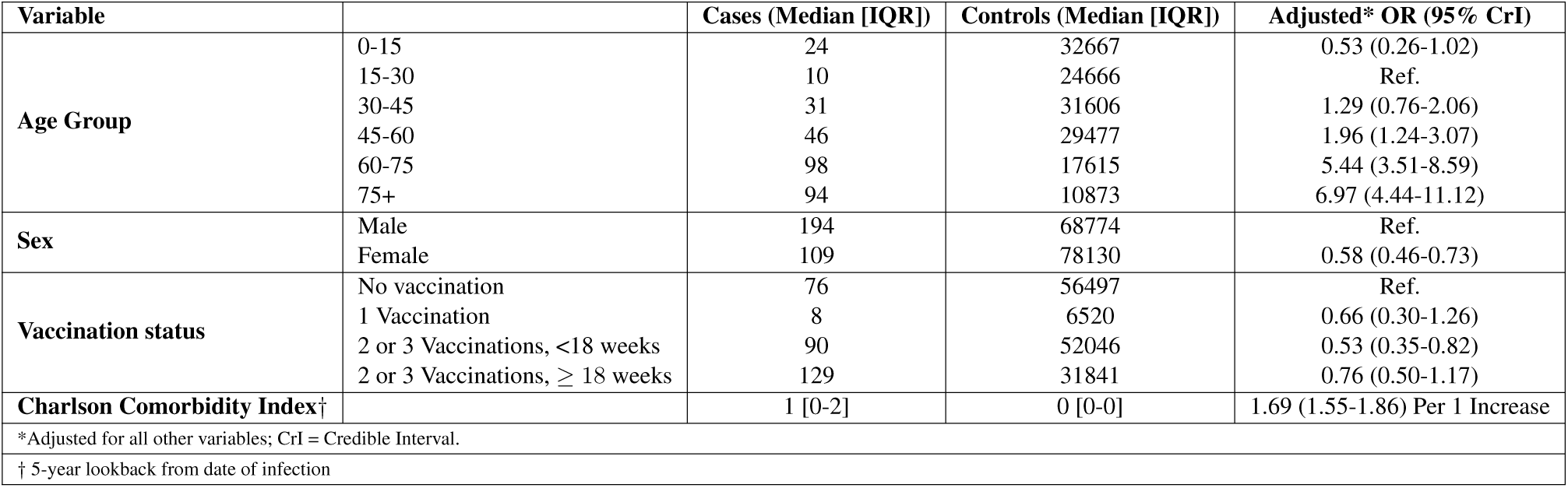
Risk factors for persistent infection. Values are presented as median [IQR] for continuous variables and n (%) for categorical variables. Adjusted odds ratios (OR) with 95% credible intervals (CrI) are presented from the conditional logistic regression output, with adjustments for all other variables. CrI = credible interval. The Charlson Comorbidity Index was calculated with a 5-year lookback period from the date of infection. Vaccination status combines the number of vaccinations at infection onset and the time since the last vaccination (*<* 18 weeks or *≥* 18 weeks) for those with 2 or 3 vaccinations. The total number of controls is *n* = 146,904, corresponding to 62,648 unique individuals.

**Table S5.** Diagnoses associated with persistent infection, adjusting for all other diagnoses. Values are presented as n for categorical variables. Unadjusted and adjusted odds ratios (OR) with 95% credible intervals (CrI) are presented from the conditional logistic regression output, adjusted for sex, age, and vaccination status. CrI = credible interval; IMID = immune-mediated inflammatory diseases. Diagnoses are based on pre-pandemic conditions (i.e., those prior to January 1, 2020). Vaccination status combines the number of vaccinations at infection onset and the time since the last vaccination (*<* 18 weeks or *≥* 18 weeks) for those with 2 or 3 vaccinations. The total number of controls is *n* = 146,904, corresponding to *n* = 62,648 unique individuals.

| Diagnosis Group† |  | Cases | Controls | Adjusted* OR (95% CrI) |
| --- | --- | --- | --- | --- |
| **Immunosuppression** | Yes | 76 | 1929 | 5.79 (4.73-7.22) |
|  | No | 227 | 144975 | Ref. |
| **Model 2: Autoimmune** | Yes | 13 | 1611 | 1.43 (0.92-2.08) |
|  | No | 290 | 145293 | Ref. |
| **Model 3: Diabetes** | Yes | 28 | 4717 | 1.00 (0.75-1.30) |
|  | No | 275 | 142187 | Ref. |
| **Model 4: IMID** | Yes | 18 | 4775 | 1.06 (0.74-1.47) |
|  | No | 285 | 142129 | Ref. |
| *Apart from adjusting for all other diagnoses, the model is also adjusted for sex, age, and vaccination status; CrI = Credible Interval. |  |  |  |  |
| † Using pre-pandemic diagnoses (i.e., those prior to 2020-01-01). |  |  |  |  |

**Table S6.** Most common recurrent mutations between first and last sequences in infection episodes.

| Mutation | Amino acid change | Counts |
| --- | --- | --- |
| G22580A | S:E340K | 15 |
| C11750T | ORF1ab:L3829F | 9 |
| C5178T | ORF1a:T1638I | 9 |
| A22775G | S:N405D | 8 |
| G15451A | ORF1ab:G5063S | 8 |
| G22599C | S:R346T | 7 |
| A22893G | S:K444R | 5 |
| C28550A | N:R93R | 5 |
| A22812T | S:N417I | 4 |
| C26333T | E:T30I | 4 |
| G5180A | ORF1ab:D1639N | 4 |
| A12759G | ORF1ab:D1323N | 3 |
| A22629C | S:K356T | 3 |
| A22881G | S:K440R | 3 |
| A23854C | S:K764N | 3 |
| C13458T | ORF1ab:S4398L | 3 |
| C21588T | S:P9L | 3 |
| C21846T | S:T95I | 3 |
| C4878T | ORF1ab:T1538I | 3 |
| C7851T | ORF1ab:A2529V | 3 |
| G22599T | S:R346I | 3 |
| G22775A | S:D405N | 3 |
| G22898A | S:G446S | 3 |
| G24570T | S:S1003I | 3 |
| G5305A | ORF1ab:L1680L | 3 |
| G569A | ORF1ab:E102K | 3 |
| G9479T | ORF1ab:G3072C | 3 |
| T22579C | S:D339D | 3 |

**Table S7.** Most common recurrent mutations observed across consecutive sequences, with each infection contributing only once to the counts. Bold mutations are ones that do not appear *≥* 3 times when comparing mutations between first and last sequences directly.

| Mutation | Amino acid change | Counts |
| --- | --- | --- |
| G22580A | S:E340K | 14 |
| A22775G | S:N405D | 12 |
| C11750T | ORF1ab:L3829F | 8 |
| G15451A | ORF1ab:G5063S | 8 |
| C5178T | ORF1a:T1638I | 7 |
| G22599C | S:R346T | 7 |
| A22893G | S:K444R | 6 |
| C28550A | N:R93R | 5 |
| C26333T | E:T30I | 5 |
| A22812T | S:N417I | 4 |
| G5180A | ORF1ab:D1639N | 4 |
| C22916A | **S:L452M** | 4 |
| G15814A | **ORF1ab:V5184I** | 4 |
| G22599T | S:R346I | 4 |
| G22775A | S:D405N | 4 |
| A23854C | S:K764N | 4 |
| C12789T | **ORF1ab:T4175I** | 3 |
| C21618T | **S:T19I** | 3 |
| C25614T | **ORF3a:S74S** | 3 |
| C29250T | **N:P326L** | 3 |
| C4230T | **ORF1ab:T1322I** | 3 |
| G22580C | **S:E340Q** | 3 |
| G22894T | **S:K444N** | 3 |
| T21618C | **S:I19T** | 3 |
| T23042C | **S:S494P** | 3 |
| A22881G | S:K440R | 3 |
| C13458T | ORF1ab:S4398L | 3 |
| C21588T | S:P9L | 3 |
| C21846T | S:T95I | 3 |
| C4878T | ORF1ab:T1538I | 3 |
| C7851T | ORF1ab:A2529V | 3 |
| G569A | ORF1ab:E102K | 3 |
| G9479T | ORF1ab:G3072C | 3 |

**Table S8.** Amino acid substitutions in the viral genome associated with resistance to monoclonal antibodies, molnupiravir, and nirmatrelvir/ritonavir, and their correlation with immunosuppression diagnosis.

| Amino Acid Substitution | Immunosuppression Diagnosis | Proportion with Immunosuppression |
| --- | --- | --- |
| S:E340K | 14/15 | 93.3% |
| S:R346T | 5/7 | 71.4% |
| S:K444R | 3/5 | 60.0% |
| S:R346I | 3/3 | 100% |
| S:K356T | 2/3 | 66.7% |
| S:S494P | 1/2 | 50.0% |
| S:K440R | 3/3 | 100% |
| S:P9L | 3/3 | 100% |
| ORF1ab:L3829F | 4/7 | 57.1% |

**Table S9.**
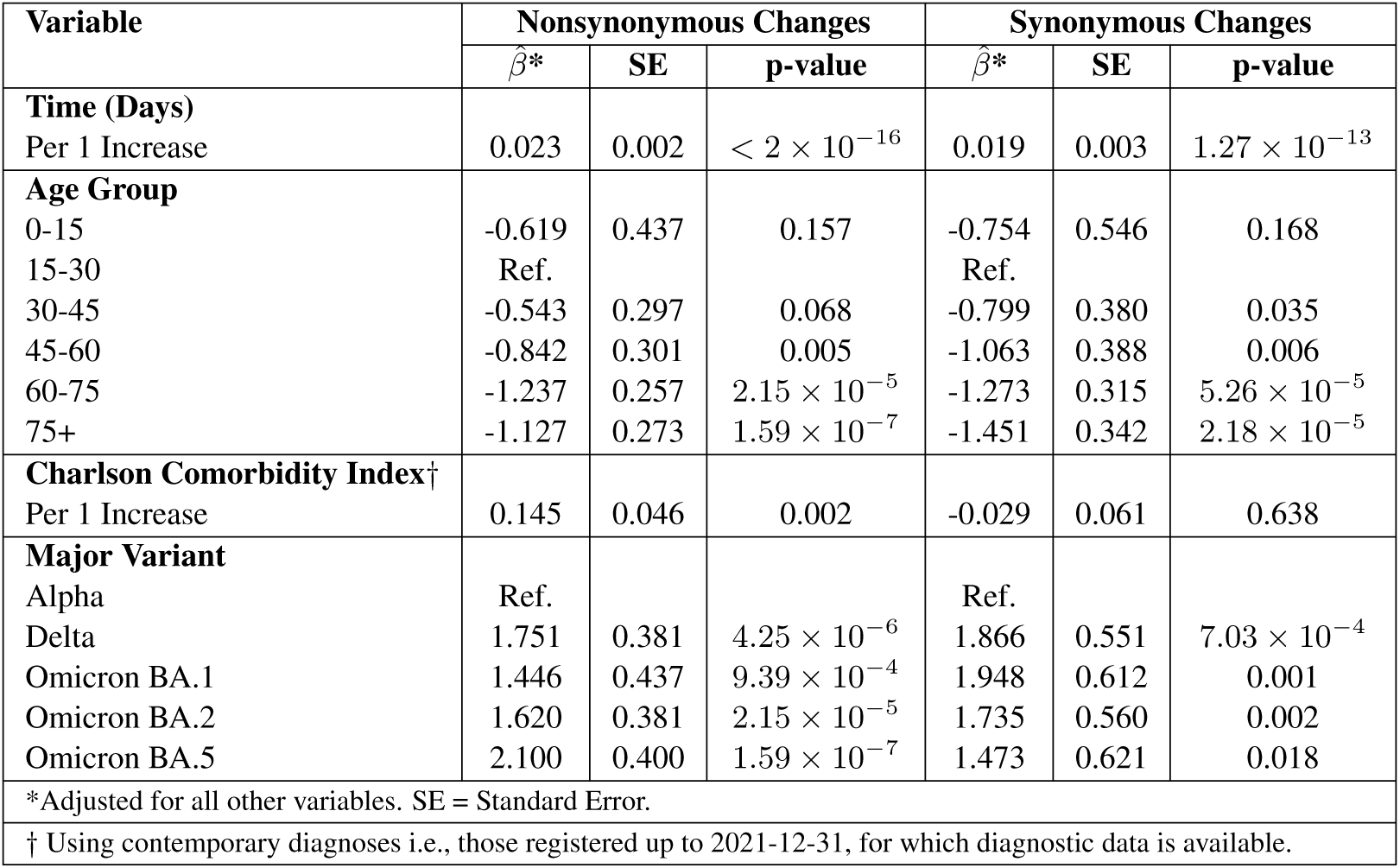
Regression results for nonsynonymous and synonymous changes. Estimates (*β̂*), standard errors (SE), and p-values are from a zero-inflated negative binomial regression model, adjusted for all other variables. Results are shown separately for nonsynonymous and synonymous changes. The Charlson Comorbidity Index was calculated using contemporary diagnoses (up to December 31, 2021). Estimates denote the absolute number of (non)synonymous changes across all coding regions relative to the reference group.

**Table S10.** Regression Results for Nonsynonymous and Synonymous Changes by Diagnostic Group. Coefficients (*β̂*), standard errors (SE), and p-values are from a zero-inflated negative binomial regression model, adjusted for time between samples, major variant, and age group. Diagnoses are based on pre-pandemic data (before January 1, 2020).

| Diagnosis Group<sup>†</sup> | Nonsynonymous Changes |  |  | Synonymous Changes |  |  |
| --- | --- | --- | --- | --- | --- | --- |
| | $\hat{\beta}^*$ | SE | p-value | $\hat{\beta}^*$ | SE | p-value |
| **Model 1: Immunosuppression** | 0.458 | 0.105 | $1.39 \times 10^{-5}$ | 0.082 | 0.167 | 0.625 |
| **Model 2: Autoimmune** | 0.138 | 0.216 | 0.523 | -0.291 | 0.365 | 0.426 |
| **Model 3: Diabetes** | -0.119 | 0.182 | 0.511 | -0.365 | 0.327 | 0.264 |
| **Model 4: IMID** | -0.370 | 0.254 | 0.146 | -0.060 | 0.345 | 0.861 |
| *Adjusted for time between samples, major variant and age group. IMID = Immune-mediated inflammatory diseases. SE = Standard Error. |  |  |  |  |  |  |
| <sup>†</sup> Using pre-pandemic diagnoses prior to 2020-01-01. |  |  |  |  |  |  |

**Table S11.** Regression Results for Nonsynonymous and Synonymous Changes. Coefficients (*β̂*), standard errors (SE), and p-values are from a zero-inflated negative binomial regression model, adjusted for time between samples, major variant, and age group. The Charlson Comorbidity Index is based on a 5-year lookback from 2020-01-01.

| Variable | Nonsynonymous Changes |  |  | Synonymous Changes |  |  |
| --- | --- | --- | --- | --- | --- | --- |
| | $\hat{\beta}^*$ | SE | p-value | $\hat{\beta}^*$ | SE | p-value |
| **Time (Days)**<br>Per 1 Increase | 0.023 | 0.002 | $< 2 \times 10^{-16}$ | 0.019 | 0.002 | $3.22 \times 10^{-14}$ |
| **Age Group** |  |  |  |  |  |  |
| 0-15 | -0.611 | 0.442 | 0.167 | -0.682 | 0.542 | 0.208 |
| 15-30 | Ref. |  |  | Ref. |  |  |
| 30-45 | -0.499 | 0.297 | 0.093 | -0.746 | 0.381 | 0.050 |
| 45-60 | -0.781 | 0.298 | 0.009 | -1.052 | 0.388 | 0.007 |
| 60-75 | -0.610 | 0.254 | 0.016 | -1.270 | 0.317 | $6.05 \times 10^{-5}$ |
| 75+ | -1.154 | 0.268 | $1.70 \times 10^{-5}$ | -1.466 | 0.343 | $1.96 \times 10^{-5}$ |
| **Charlson Comorbidity Index†**<br>Per 1 Increase | 0.094 | 0.043 | 0.029 | -0.030 | 0.062 | 0.627 |
| **Major Variant** |  |  |  |  |  |  |
| Alpha | Ref. |  |  | Ref. |  |  |
| Delta | 1.753 | 0.381 | $4.08 \times 10^{-6}$ | 1.857 | 0.550 | $7.30 \times 10^{-4}$ |
| Omicron BA.1 | 1.507 | 0.434 | $5.18 \times 10^{-4}$ | 1.922 | 0.611 | 0.002 |
| Omicron BA.2 | 1.616 | 0.381 | $2.28 \times 10^{-5}$ | 1.681 | 0.560 | 0.003 |
| Omicron BA.5 | 2.183 | 0.398 | $4.02 \times 10^{-8}$ | 1.421 | 0.617 | 0.021 |
| *Adjusted for all other variables; † 5-year lookback from 2020-01-01; SE = Standard Error |  |  |  |  |  |  |

**Table S12.** Regression results for nonsynonymous and synonymous changes. Coefficients (*β̂*), standard errors (SE), and p-values are from a zero-inflated negative binomial regression model, adjusted for time between samples, major variant, age group, and number of vaccinations. The Charlson Comorbidity Index is based on a 5-year lookback from 2020-01-01. SE = Standard Error.

| Variable | Nonsynonymous Changes |  |  | Synonymous Changes |  |  |
| --- | --- | --- | --- | --- | --- | --- |
| | $\hat{\beta}^*$ | SE | p-value | $\hat{\beta}^*$ | SE | p-value |
| **Time (Days)**<br>Per 1 Increase | 0.023 | 0.002 | $< 2 \times 10^{-16}$ | 0.019 | 0.002 | $3.22 \times 10^{-14}$ |
| **Age Group** |  |  |  |  |  |  |
| 0-15 | -0.573 | 0.442 | 0.195 | -0.699 | 0.544 | 0.199 |
| 15-30 | Ref. |  |  | Ref. |  |  |
| 30-45 | -0.540 | 0.301 | 0.073 | -0.725 | 0.385 | 0.060 |
| 45-60 | -0.879 | 0.318 | 0.006 | -1.014 | 0.400 | 0.011 |
| 60-75 | -0.733 | 0.289 | 0.011 | -1.215 | 0.349 | 0.00049 |
| 75+ | -1.282 | 0.305 | $2.57 \times 10^{-5}$ | -1.408 | 0.376 | $1.79 \times 10^{-4}$ |
| **Charlson Comorbidity Index†**<br>Per 1 Increase | 0.086 | 0.044 | 0.053 | 0.037 | 0.065 | 0.567 |
| **Major Variant** |  |  |  |  |  |  |
| Alpha | Ref. |  |  | Ref. |  |  |
| Delta | 1.696 | 0.386 | $1.14 \times 10^{-5}$ | 1.885 | 0.555 | $6.74 \times 10^{-4}$ |
| Omicron BA.1 | 1.393 | 0.454 | 0.002 | 1.983 | 0.631 | 0.002 |
| Omicron BA.2 | 1.489 | 0.408 | $2.61 \times 10^{-4}$ | 1.753 | 0.591 | 0.003 |
| Omicron BA.5 | 2.038 | 0.430 | $2.13 \times 10^{-6}$ | 1.505 | 0.656 | 0.022 |
| **Number of vaccinations**<br>Per 1 Increase | 0.083 | 0.093 | 0.370 | -0.045 | 0.121 | 0.707 |
| *Adjusted for all other variables; † 5-year lookback from 2020-01-01; SE = Standard Error |  |  |  |  |  |  |

**Table S13.** Quasi-Poisson regression results for rate per site, using the first and last sequences for each person to calculate the rate. Among those with at least one consensus-level change, the mean rate was 1.03 *×* 10^−3^ substitutions per site per year (95% CI: 2.50 *×* 10^−4^, 2.78 *×* 10^−3^).

| Variable | Rate Ratio (RR) | 95% CI | p-value |
| --- | --- | --- | --- |
| **Time (Days)** |  |  |  |
| Per 1 Increase | 1.009 | (1.004, 1.013) | $1.95 \times 10^{-4}$ |
| **Sex** |  |  |  |
| Female | 0.916 | (0.611, 1.373) | 0.671 |
| Male | Ref. | – | – |
| **Number of Vaccinations** |  |  |  |
| Per 1 Increase | 1.166 | (0.955, 1.424) | 0.132 |
| **Age Group** |  |  |  |
| 0-15 | 0.482 | (0.132, 1.756) | 0.269 |
| 15-30 | Ref. | – | – |
| 30-45 | 0.501 | (0.226, 1.109) | 0.089 |
| 45-60 | 0.359 | (0.164, 0.786) | 0.011 |
| 60-75 | 0.451 | (0.233, 0.873) | 0.019 |
| 75+ | 0.311 | (0.151, 0.639) | 0.002 |
| All estimates are from a quasi-Poisson regression model. |  |  |  |

**Table S14.** Summary of Ka-Ks Estimates with Credible Intervals for Persistent Infections.

| Coding Region | Estimate | Lower 95% CrI | Upper 95% CrI |
| --- | --- | --- | --- |
| ORF1a | $2.78 \times 10^{-5}$ | $-3.68 \times 10^{-5}$ | $9.25 \times 10^{-5}$ |
| ORF1b | $-6.91 \times 10^{-5}$ | $-1.87 \times 10^{-4}$ | $5.14 \times 10^{-5}$ |
| S | $4.12 \times 10^{-4}$ | $2.66 \times 10^{-4}$ | $5.56 \times 10^{-4}$ |
| ORF3a | $-1.60 \times 10^{-4}$ | $-1.88 \times 10^{-3}$ | $1.59 \times 10^{-3}$ |
| E | $6.59 \times 10^{-3}$ | $5.87 \times 10^{-3}$ | $7.28 \times 10^{-3}$ |
| M | $-1.28 \times 10^{-3}$ | $-3.96 \times 10^{-3}$ | $1.26 \times 10^{-3}$ |
| ORF6 | $-7.78 \times 10^{-4}$ | $-1.75 \times 10^{-2}$ | $1.74 \times 10^{-2}$ |
| ORF7a | $3.58 \times 10^{-3}$ | $5.27 \times 10^{-4}$ | $6.60 \times 10^{-3}$ |
| ORF7b | $9.98 \times 10^{-3}$ | $-8.25 \times 10^{-3}$ | $2.04 \times 10^{-2}$ |
| ORF8 | $-6.72 \times 10^{-4}$ | $-1.01 \times 10^{-2}$ | $8.96 \times 10^{-3}$ |
| N | $-2.52 \times 10^{-4}$ | $-8.53 \times 10^{-4}$ | $3.62 \times 10^{-4}$ |
| ORF10 | $-3.30 \times 10^{-3}$ | $-2.09 \times 10^{-2}$ | $1.51 \times 10^{-2}$ |

**Table S15.** Summary of Ka-Ks Estimates with Credible Intervals for Controls.

| Coding Region | Estimate | Lower 95% CrI | Upper 95% CrI |
| --- | --- | --- | --- |
| ORF1a | $-1.59 \times 10^{-4}$ | $-2.32 \times 10^{-4}$ | $-8.75 \times 10^{-5}$ |
| ORF1b | $-4.32 \times 10^{-4}$ | $-5.64 \times 10^{-4}$ | $-3.03 \times 10^{-4}$ |
| S | $2.43 \times 10^{-4}$ | $-1.79 \times 10^{-4}$ | $6.74 \times 10^{-4}$ |
| ORF3a | $4.24 \times 10^{-4}$ | $-6.06 \times 10^{-4}$ | $1.48 \times 10^{-3}$ |
| E | $6.99 \times 10^{-3}$ | $6.99 \times 10^{-3}$ | $6.99 \times 10^{-3}$ |
| M | $-1.69 \times 10^{-3}$ | $-3.41 \times 10^{-3}$ | $8.14 \times 10^{-5}$ |
| ORF6 | $-9.70 \times 10^{-3}$ | $-1.55 \times 10^{-2}$ | $-3.22 \times 10^{-3}$ |
| ORF7a | $1.80 \times 10^{-3}$ | $-3.40 \times 10^{-3}$ | $7.14 \times 10^{-3}$ |
| ORF7b | $-8.21 \times 10^{-4}$ | $-1.13 \times 10^{-2}$ | $9.75 \times 10^{-3}$ |
| ORF8 | $2.86 \times 10^{-3}$ | $-3.03 \times 10^{-4}$ | $5.84 \times 10^{-3}$ |
| N | $-8.28 \times 10^{-5}$ | $-9.64 \times 10^{-4}$ | $7.77 \times 10^{-4}$ |
| ORF10 | $1.22 \times 10^{-2}$ | $1.05 \times 10^{-2}$ | $1.38 \times 10^{-2}$ |

**Table S16.** Bayesian logistic regression model results without adjusting for infection duration. The table includes the estimate, standard error, and 95% credible intervals (CrI) for each parameter.

| Parameter | Estimate | Est. Error | 95% CrI |
| --- | --- | --- | --- |
| Intercept | -1.59 | 0.31 | [-2.19, -0.96] |
| Immunosuppression Diagnosis ($\beta$) | -0.28 | 0.22 | [-0.72, 0.15] |

**Table S17.** Bayesian logistic regression model results adjusting for infection duration. The table includes the estimate, standard error, and 95% credible intervals (CrI) for each parameter.

| Parameter | Estimate | Est. Error | 95% CrI |
| --- | --- | --- | --- |
| Intercept | -1.59 | 0.31 | [-2.19, -0.96] |
| Immunosuppression Diagnosis ($\beta$) | -0.29 | 0.23 | [-0.75, 0.15] |
| Infection Duration | 0.00 | 0.01 | [-0.01, 0.01] |

**Table S18.**
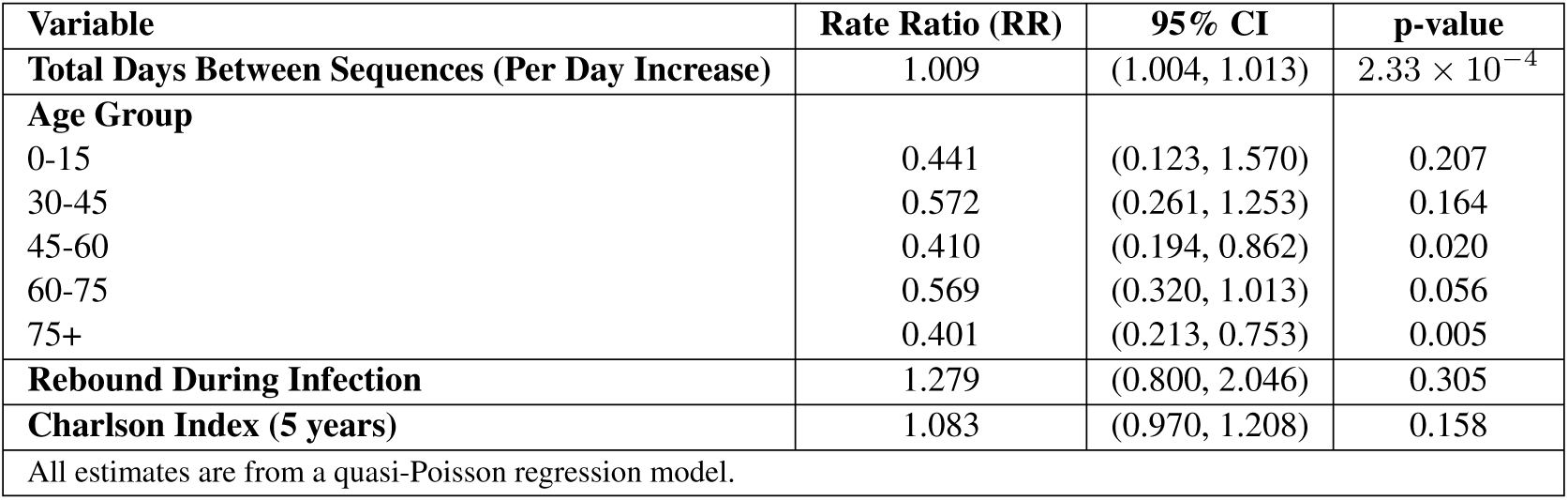
Quasi-Poisson regression results for rate per site, adjusted for total days between sequences, age group, rebound status, and Charlson index (5 year lookback).

**Table S19.**
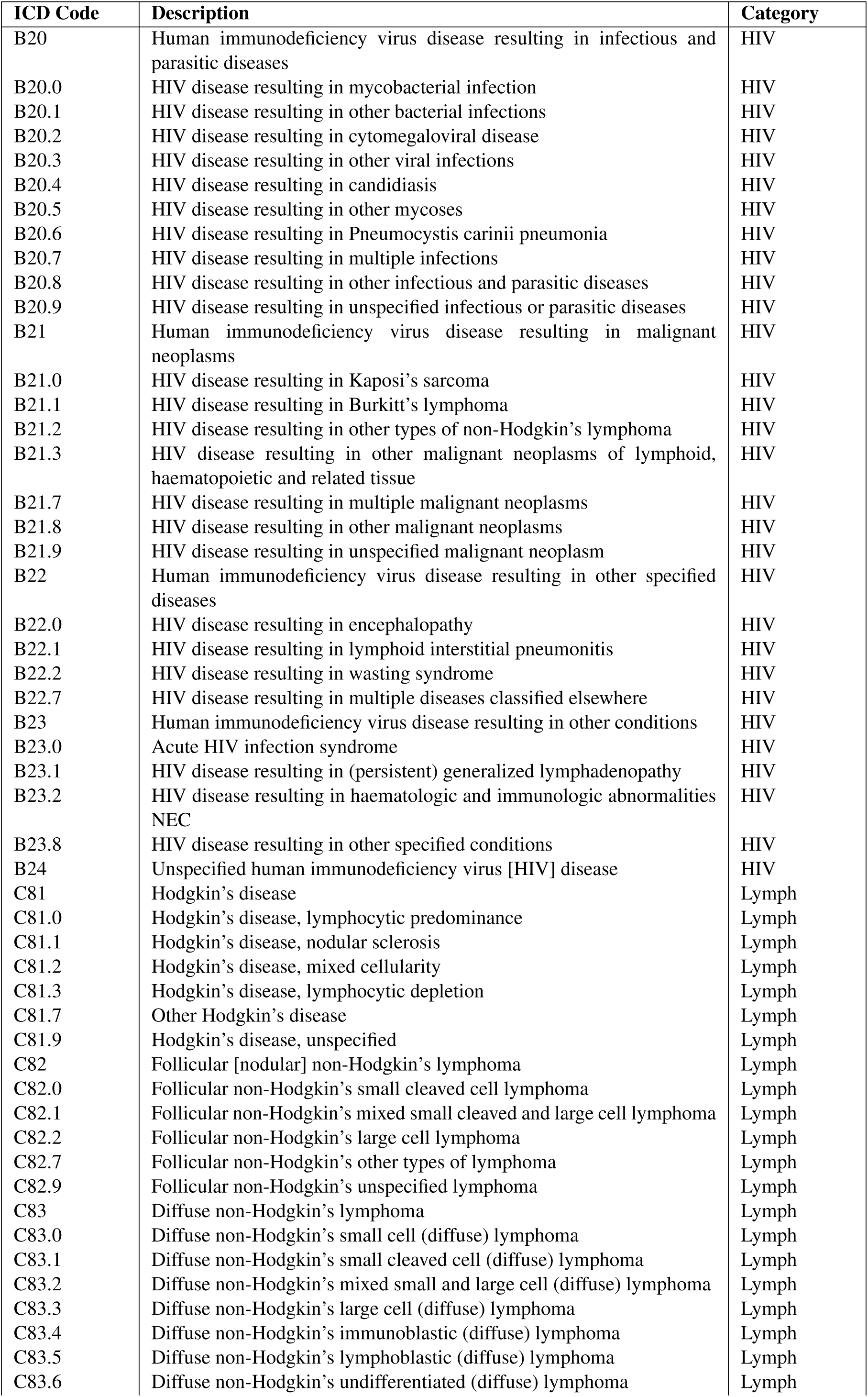
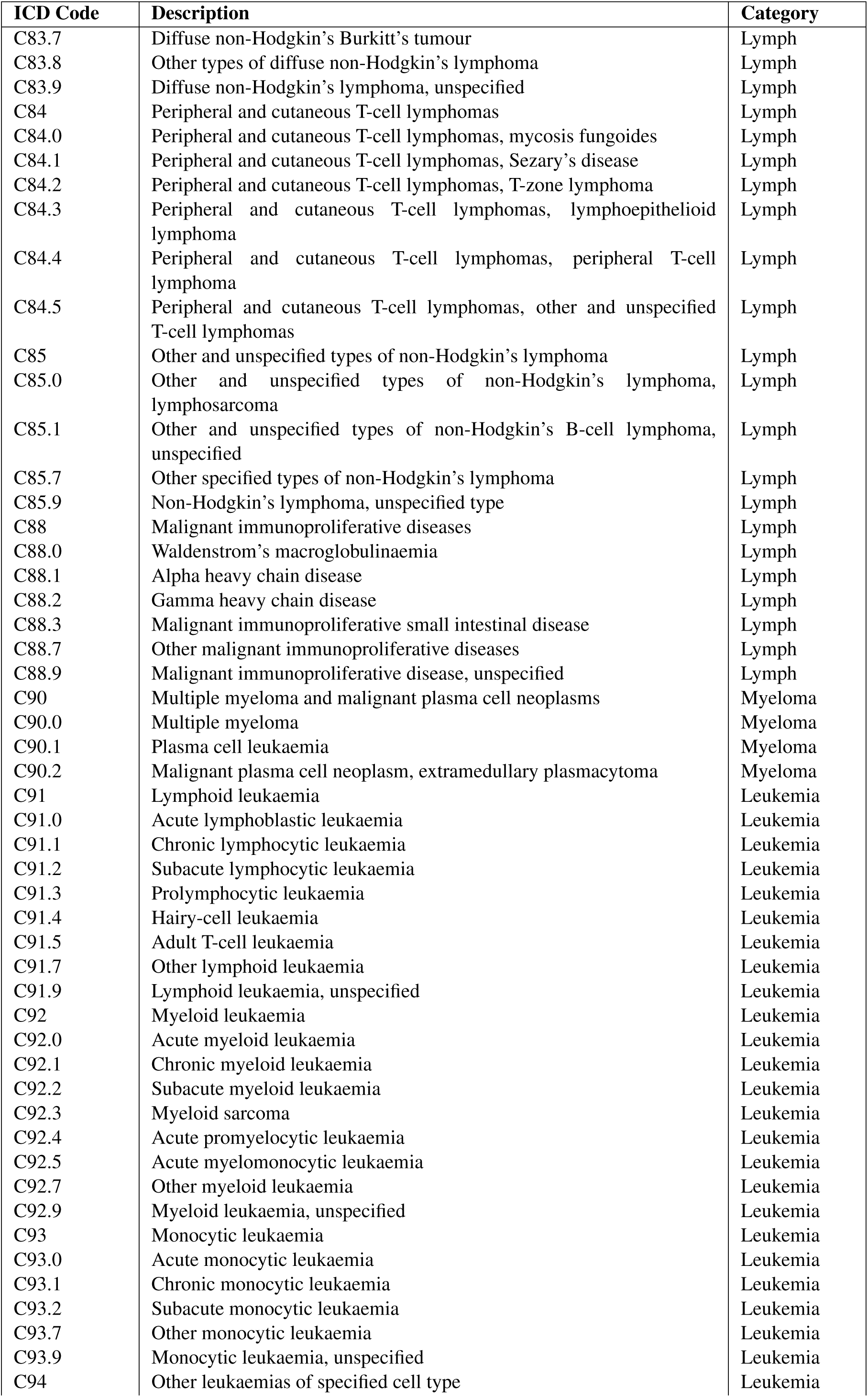
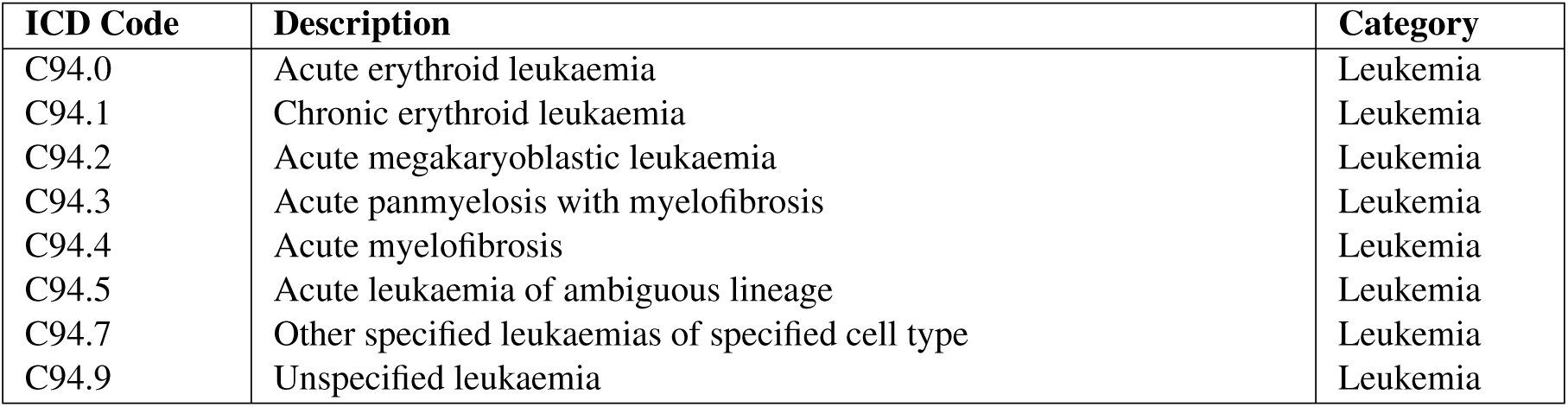
Immunosuppression Diagnosis Codes. From Mansfield et al. [81].

**Table S20.** Diabetes ICD-10 Codes. From Mansfield et al. [82].

| ICD Code | Description | Category |
| --- | --- | --- |
| E10 | Insulin-dependent diabetes mellitus | T1DM |
| E10.0 | Insulin-dependent diabetes mellitus with coma | T1DM |
| E10.1 | Insulin-dependent diabetes mellitus with ketoacidosis | T1DM |
| E10.2 | Insulin-dependent diabetes mellitus with renal complications | T1DM |
| E10.3 | Insulin-dependent diabetes mellitus with ophthalmic comps | T1DM |
| E10.4 | Insulin-dependent diabetes mellitus with neurological comps | T1DM |
| E10.5 | Insulin-dependent diabetes mellitus with periph circ comps | T1DM |
| E10.6 | Insulin-dependent diabetes mellitus with other spec comps | T1DM |
| E10.7 | Insulin-dependent diabetes mellitus with multiple comps | T1DM |
| E10.8 | Insulin-dependent diabetes mellitus with unspec comps | T1DM |
| E10.9 | Insulin-dependent diabetes mellitus without complications | T1DM |
| E11 | Non-insulin-dependent diabetes mellitus | T2DM |
| E11.0 | Non-insulin-dependent diabetes mellitus with coma | T2DM |
| E11.1 | Non-insulin-dependent diabetes mellitus with ketoacidosis | T2DM |
| E11.2 | Non-insulin-dependent diabetes mellitus with renal comps | T2DM |
| E11.3 | Non-insulin-dependent diabetes mellitus with ophthalm comps | T2DM |
| E11.4 | Non-insulin-dependent diabetes mellitus with neuro comps | T2DM |
| E11.5 | Non-insulin-depend diabetes mellitus with periph circ comp | T2DM |
| E11.6 | Non-insulin-depend diabetes mellitus with other spec comp | T2DM |
| E11.7 | Non-insulin-dependent diabetes mellitus with multiple comps | T2DM |
| E11.8 | Non-insulin-dependent diabetes mellitus with unspec comps | T2DM |
| E11.9 | Non-insulin-depend diabetes mellitus without complication | T2DM |
| E13 | Other specified diabetes mellitus | DM NOS |
| E13.0 | Other specified diabetes mellitus with coma | DM NOS |
| E13.1 | Other specified diabetes mellitus with ketoacidosis | DM NOS |
| E13.2 | Other specified diabetes mellitus with renal complications | DM NOS |
| E13.3 | Other specified diabetes mellitus with ophthalmic complications | DM NOS |
| E13.4 | Other specified diabetes mellitus with neurological comps | DM NOS |
| E13.5 | Other specified diabetes mellitus with periph circulatory comps | DM NOS |
| E13.6 | Other specified diabetes mellitus with other specified comps | DM NOS |
| E13.7 | Other specified diabetes mellitus with multiple complications | DM NOS |
| E13.8 | Other specified diabetes mellitus with unspecified comps | DM NOS |
| E13.9 | Other specified diabetes mellitus without complications | DM NOS |
| E14 | Unspecified diabetes mellitus | DM NOS |
| E14.0 | Unspecified diabetes mellitus with coma | DM NOS |
| E14.1 | Unspecified diabetes mellitus with ketoacidosis | DM NOS |
| E14.2 | Unspecified diabetes mellitus with renal complications | DM NOS |
| E14.3 | Unspecified diabetes mellitus with ophthalmic complications | DM NOS |
| E14.4 | Unspecified diabetes mellitus with neurological comps | DM NOS |
| E14.5 | Unspecified diabetes mellitus with periph circulatory comps | DM NOS |
| E14.6 | Unspecified diabetes mellitus with other specified comps | DM NOS |
| E14.7 | Unspecified diabetes mellitus with multiple complications | DM NOS |
| E14.8 | Unspecified diabetes mellitus with unspecified complications | DM NOS |
| E14.9 | Unspecified diabetes mellitus without complications | DM NOS |
| G59.0 | Diabetic mononeuropathy | DM NOS |
| G63.2 | Diabetic polyneuropathy | DM NOS |
| H28.0 | Diabetic cataract | DM NOS |
| H36.0 | Diabetic retinopathy | DM NOS |
| M14.2 | Diabetic arthropathy | DM NOS |
| N08.3 | Glomerular disorders in diabetes mellitus | DM NOS |
| O24.0 | Pre-existing diabetes mellitus, insulin-dependent | T1DM |
| O24.1 | Pre-existing diabetes mellitus, non-insulin-dependent | T2DM |
| O24.3 | Pre-existing diabetes mellitus, unspecified | DM NOS |
| Y42.3 | Insulin and oral hypoglycaemic [antidiabetic] drugs | Possible |

**Table S21.**
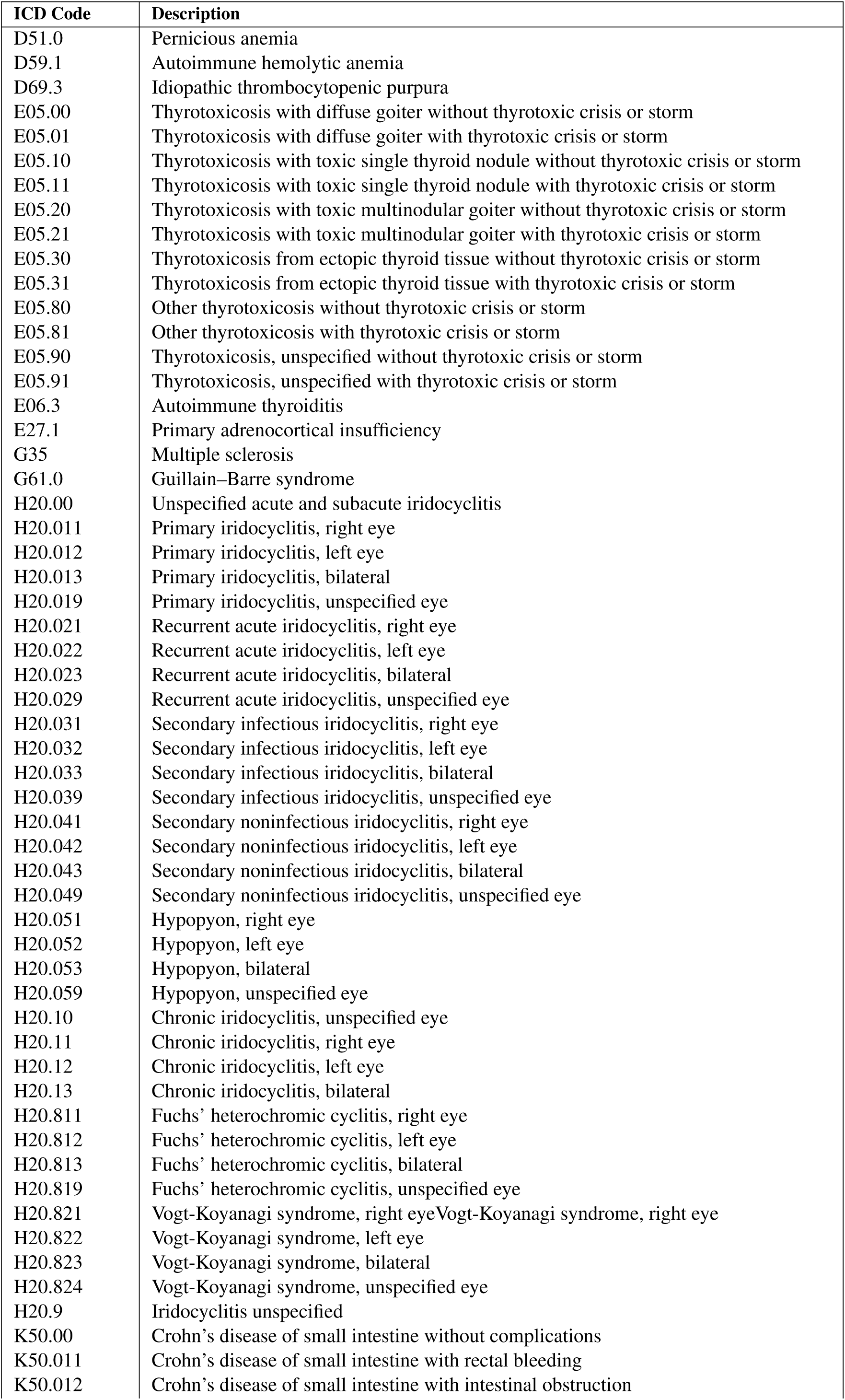
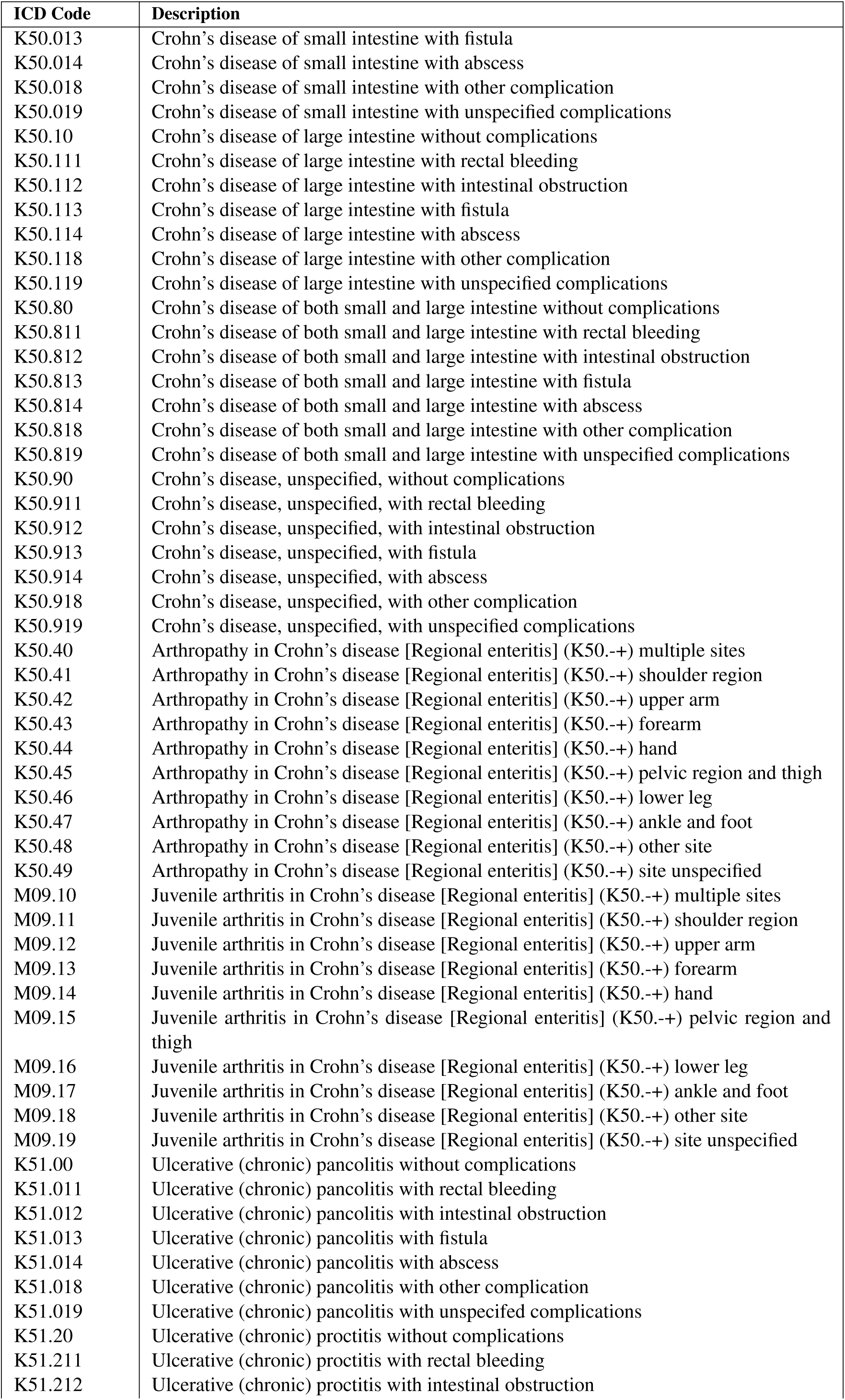
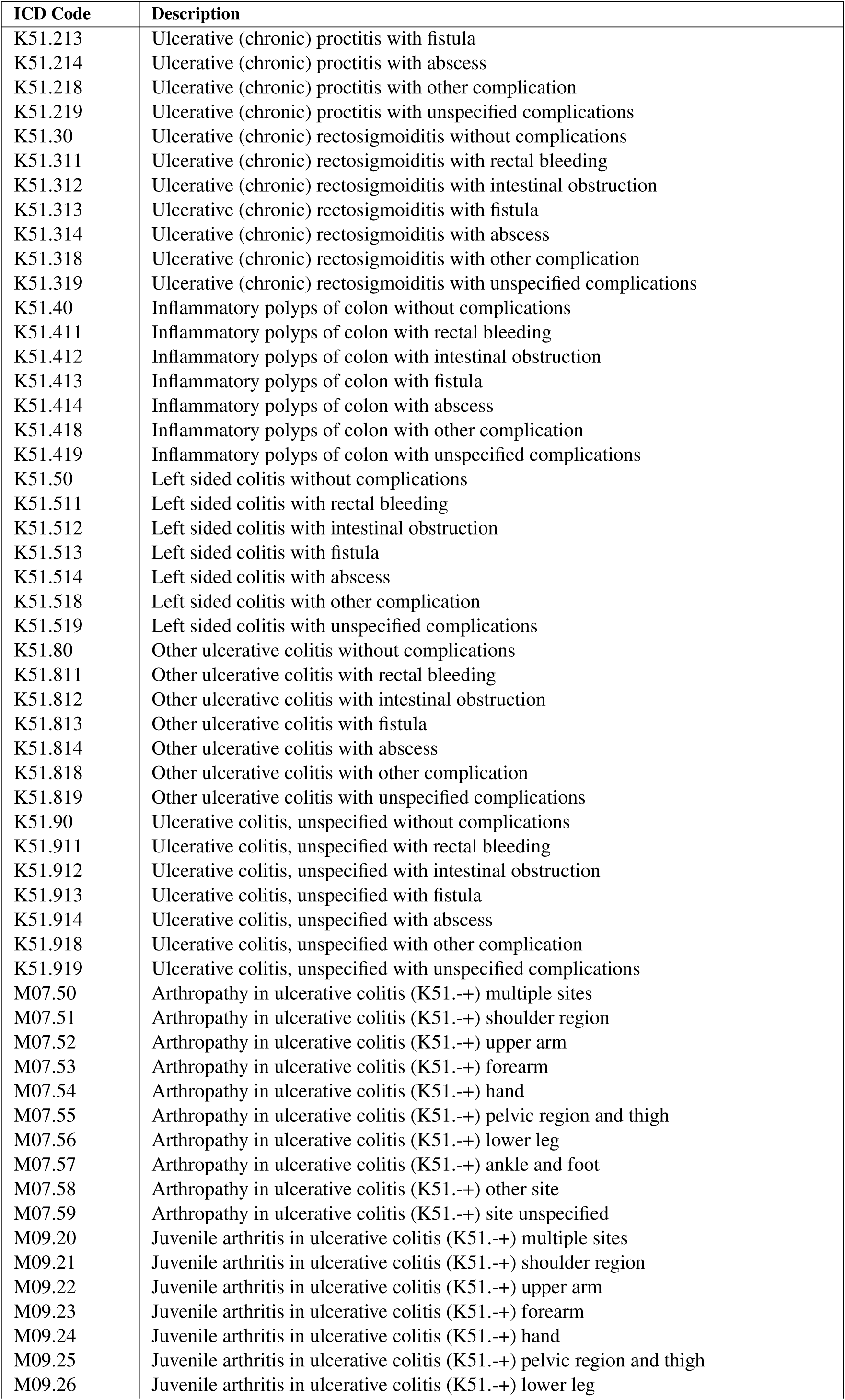
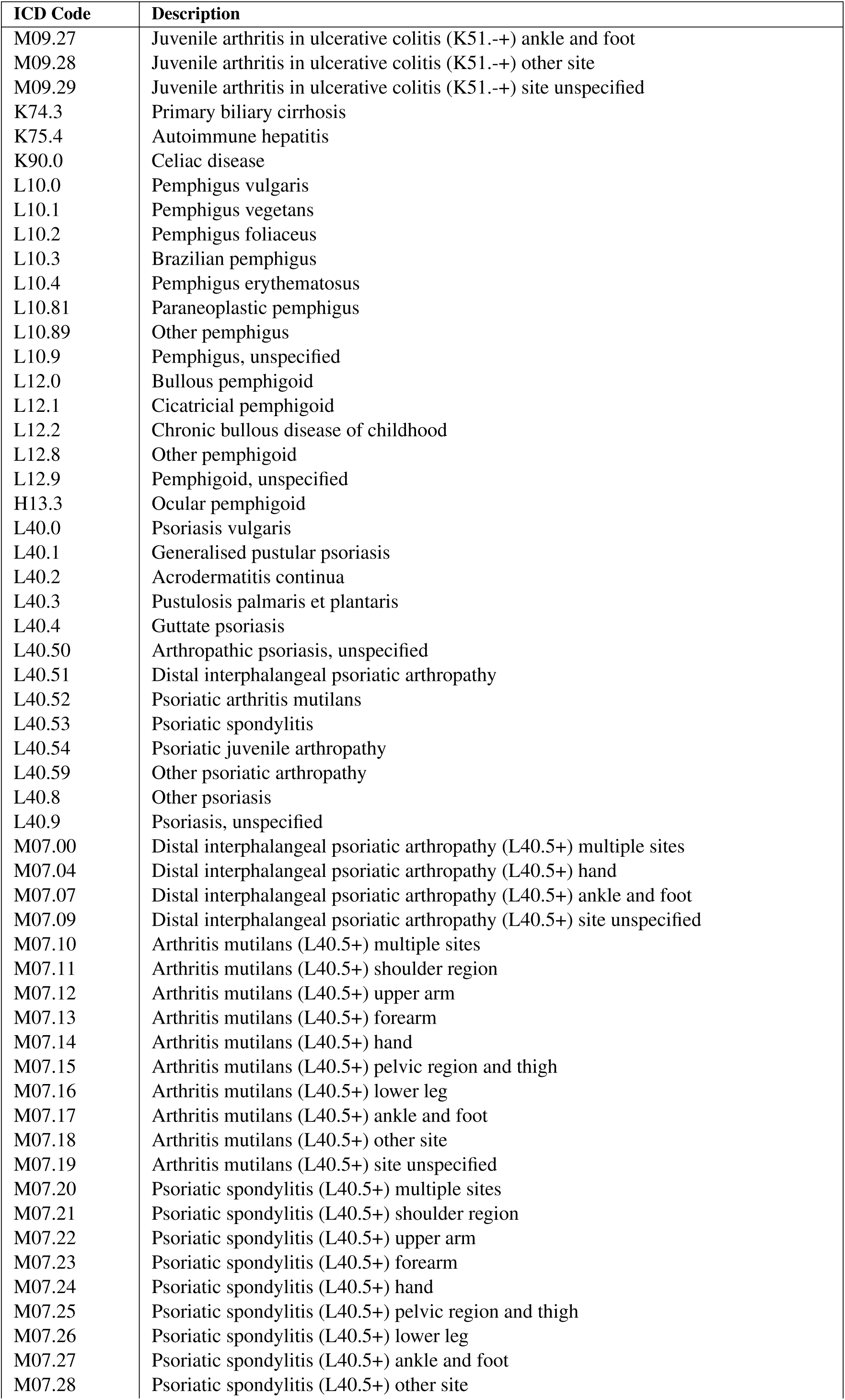
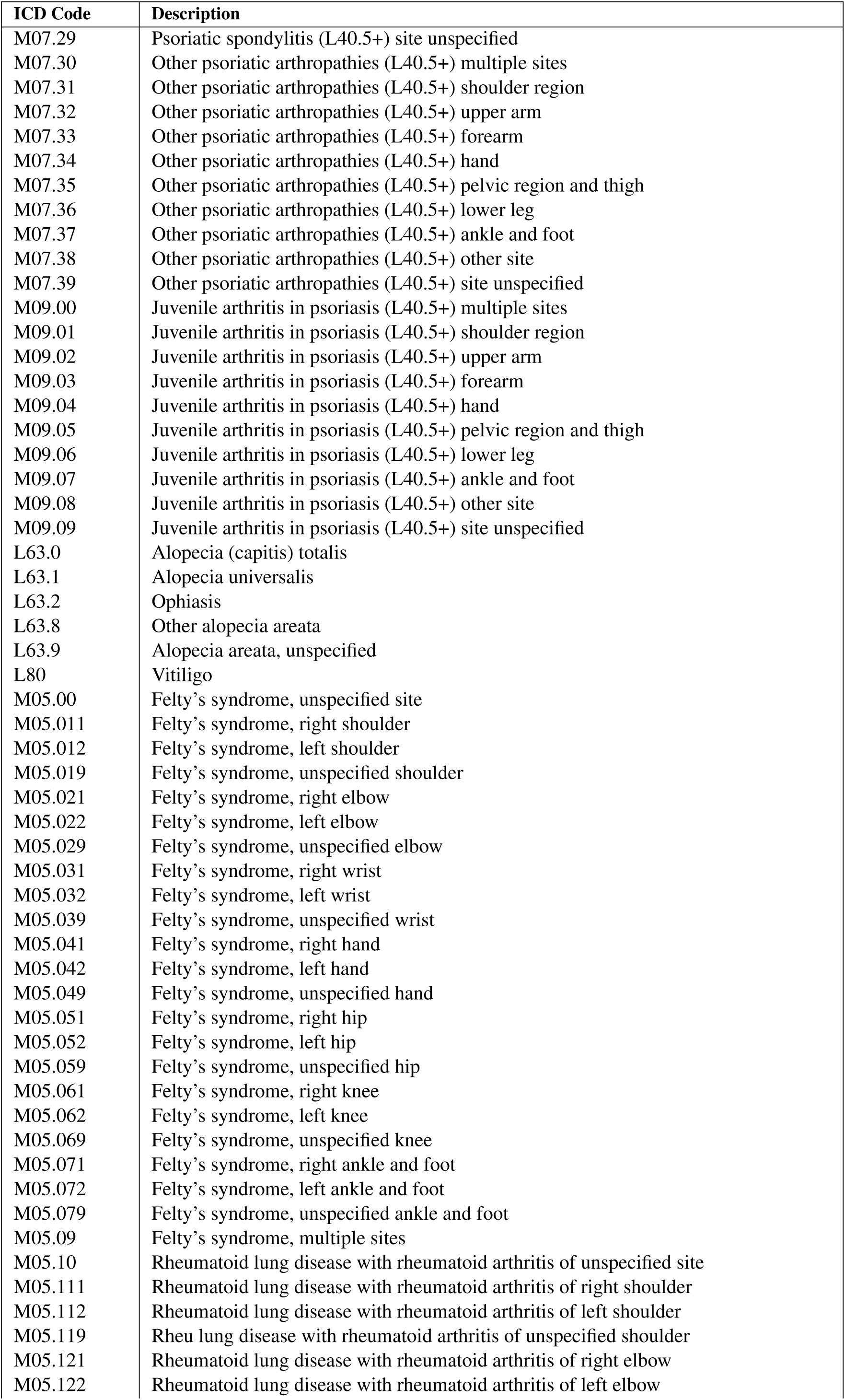
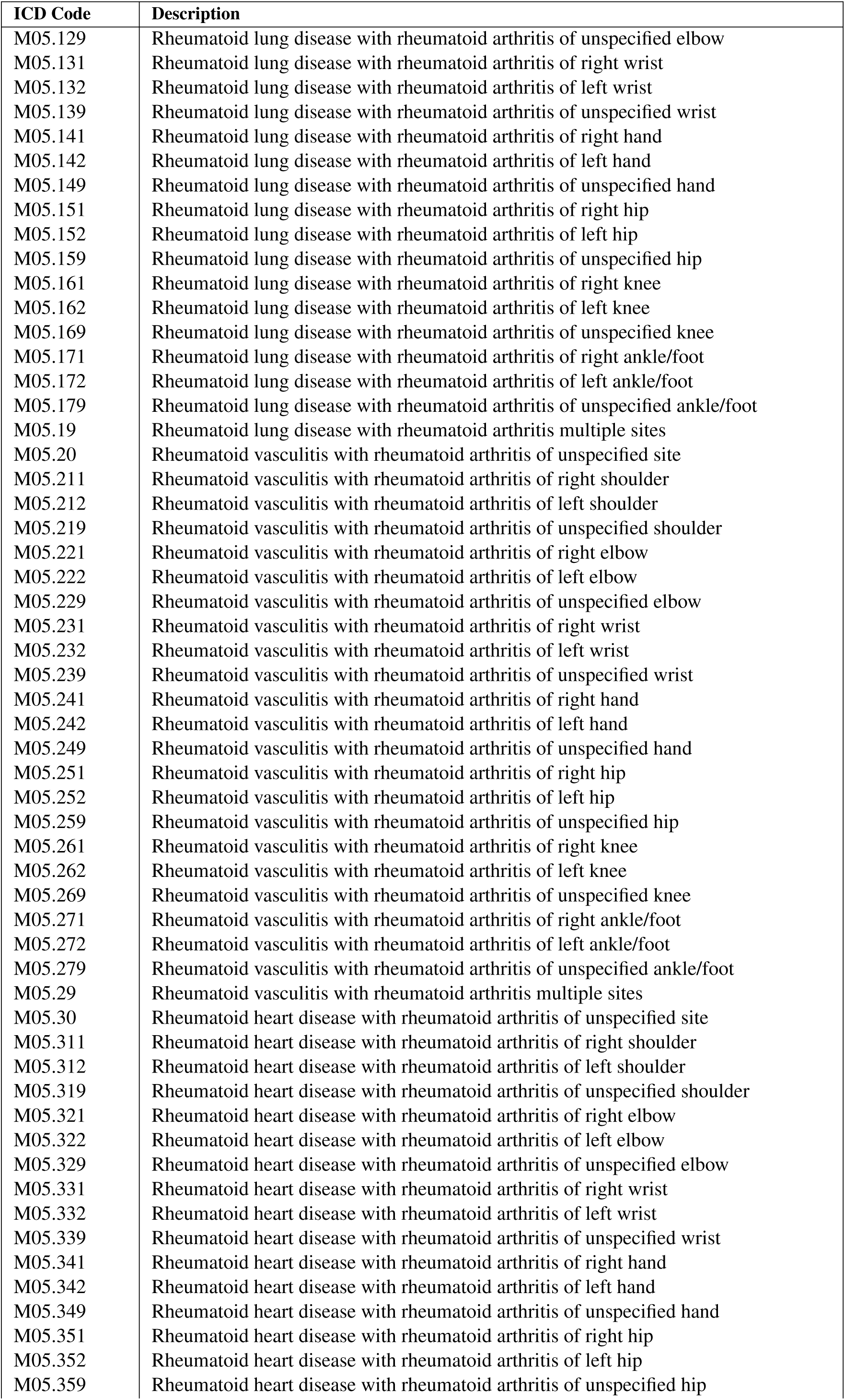
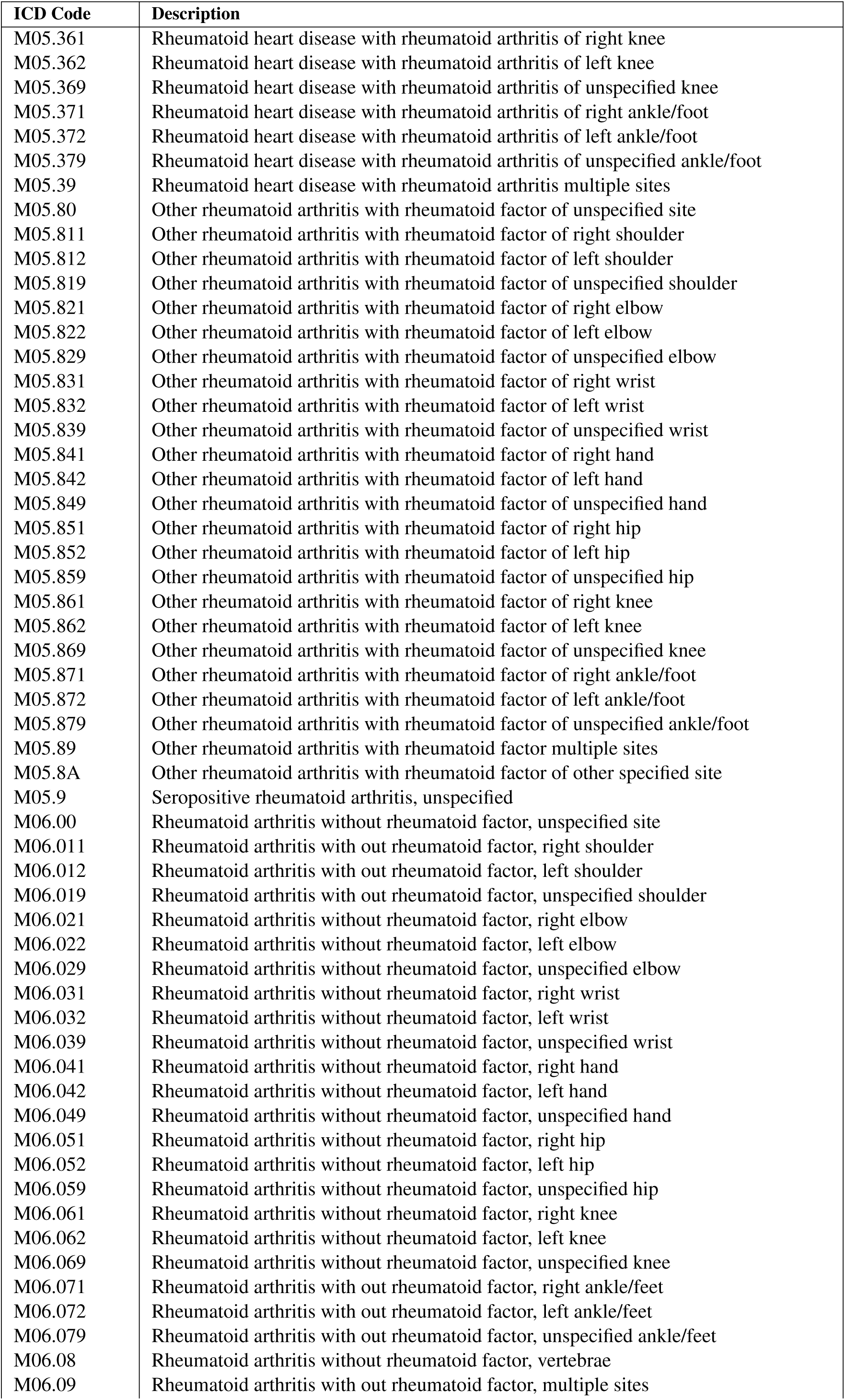
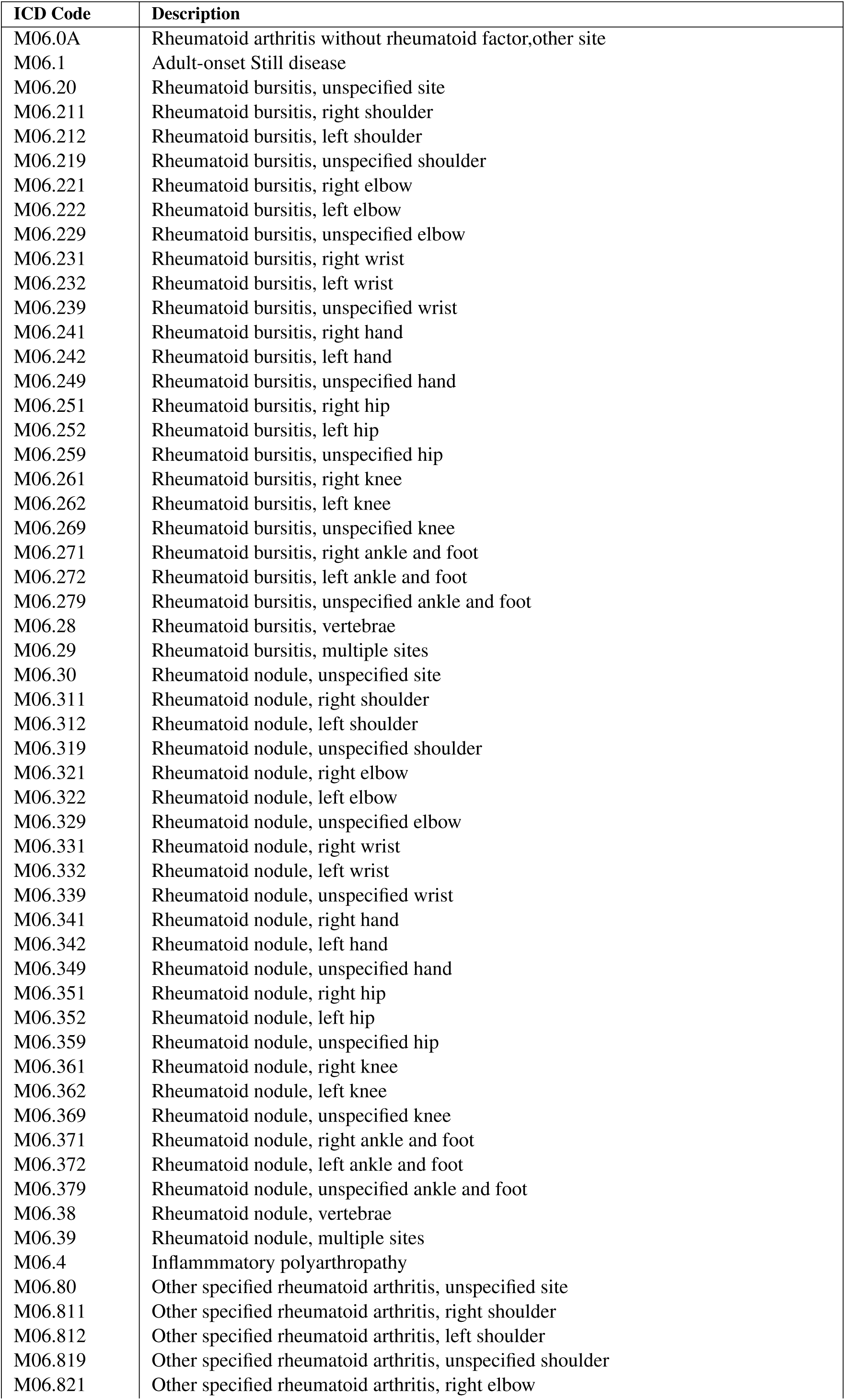
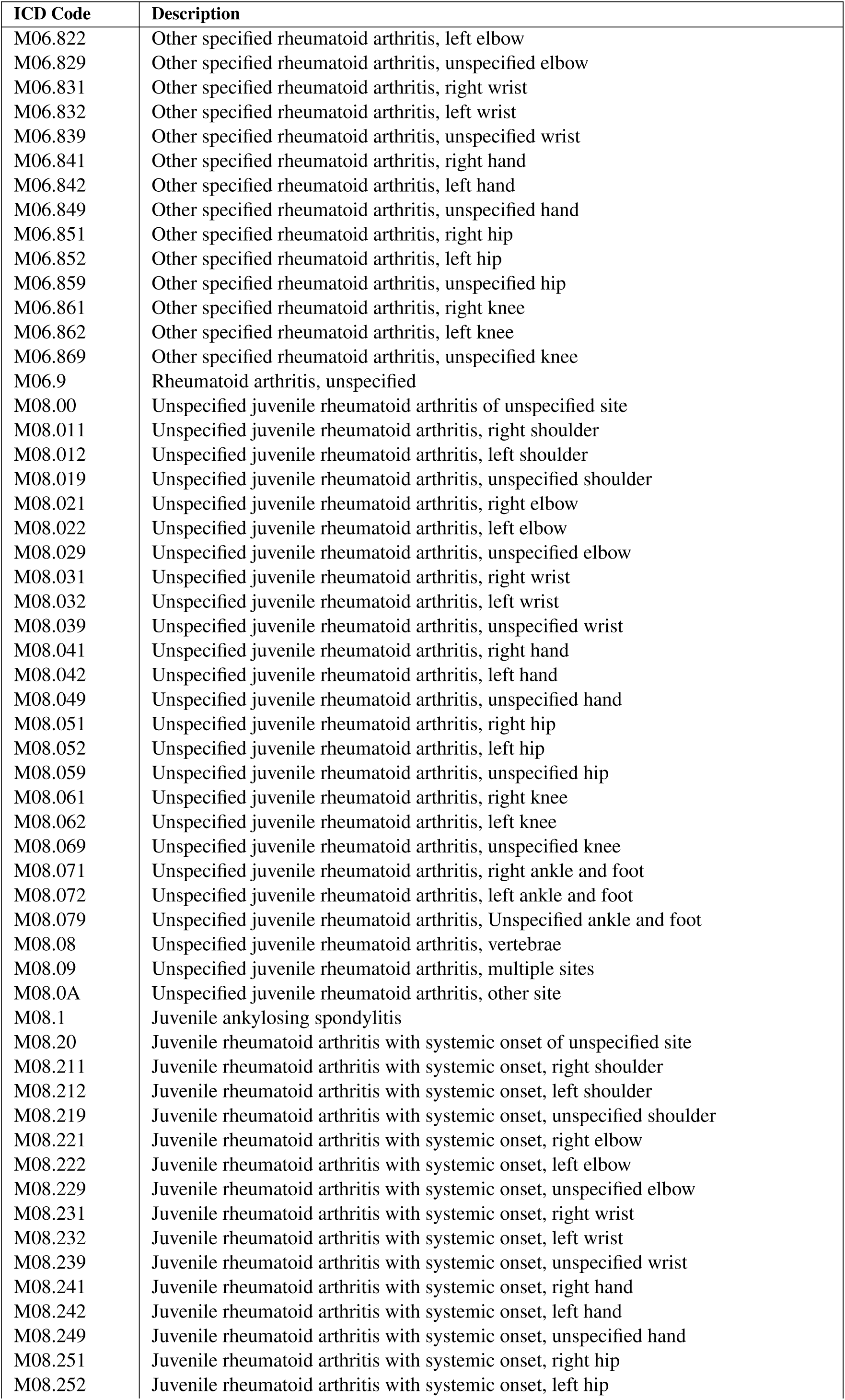
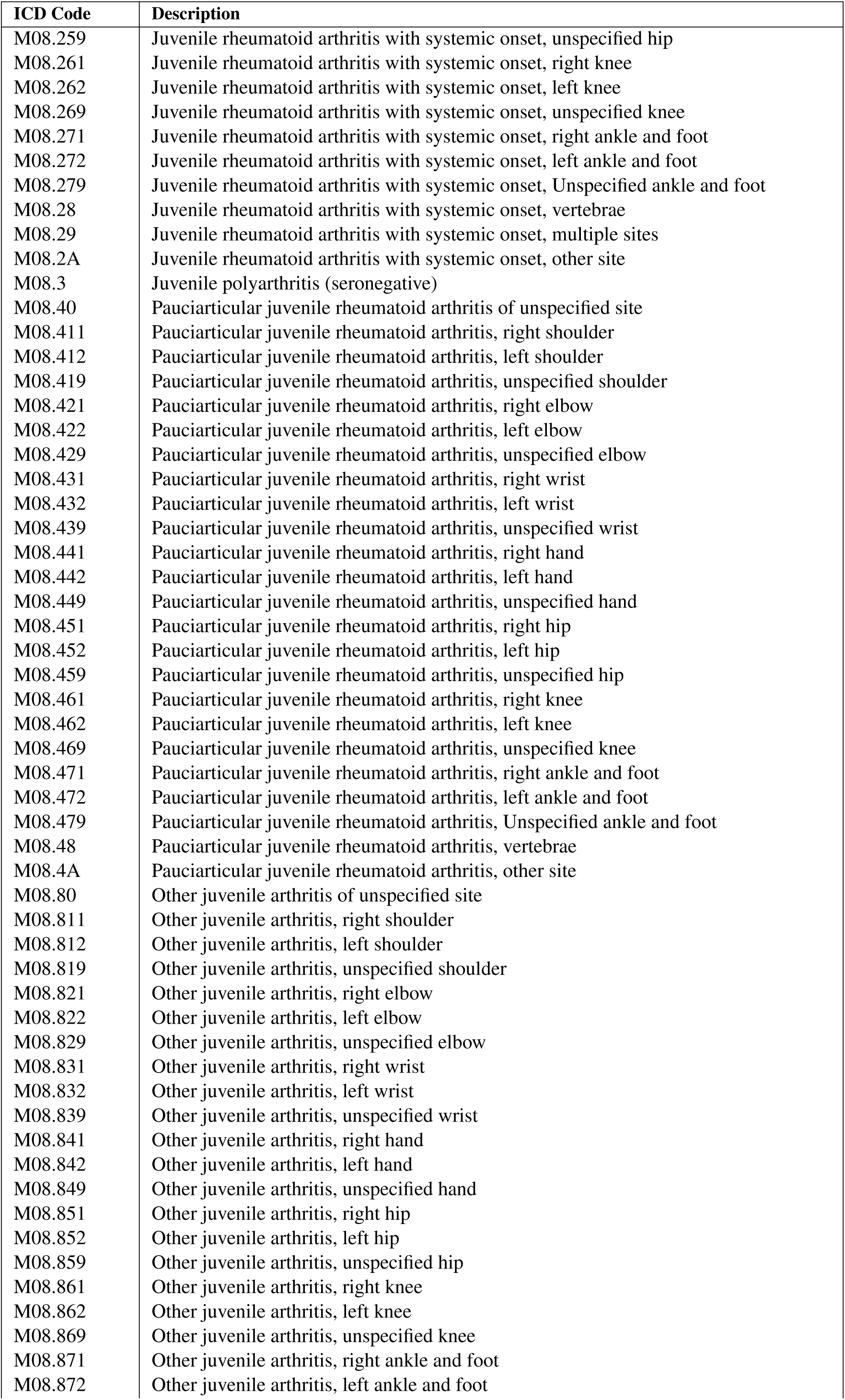
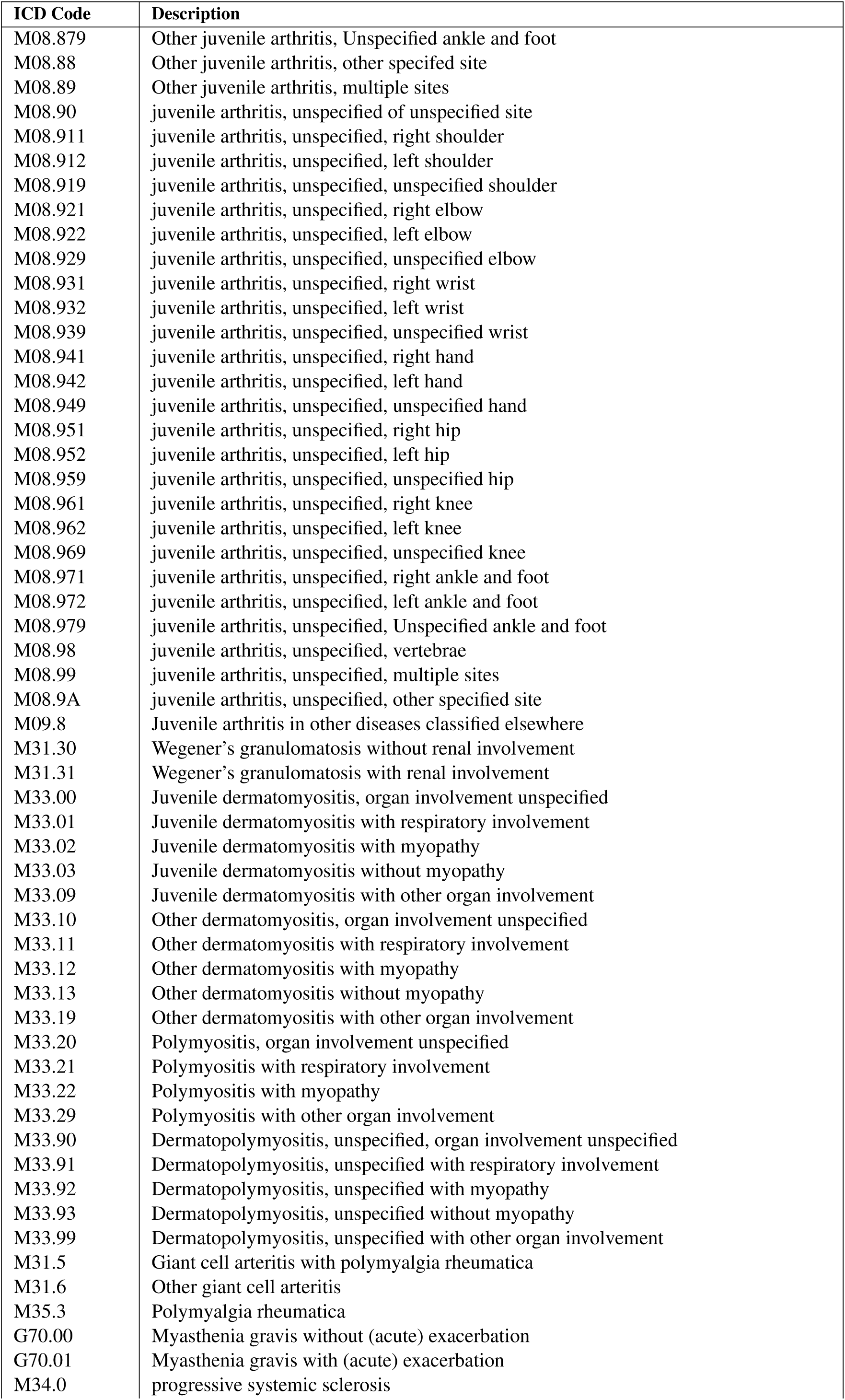
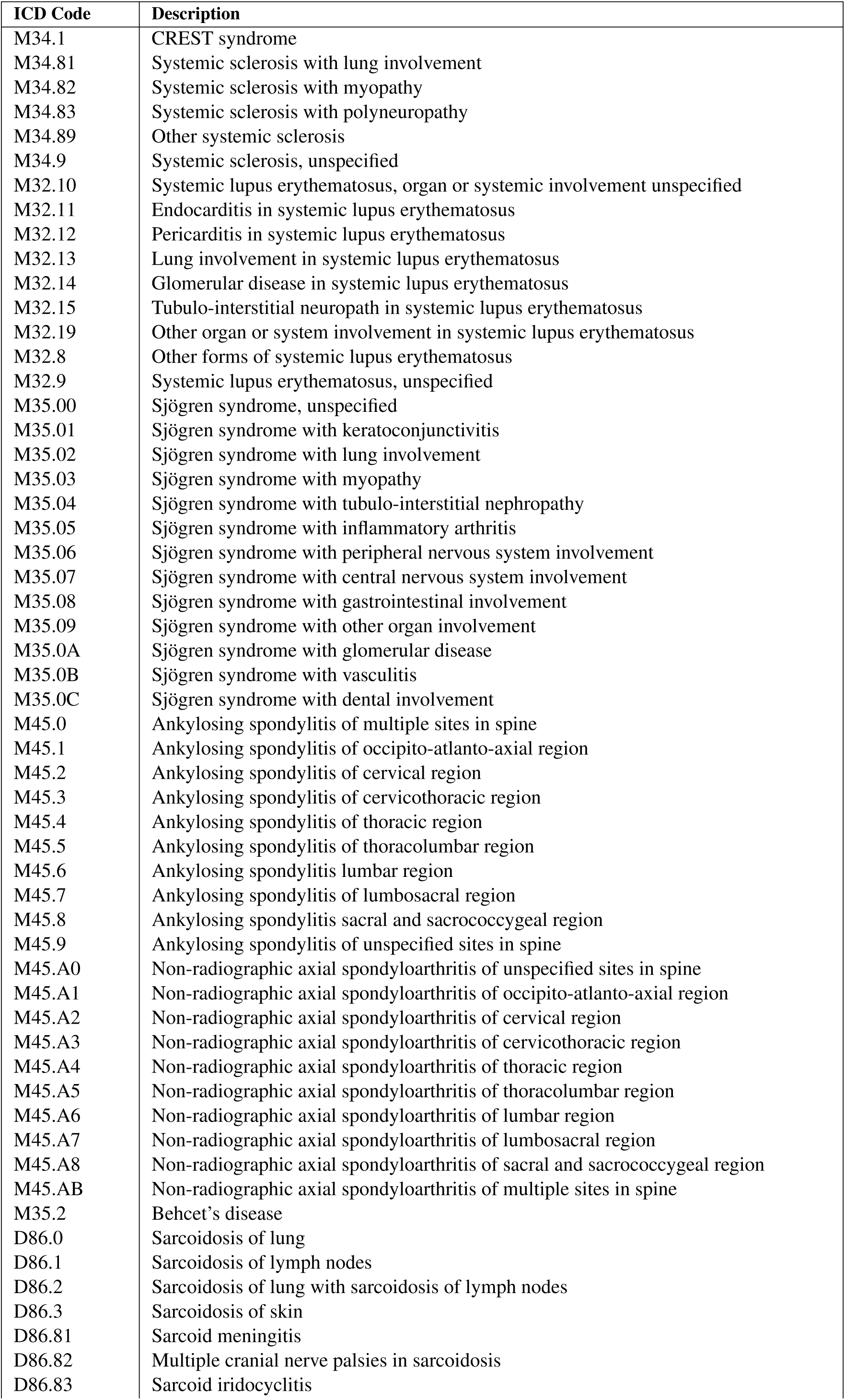
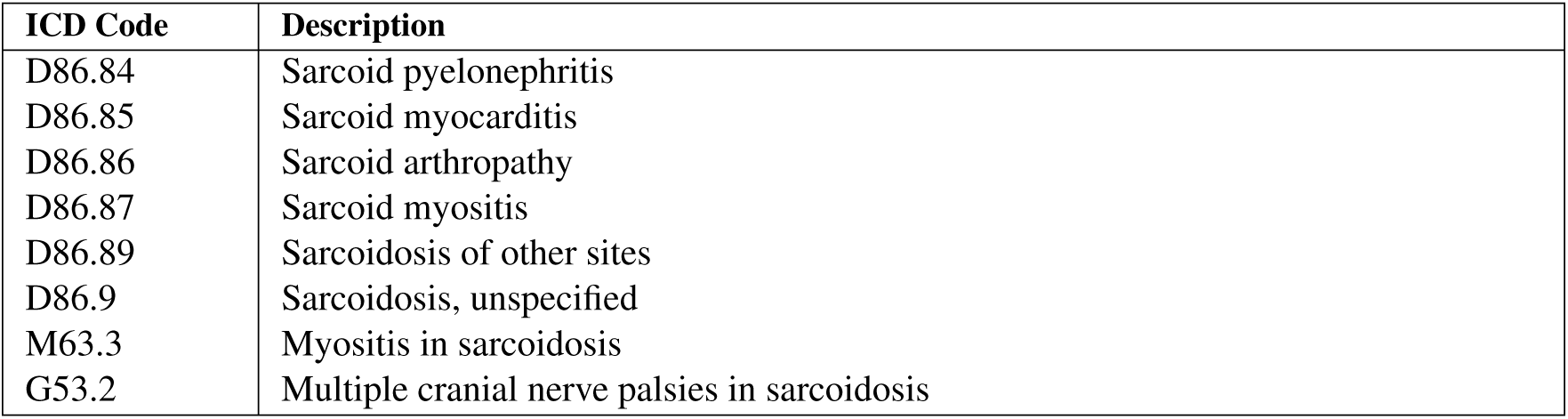
Autoimmune ICD-10 Codes. From Tunnicliffe et al. [83].

| ICD Code | Description |
| --- | --- |
| D51.0 | Pernicious anemia |
| D59.1 | Autoimmune hemolytic anemia |
| D69.3 | Idiopathic thrombocytopenic purpura |
| E05.00 | Thyrotoxicosis with diffuse goiter without thyrotoxic crisis or storm |
| E05.01 | Thyrotoxicosis with diffuse goiter with thyrotoxic crisis or storm |
| E05.10 | Thyrotoxicosis with toxic single thyroid nodule without thyrotoxic crisis or storm |
| E05.11 | Thyrotoxicosis with toxic single thyroid nodule with thyrotoxic crisis or storm |
| E05.20 | Thyrotoxicosis with toxic multinodular goiter without thyrotoxic crisis or storm |
| E05.21 | Thyrotoxicosis with toxic multinodular goiter with thyrotoxic crisis or storm |
| E05.30 | Thyrotoxicosis from ectopic thyroid tissue without thyrotoxic crisis or storm |
| E05.31 | Thyrotoxicosis from ectopic thyroid tissue with thyrotoxic crisis or storm |
| E05.80 | Other thyrotoxicosis without thyrotoxic crisis or storm |
| E05.81 | Other thyrotoxicosis with thyrotoxic crisis or storm |
| E05.90 | Thyrotoxicosis, unspecified without thyrotoxic crisis or storm |
| E05.91 | Thyrotoxicosis, unspecified with thyrotoxic crisis or storm |
| E06.3 | Autoimmune thyroiditis |
| E27.1 | Primary adrenocortical insufficiency |
| G35 | Multiple sclerosis |
| G61.0 | Guillain–Barre syndrome |
| H20.00 | Unspecified acute and subacute iridocyclitis |
| H20.011 | Primary iridocyclitis, right eye |
| H20.012 | Primary iridocyclitis, left eye |
| H20.013 | Primary iridocyclitis, bilateral |
| H20.019 | Primary iridocyclitis, unspecified eye |
| H20.021 | Recurrent acute iridocyclitis, right eye |
| H20.022 | Recurrent acute iridocyclitis, left eye |
| H20.023 | Recurrent acute iridocyclitis, bilateral |
| H20.029 | Recurrent acute iridocyclitis, unspecified eye |
| H20.031 | Secondary infectious iridocyclitis, right eye |
| H20.032 | Secondary infectious iridocyclitis, left eye |
| H20.033 | Secondary infectious iridocyclitis, bilateral |
| H20.039 | Secondary infectious iridocyclitis, unspecified eye |
| H20.041 | Secondary noninfectious iridocyclitis, right eye |
| H20.042 | Secondary noninfectious iridocyclitis, left eye |
| H20.043 | Secondary noninfectious iridocyclitis, bilateral |
| H20.049 | Secondary noninfectious iridocyclitis, unspecified eye |
| H20.051 | Hypopyon, right eye |
| H20.052 | Hypopyon, left eye |
| H20.053 | Hypopyon, bilateral |
| H20.059 | Hypopyon, unspecified eye |
| H20.10 | Chronic iridocyclitis, unspecified eye |
| H20.11 | Chronic iridocyclitis, right eye |
| H20.12 | Chronic iridocyclitis, left eye |
| H20.13 | Chronic iridocyclitis, bilateral |
| H20.811 | Fuchs' heterochromic cyclitis, right eye |
| H20.812 | Fuchs' heterochromic cyclitis, left eye |
| H20.813 | Fuchs' heterochromic cyclitis, bilateral |
| H20.819 | Fuchs' heterochromic cyclitis, unspecified eye |
| H20.821 | Vogt-Koyanagi syndrome, right eye |
| H20.822 | Vogt-Koyanagi syndrome, left eye |
| H20.823 | Vogt-Koyanagi syndrome, bilateral |
| H20.824 | Vogt-Koyanagi syndrome, unspecified eye |
| H20.9 | Iridocyclitis unspecified |
| K50.00 | Crohn's disease of small intestine without complications |
| K50.011 | Crohn's disease of small intestine with rectal bleeding |
| K50.012 | Crohn's disease of small intestine with intestinal obstruction |
| K50.013 | Crohn's disease of small intestine with fistula |
| K50.014 | Crohn's disease of small intestine with abscess |
| K50.018 | Crohn's disease of small intestine with other complication |
| K50.019 | Crohn's disease of small intestine with unspecified complications |
| K50.10 | Crohn's disease of large intestine without complications |
| K50.111 | Crohn's disease of large intestine with rectal bleeding |
| K50.112 | Crohn's disease of large intestine with intestinal obstruction |
| K50.113 | Crohn's disease of large intestine with fistula |
| K50.114 | Crohn's disease of large intestine with abscess |
| K50.118 | Crohn's disease of large intestine with other complication |
| K50.119 | Crohn's disease of large intestine with unspecified complications |
| K50.80 | Crohn's disease of both small and large intestine without complications |
| K50.811 | Crohn's disease of both small and large intestine with rectal bleeding |
| K50.812 | Crohn's disease of both small and large intestine with intestinal obstruction |
| K50.813 | Crohn's disease of both small and large intestine with fistula |
| K50.814 | Crohn's disease of both small and large intestine with abscess |
| K50.818 | Crohn's disease of both small and large intestine with other complication |
| K50.819 | Crohn's disease of both small and large intestine with unspecified complications |
| K50.90 | Crohn's disease, unspecified, without complications |
| K50.911 | Crohn's disease, unspecified, with rectal bleeding |
| K50.912 | Crohn's disease, unspecified, with intestinal obstruction |
| K50.913 | Crohn's disease, unspecified, with fistula |
| K50.914 | Crohn's disease, unspecified, with abscess |
| K50.918 | Crohn's disease, unspecified, with other complication |
| K50.919 | Crohn's disease, unspecified, with unspecified complications |
| K50.40 | Arthropathy in Crohn's disease [Regional enteritis] (K50.-+) multiple sites |
| K50.41 | Arthropathy in Crohn's disease [Regional enteritis] (K50.-+) shoulder region |
| K50.42 | Arthropathy in Crohn's disease [Regional enteritis] (K50.-+) upper arm |
| K50.43 | Arthropathy in Crohn's disease [Regional enteritis] (K50.-+) forearm |
| K50.44 | Arthropathy in Crohn's disease [Regional enteritis] (K50.-+) hand |
| K50.45 | Arthropathy in Crohn's disease [Regional enteritis] (K50.-+) pelvic region and thigh |
| K50.46 | Arthropathy in Crohn's disease [Regional enteritis] (K50.-+) lower leg |
| K50.47 | Arthropathy in Crohn's disease [Regional enteritis] (K50.-+) ankle and foot |
| K50.48 | Arthropathy in Crohn's disease [Regional enteritis] (K50.-+) other site |
| K50.49 | Arthropathy in Crohn's disease [Regional enteritis] (K50.-+) site unspecified |
| M09.10 | Juvenile arthritis in Crohn's disease [Regional enteritis] (K50.-+) multiple sites |
| M09.11 | Juvenile arthritis in Crohn's disease [Regional enteritis] (K50.-+) shoulder region |
| M09.12 | Juvenile arthritis in Crohn's disease [Regional enteritis] (K50.-+) upper arm |
| M09.13 | Juvenile arthritis in Crohn's disease [Regional enteritis] (K50.-+) forearm |
| M09.14 | Juvenile arthritis in Crohn's disease [Regional enteritis] (K50.-+) hand |
| M09.15 | Juvenile arthritis in Crohn's disease [Regional enteritis] (K50.-+) pelvic region and thigh |
| M09.16 | Juvenile arthritis in Crohn's disease [Regional enteritis] (K50.-+) lower leg |
| M09.17 | Juvenile arthritis in Crohn's disease [Regional enteritis] (K50.-+) ankle and foot |
| M09.18 | Juvenile arthritis in Crohn's disease [Regional enteritis] (K50.-+) other site |
| M09.19 | Juvenile arthritis in Crohn's disease [Regional enteritis] (K50.-+) site unspecified |
| K51.00 | Ulcerative (chronic) pancolitis without complications |
| K51.011 | Ulcerative (chronic) pancolitis with rectal bleeding |
| K51.012 | Ulcerative (chronic) pancolitis with intestinal obstruction |
| K51.013 | Ulcerative (chronic) pancolitis with fistula |
| K51.014 | Ulcerative (chronic) pancolitis with abscess |
| K51.018 | Ulcerative (chronic) pancolitis with other complication |
| K51.019 | Ulcerative (chronic) pancolitis with unspecified complications |
| K51.20 | Ulcerative (chronic) proctitis without complications |
| K51.211 | Ulcerative (chronic) proctitis with rectal bleeding |
| K51.212 | Ulcerative (chronic) proctitis with intestinal obstruction |
| K51.213 | Ulcerative (chronic) proctitis with fistula |
| K51.214 | Ulcerative (chronic) proctitis with abscess |
| K51.218 | Ulcerative (chronic) proctitis with other complication |
| K51.219 | Ulcerative (chronic) proctitis with unspecified complications |
| K51.30 | Ulcerative (chronic) rectosigmoiditis without complications |
| K51.311 | Ulcerative (chronic) rectosigmoiditis with rectal bleeding |
| K51.312 | Ulcerative (chronic) rectosigmoiditis with intestinal obstruction |
| K51.313 | Ulcerative (chronic) rectosigmoiditis with fistula |
| K51.314 | Ulcerative (chronic) rectosigmoiditis with abscess |
| K51.318 | Ulcerative (chronic) rectosigmoiditis with other complication |
| K51.319 | Ulcerative (chronic) rectosigmoiditis with unspecified complications |
| K51.40 | Inflammatory polyps of colon without complications |
| K51.411 | Inflammatory polyps of colon with rectal bleeding |
| K51.412 | Inflammatory polyps of colon with intestinal obstruction |
| K51.413 | Inflammatory polyps of colon with fistula |
| K51.414 | Inflammatory polyps of colon with abscess |
| K51.418 | Inflammatory polyps of colon with other complication |
| K51.419 | Inflammatory polyps of colon with unspecified complications |
| K51.50 | Left sided colitis without complications |
| K51.511 | Left sided colitis with rectal bleeding |
| K51.512 | Left sided colitis with intestinal obstruction |
| K51.513 | Left sided colitis with fistula |
| K51.514 | Left sided colitis with abscess |
| K51.518 | Left sided colitis with other complication |
| K51.519 | Left sided colitis with unspecified complications |
| K51.80 | Other ulcerative colitis without complications |
| K51.811 | Other ulcerative colitis with rectal bleeding |
| K51.812 | Other ulcerative colitis with intestinal obstruction |
| K51.813 | Other ulcerative colitis with fistula |
| K51.814 | Other ulcerative colitis with abscess |
| K51.818 | Other ulcerative colitis with other complication |
| K51.819 | Other ulcerative colitis with unspecified complications |
| K51.90 | Ulcerative colitis, unspecified without complications |
| K51.911 | Ulcerative colitis, unspecified with rectal bleeding |
| K51.912 | Ulcerative colitis, unspecified with intestinal obstruction |
| K51.913 | Ulcerative colitis, unspecified with fistula |
| K51.914 | Ulcerative colitis, unspecified with abscess |
| K51.918 | Ulcerative colitis, unspecified with other complication |
| K51.919 | Ulcerative colitis, unspecified with unspecified complications |
| M07.50 | Arthropathy in ulcerative colitis (K51.-+) multiple sites |
| M07.51 | Arthropathy in ulcerative colitis (K51.-+) shoulder region |
| M07.52 | Arthropathy in ulcerative colitis (K51.-+) upper arm |
| M07.53 | Arthropathy in ulcerative colitis (K51.-+) forearm |
| M07.54 | Arthropathy in ulcerative colitis (K51.-+) hand |
| M07.55 | Arthropathy in ulcerative colitis (K51.-+) pelvic region and thigh |
| M07.56 | Arthropathy in ulcerative colitis (K51.-+) lower leg |
| M07.57 | Arthropathy in ulcerative colitis (K51.-+) ankle and foot |
| M07.58 | Arthropathy in ulcerative colitis (K51.-+) other site |
| M07.59 | Arthropathy in ulcerative colitis (K51.-+) site unspecified |
| M09.20 | Juvenile arthritis in ulcerative colitis (K51.-+) multiple sites |
| M09.21 | Juvenile arthritis in ulcerative colitis (K51.-+) shoulder region |
| M09.22 | Juvenile arthritis in ulcerative colitis (K51.-+) upper arm |
| M09.23 | Juvenile arthritis in ulcerative colitis (K51.-+) forearm |
| M09.24 | Juvenile arthritis in ulcerative colitis (K51.-+) hand |
| M09.25 | Juvenile arthritis in ulcerative colitis (K51.-+) pelvic region and thigh |
| M09.26 | Juvenile arthritis in ulcerative colitis (K51.-+) lower leg |
| M09.27 | Juvenile arthritis in ulcerative colitis (K51.-+) ankle and foot |
| M09.28 | Juvenile arthritis in ulcerative colitis (K51.-+) other site |
| M09.29 | Juvenile arthritis in ulcerative colitis (K51.-+) site unspecified |
| K74.3 | Primary biliary cirrhosis |
| K75.4 | Autoimmune hepatitis |
| K90.0 | Celiac disease |
| L10.0 | Pemphigus vulgaris |
| L10.1 | Pemphigus vegetans |
| L10.2 | Pemphigus foliaceus |
| L10.3 | Brazilian pemphigus |
| L10.4 | Pemphigus erythematosus |
| L10.81 | Paraneoplastic pemphigus |
| L10.89 | Other pemphigus |
| L10.9 | Pemphigus, unspecified |
| L12.0 | Bullous pemphigoid |
| L12.1 | Cicatricial pemphigoid |
| L12.2 | Chronic bullous disease of childhood |
| L12.8 | Other pemphigoid |
| L12.9 | Pemphigoid, unspecified |
| H13.3 | Ocular pemphigoid |
| L40.0 | Psoriasis vulgaris |
| L40.1 | Generalised pustular psoriasis |
| L40.2 | Acrodermatitis continua |
| L40.3 | Pustulosis palmaris et plantaris |
| L40.4 | Guttate psoriasis |
| L40.50 | Arthropathic psoriasis, unspecified |
| L40.51 | Distal interphalangeal psoriatic arthropathy |
| L40.52 | Psoriatic arthritis mutilans |
| L40.53 | Psoriatic spondylitis |
| L40.54 | Psoriatic juvenile arthropathy |
| L40.59 | Other psoriatic arthropathy |
| L40.8 | Other psoriasis |
| L40.9 | Psoriasis, unspecified |
| M07.00 | Distal interphalangeal psoriatic arthropathy (L40.5+) multiple sites |
| M07.04 | Distal interphalangeal psoriatic arthropathy (L40.5+) hand |
| M07.07 | Distal interphalangeal psoriatic arthropathy (L40.5+) ankle and foot |
| M07.09 | Distal interphalangeal psoriatic arthropathy (L40.5+) site unspecified |
| M07.10 | Arthritis mutilans (L40.5+) multiple sites |
| M07.11 | Arthritis mutilans (L40.5+) shoulder region |
| M07.12 | Arthritis mutilans (L40.5+) upper arm |
| M07.13 | Arthritis mutilans (L40.5+) forearm |
| M07.14 | Arthritis mutilans (L40.5+) hand |
| M07.15 | Arthritis mutilans (L40.5+) pelvic region and thigh |
| M07.16 | Arthritis mutilans (L40.5+) lower leg |
| M07.17 | Arthritis mutilans (L40.5+) ankle and foot |
| M07.18 | Arthritis mutilans (L40.5+) other site |
| M07.19 | Arthritis mutilans (L40.5+) site unspecified |
| M07.20 | Psoriatic spondylitis (L40.5+) multiple sites |
| M07.21 | Psoriatic spondylitis (L40.5+) shoulder region |
| M07.22 | Psoriatic spondylitis (L40.5+) upper arm |
| M07.23 | Psoriatic spondylitis (L40.5+) forearm |
| M07.24 | Psoriatic spondylitis (L40.5+) hand |
| M07.25 | Psoriatic spondylitis (L40.5+) pelvic region and thigh |
| M07.26 | Psoriatic spondylitis (L40.5+) lower leg |
| M07.27 | Psoriatic spondylitis (L40.5+) ankle and foot |
| M07.28 | Psoriatic spondylitis (L40.5+) other site |
| M07.29 | Psoriatic spondylitis (L40.5+) site unspecified |
| M07.30 | Other psoriatic arthropathies (L40.5+) multiple sites |
| M07.31 | Other psoriatic arthropathies (L40.5+) shoulder region |
| M07.32 | Other psoriatic arthropathies (L40.5+) upper arm |
| M07.33 | Other psoriatic arthropathies (L40.5+) forearm |
| M07.34 | Other psoriatic arthropathies (L40.5+) hand |
| M07.35 | Other psoriatic arthropathies (L40.5+) pelvic region and thigh |
| M07.36 | Other psoriatic arthropathies (L40.5+) lower leg |
| M07.37 | Other psoriatic arthropathies (L40.5+) ankle and foot |
| M07.38 | Other psoriatic arthropathies (L40.5+) other site |
| M07.39 | Other psoriatic arthropathies (L40.5+) site unspecified |
| M09.00 | Juvenile arthritis in psoriasis (L40.5+) multiple sites |
| M09.01 | Juvenile arthritis in psoriasis (L40.5+) shoulder region |
| M09.02 | Juvenile arthritis in psoriasis (L40.5+) upper arm |
| M09.03 | Juvenile arthritis in psoriasis (L40.5+) forearm |
| M09.04 | Juvenile arthritis in psoriasis (L40.5+) hand |
| M09.05 | Juvenile arthritis in psoriasis (L40.5+) pelvic region and thigh |
| M09.06 | Juvenile arthritis in psoriasis (L40.5+) lower leg |
| M09.07 | Juvenile arthritis in psoriasis (L40.5+) ankle and foot |
| M09.08 | Juvenile arthritis in psoriasis (L40.5+) other site |
| M09.09 | Juvenile arthritis in psoriasis (L40.5+) site unspecified |
| L63.0 | Alopecia (capitis) totalis |
| L63.1 | Alopecia universalis |
| L63.2 | Ophiasis |
| L63.8 | Other alopecia areata |
| L63.9 | Alopecia areata, unspecified |
| L80 | Vitiligo |
| M05.00 | Felty's syndrome, unspecified site |
| M05.011 | Felty's syndrome, right shoulder |
| M05.012 | Felty's syndrome, left shoulder |
| M05.019 | Felty's syndrome, unspecified shoulder |
| M05.021 | Felty's syndrome, right elbow |
| M05.022 | Felty's syndrome, left elbow |
| M05.029 | Felty's syndrome, unspecified elbow |
| M05.031 | Felty's syndrome, right wrist |
| M05.032 | Felty's syndrome, left wrist |
| M05.039 | Felty's syndrome, unspecified wrist |
| M05.041 | Felty's syndrome, right hand |
| M05.042 | Felty's syndrome, left hand |
| M05.049 | Felty's syndrome, unspecified hand |
| M05.051 | Felty's syndrome, right hip |
| M05.052 | Felty's syndrome, left hip |
| M05.059 | Felty's syndrome, unspecified hip |
| M05.061 | Felty's syndrome, right knee |
| M05.062 | Felty's syndrome, left knee |
| M05.069 | Felty's syndrome, unspecified knee |
| M05.071 | Felty's syndrome, right ankle and foot |
| M05.072 | Felty's syndrome, left ankle and foot |
| M05.079 | Felty's syndrome, unspecified ankle and foot |
| M05.09 | Felty's syndrome, multiple sites |
| M05.10 | Rheumatoid lung disease with rheumatoid arthritis of unspecified site |
| M05.111 | Rheumatoid lung disease with rheumatoid arthritis of right shoulder |
| M05.112 | Rheumatoid lung disease with rheumatoid arthritis of left shoulder |
| M05.119 | Rheu lung disease with rheumatoid arthritis of unspecified shoulder |
| M05.121 | Rheumatoid lung disease with rheumatoid arthritis of right elbow |
| M05.122 | Rheumatoid lung disease with rheumatoid arthritis of left elbow |
| M05.129 | Rheumatoid lung disease with rheumatoid arthritis of unspecified elbow |
| M05.131 | Rheumatoid lung disease with rheumatoid arthritis of right wrist |
| M05.132 | Rheumatoid lung disease with rheumatoid arthritis of left wrist |
| M05.139 | Rheumatoid lung disease with rheumatoid arthritis of unspecified wrist |
| M05.141 | Rheumatoid lung disease with rheumatoid arthritis of right hand |
| M05.142 | Rheumatoid lung disease with rheumatoid arthritis of left hand |
| M05.149 | Rheumatoid lung disease with rheumatoid arthritis of unspecified hand |
| M05.151 | Rheumatoid lung disease with rheumatoid arthritis of right hip |
| M05.152 | Rheumatoid lung disease with rheumatoid arthritis of left hip |
| M05.159 | Rheumatoid lung disease with rheumatoid arthritis of unspecified hip |
| M05.161 | Rheumatoid lung disease with rheumatoid arthritis of right knee |
| M05.162 | Rheumatoid lung disease with rheumatoid arthritis of left knee |
| M05.169 | Rheumatoid lung disease with rheumatoid arthritis of unspecified knee |
| M05.171 | Rheumatoid lung disease with rheumatoid arthritis of right ankle/foot |
| M05.172 | Rheumatoid lung disease with rheumatoid arthritis of left ankle/foot |
| M05.179 | Rheumatoid lung disease with rheumatoid arthritis of unspecified ankle/foot |
| M05.19 | Rheumatoid lung disease with rheumatoid arthritis multiple sites |
| M05.20 | Rheumatoid vasculitis with rheumatoid arthritis of unspecified site |
| M05.211 | Rheumatoid vasculitis with rheumatoid arthritis of right shoulder |
| M05.212 | Rheumatoid vasculitis with rheumatoid arthritis of left shoulder |
| M05.219 | Rheumatoid vasculitis with rheumatoid arthritis of unspecified shoulder |
| M05.221 | Rheumatoid vasculitis with rheumatoid arthritis of right elbow |
| M05.222 | Rheumatoid vasculitis with rheumatoid arthritis of left elbow |
| M05.229 | Rheumatoid vasculitis with rheumatoid arthritis of unspecified elbow |
| M05.231 | Rheumatoid vasculitis with rheumatoid arthritis of right wrist |
| M05.232 | Rheumatoid vasculitis with rheumatoid arthritis of left wrist |
| M05.239 | Rheumatoid vasculitis with rheumatoid arthritis of unspecified wrist |
| M05.241 | Rheumatoid vasculitis with rheumatoid arthritis of right hand |
| M05.242 | Rheumatoid vasculitis with rheumatoid arthritis of left hand |
| M05.249 | Rheumatoid vasculitis with rheumatoid arthritis of unspecified hand |
| M05.251 | Rheumatoid vasculitis with rheumatoid arthritis of right hip |
| M05.252 | Rheumatoid vasculitis with rheumatoid arthritis of left hip |
| M05.259 | Rheumatoid vasculitis with rheumatoid arthritis of unspecified hip |
| M05.261 | Rheumatoid vasculitis with rheumatoid arthritis of right knee |
| M05.262 | Rheumatoid vasculitis with rheumatoid arthritis of left knee |
| M05.269 | Rheumatoid vasculitis with rheumatoid arthritis of unspecified knee |
| M05.271 | Rheumatoid vasculitis with rheumatoid arthritis of right ankle/foot |
| M05.272 | Rheumatoid vasculitis with rheumatoid arthritis of left ankle/foot |
| M05.279 | Rheumatoid vasculitis with rheumatoid arthritis of unspecified ankle/foot |
| M05.29 | Rheumatoid vasculitis with rheumatoid arthritis multiple sites |
| M05.30 | Rheumatoid heart disease with rheumatoid arthritis of unspecified site |
| M05.311 | Rheumatoid heart disease with rheumatoid arthritis of right shoulder |
| M05.312 | Rheumatoid heart disease with rheumatoid arthritis of left shoulder |
| M05.319 | Rheumatoid heart disease with rheumatoid arthritis of unspecified shoulder |
| M05.321 | Rheumatoid heart disease with rheumatoid arthritis of right elbow |
| M05.322 | Rheumatoid heart disease with rheumatoid arthritis of left elbow |
| M05.329 | Rheumatoid heart disease with rheumatoid arthritis of unspecified elbow |
| M05.331 | Rheumatoid heart disease with rheumatoid arthritis of right wrist |
| M05.332 | Rheumatoid heart disease with rheumatoid arthritis of left wrist |
| M05.339 | Rheumatoid heart disease with rheumatoid arthritis of unspecified wrist |
| M05.341 | Rheumatoid heart disease with rheumatoid arthritis of right hand |
| M05.342 | Rheumatoid heart disease with rheumatoid arthritis of left hand |
| M05.349 | Rheumatoid heart disease with rheumatoid arthritis of unspecified hand |
| M05.351 | Rheumatoid heart disease with rheumatoid arthritis of right hip |
| M05.352 | Rheumatoid heart disease with rheumatoid arthritis of left hip |
| M05.359 | Rheumatoid heart disease with rheumatoid arthritis of unspecified hip |
| M05.361 | Rheumatoid heart disease with rheumatoid arthritis of right knee |
| M05.362 | Rheumatoid heart disease with rheumatoid arthritis of left knee |
| M05.369 | Rheumatoid heart disease with rheumatoid arthritis of unspecified knee |
| M05.371 | Rheumatoid heart disease with rheumatoid arthritis of right ankle/foot |
| M05.372 | Rheumatoid heart disease with rheumatoid arthritis of left ankle/foot |
| M05.379 | Rheumatoid heart disease with rheumatoid arthritis of unspecified ankle/foot |
| M05.39 | Rheumatoid heart disease with rheumatoid arthritis multiple sites |
| M05.80 | Other rheumatoid arthritis with rheumatoid factor of unspecified site |
| M05.811 | Other rheumatoid arthritis with rheumatoid factor of right shoulder |
| M05.812 | Other rheumatoid arthritis with rheumatoid factor of left shoulder |
| M05.819 | Other rheumatoid arthritis with rheumatoid factor of unspecified shoulder |
| M05.821 | Other rheumatoid arthritis with rheumatoid factor of right elbow |
| M05.822 | Other rheumatoid arthritis with rheumatoid factor of left elbow |
| M05.829 | Other rheumatoid arthritis with rheumatoid factor of unspecified elbow |
| M05.831 | Other rheumatoid arthritis with rheumatoid factor of right wrist |
| M05.832 | Other rheumatoid arthritis with rheumatoid factor of left wrist |
| M05.839 | Other rheumatoid arthritis with rheumatoid factor of unspecified wrist |
| M05.841 | Other rheumatoid arthritis with rheumatoid factor of right hand |
| M05.842 | Other rheumatoid arthritis with rheumatoid factor of left hand |
| M05.849 | Other rheumatoid arthritis with rheumatoid factor of unspecified hand |
| M05.851 | Other rheumatoid arthritis with rheumatoid factor of right hip |
| M05.852 | Other rheumatoid arthritis with rheumatoid factor of left hip |
| M05.859 | Other rheumatoid arthritis with rheumatoid factor of unspecified hip |
| M05.861 | Other rheumatoid arthritis with rheumatoid factor of right knee |
| M05.862 | Other rheumatoid arthritis with rheumatoid factor of left knee |
| M05.869 | Other rheumatoid arthritis with rheumatoid factor of unspecified knee |
| M05.871 | Other rheumatoid arthritis with rheumatoid factor of right ankle/foot |
| M05.872 | Other rheumatoid arthritis with rheumatoid factor of left ankle/foot |
| M05.879 | Other rheumatoid arthritis with rheumatoid factor of unspecified ankle/foot |
| M05.89 | Other rheumatoid arthritis with rheumatoid factor multiple sites |
| M05.8A | Other rheumatoid arthritis with rheumatoid factor of other specified site |
| M05.9 | Seropositive rheumatoid arthritis, unspecified |
| M06.00 | Rheumatoid arthritis without rheumatoid factor, unspecified site |
| M06.011 | Rheumatoid arthritis with out rheumatoid factor, right shoulder |
| M06.012 | Rheumatoid arthritis with out rheumatoid factor, left shoulder |
| M06.019 | Rheumatoid arthritis with out rheumatoid factor, unspecified shoulder |
| M06.021 | Rheumatoid arthritis without rheumatoid factor, right elbow |
| M06.022 | Rheumatoid arthritis without rheumatoid factor, left elbow |
| M06.029 | Rheumatoid arthritis without rheumatoid factor, unspecified elbow |
| M06.031 | Rheumatoid arthritis without rheumatoid factor, right wrist |
| M06.032 | Rheumatoid arthritis without rheumatoid factor, left wrist |
| M06.039 | Rheumatoid arthritis without rheumatoid factor, unspecified wrist |
| M06.041 | Rheumatoid arthritis without rheumatoid factor, right hand |
| M06.042 | Rheumatoid arthritis without rheumatoid factor, left hand |
| M06.049 | Rheumatoid arthritis without rheumatoid factor, unspecified hand |
| M06.051 | Rheumatoid arthritis without rheumatoid factor, right hip |
| M06.052 | Rheumatoid arthritis without rheumatoid factor, left hip |
| M06.059 | Rheumatoid arthritis without rheumatoid factor, unspecified hip |
| M06.061 | Rheumatoid arthritis without rheumatoid factor, right knee |
| M06.062 | Rheumatoid arthritis without rheumatoid factor, left knee |
| M06.069 | Rheumatoid arthritis without rheumatoid factor, unspecified knee |
| M06.071 | Rheumatoid arthritis with out rheumatoid factor, right ankle/feet |
| M06.072 | Rheumatoid arthritis with out rheumatoid factor, left ankle/feet |
| M06.079 | Rheumatoid arthritis with out rheumatoid factor, unspecified ankle/feet |
| M06.08 | Rheumatoid arthritis without rheumatoid factor, vertebrae |
| M06.09 | Rheumatoid arthritis with out rheumatoid factor, multiple sites |
| M06.0A | Rheumatoid arthritis without rheumatoid factor, other site |
| M06.1 | Adult-onset Still disease |
| M06.20 | Rheumatoid bursitis, unspecified site |
| M06.211 | Rheumatoid bursitis, right shoulder |
| M06.212 | Rheumatoid bursitis, left shoulder |
| M06.219 | Rheumatoid bursitis, unspecified shoulder |
| M06.221 | Rheumatoid bursitis, right elbow |
| M06.222 | Rheumatoid bursitis, left elbow |
| M06.229 | Rheumatoid bursitis, unspecified elbow |
| M06.231 | Rheumatoid bursitis, right wrist |
| M06.232 | Rheumatoid bursitis, left wrist |
| M06.239 | Rheumatoid bursitis, unspecified wrist |
| M06.241 | Rheumatoid bursitis, right hand |
| M06.242 | Rheumatoid bursitis, left hand |
| M06.249 | Rheumatoid bursitis, unspecified hand |
| M06.251 | Rheumatoid bursitis, right hip |
| M06.252 | Rheumatoid bursitis, left hip |
| M06.259 | Rheumatoid bursitis, unspecified hip |
| M06.261 | Rheumatoid bursitis, right knee |
| M06.262 | Rheumatoid bursitis, left knee |
| M06.269 | Rheumatoid bursitis, unspecified knee |
| M06.271 | Rheumatoid bursitis, right ankle and foot |
| M06.272 | Rheumatoid bursitis, left ankle and foot |
| M06.279 | Rheumatoid bursitis, unspecified ankle and foot |
| M06.28 | Rheumatoid bursitis, vertebrae |
| M06.29 | Rheumatoid bursitis, multiple sites |
| M06.30 | Rheumatoid nodule, unspecified site |
| M06.311 | Rheumatoid nodule, right shoulder |
| M06.312 | Rheumatoid nodule, left shoulder |
| M06.319 | Rheumatoid nodule, unspecified shoulder |
| M06.321 | Rheumatoid nodule, right elbow |
| M06.322 | Rheumatoid nodule, left elbow |
| M06.329 | Rheumatoid nodule, unspecified elbow |
| M06.331 | Rheumatoid nodule, right wrist |
| M06.332 | Rheumatoid nodule, left wrist |
| M06.339 | Rheumatoid nodule, unspecified wrist |
| M06.341 | Rheumatoid nodule, right hand |
| M06.342 | Rheumatoid nodule, left hand |
| M06.349 | Rheumatoid nodule, unspecified hand |
| M06.351 | Rheumatoid nodule, right hip |
| M06.352 | Rheumatoid nodule, left hip |
| M06.359 | Rheumatoid nodule, unspecified hip |
| M06.361 | Rheumatoid nodule, right knee |
| M06.362 | Rheumatoid nodule, left knee |
| M06.369 | Rheumatoid nodule, unspecified knee |
| M06.371 | Rheumatoid nodule, right ankle and foot |
| M06.372 | Rheumatoid nodule, left ankle and foot |
| M06.379 | Rheumatoid nodule, unspecified ankle and foot |
| M06.38 | Rheumatoid nodule, vertebrae |
| M06.39 | Rheumatoid nodule, multiple sites |
| M06.4 | Inflammatory polyarthropathy |
| M06.80 | Other specified rheumatoid arthritis, unspecified site |
| M06.811 | Other specified rheumatoid arthritis, right shoulder |
| M06.812 | Other specified rheumatoid arthritis, left shoulder |
| M06.819 | Other specified rheumatoid arthritis, unspecified shoulder |
| M06.821 | Other specified rheumatoid arthritis, right elbow |
| M06.822 | Other specified rheumatoid arthritis, left elbow |
| M06.829 | Other specified rheumatoid arthritis, unspecified elbow |
| M06.831 | Other specified rheumatoid arthritis, right wrist |
| M06.832 | Other specified rheumatoid arthritis, left wrist |
| M06.839 | Other specified rheumatoid arthritis, unspecified wrist |
| M06.841 | Other specified rheumatoid arthritis, right hand |
| M06.842 | Other specified rheumatoid arthritis, left hand |
| M06.849 | Other specified rheumatoid arthritis, unspecified hand |
| M06.851 | Other specified rheumatoid arthritis, right hip |
| M06.852 | Other specified rheumatoid arthritis, left hip |
| M06.859 | Other specified rheumatoid arthritis, unspecified hip |
| M06.861 | Other specified rheumatoid arthritis, right knee |
| M06.862 | Other specified rheumatoid arthritis, left knee |
| M06.869 | Other specified rheumatoid arthritis, unspecified knee |
| M06.9 | Rheumatoid arthritis, unspecified |
| M08.00 | Unspecified juvenile rheumatoid arthritis of unspecified site |
| M08.011 | Unspecified juvenile rheumatoid arthritis, right shoulder |
| M08.012 | Unspecified juvenile rheumatoid arthritis, left shoulder |
| M08.019 | Unspecified juvenile rheumatoid arthritis, unspecified shoulder |
| M08.021 | Unspecified juvenile rheumatoid arthritis, right elbow |
| M08.022 | Unspecified juvenile rheumatoid arthritis, left elbow |
| M08.029 | Unspecified juvenile rheumatoid arthritis, unspecified elbow |
| M08.031 | Unspecified juvenile rheumatoid arthritis, right wrist |
| M08.032 | Unspecified juvenile rheumatoid arthritis, left wrist |
| M08.039 | Unspecified juvenile rheumatoid arthritis, unspecified wrist |
| M08.041 | Unspecified juvenile rheumatoid arthritis, right hand |
| M08.042 | Unspecified juvenile rheumatoid arthritis, left hand |
| M08.049 | Unspecified juvenile rheumatoid arthritis, unspecified hand |
| M08.051 | Unspecified juvenile rheumatoid arthritis, right hip |
| M08.052 | Unspecified juvenile rheumatoid arthritis, left hip |
| M08.059 | Unspecified juvenile rheumatoid arthritis, unspecified hip |
| M08.061 | Unspecified juvenile rheumatoid arthritis, right knee |
| M08.062 | Unspecified juvenile rheumatoid arthritis, left knee |
| M08.069 | Unspecified juvenile rheumatoid arthritis, unspecified knee |
| M08.071 | Unspecified juvenile rheumatoid arthritis, right ankle and foot |
| M08.072 | Unspecified juvenile rheumatoid arthritis, left ankle and foot |
| M08.079 | Unspecified juvenile rheumatoid arthritis, Unspecified ankle and foot |
| M08.08 | Unspecified juvenile rheumatoid arthritis, vertebrae |
| M08.09 | Unspecified juvenile rheumatoid arthritis, multiple sites |
| M08.0A | Unspecified juvenile rheumatoid arthritis, other site |
| M08.1 | Juvenile ankylosing spondylitis |
| M08.20 | Juvenile rheumatoid arthritis with systemic onset of unspecified site |
| M08.211 | Juvenile rheumatoid arthritis with systemic onset, right shoulder |
| M08.212 | Juvenile rheumatoid arthritis with systemic onset, left shoulder |
| M08.219 | Juvenile rheumatoid arthritis with systemic onset, unspecified shoulder |
| M08.221 | Juvenile rheumatoid arthritis with systemic onset, right elbow |
| M08.222 | Juvenile rheumatoid arthritis with systemic onset, left elbow |
| M08.229 | Juvenile rheumatoid arthritis with systemic onset, unspecified elbow |
| M08.231 | Juvenile rheumatoid arthritis with systemic onset, right wrist |
| M08.232 | Juvenile rheumatoid arthritis with systemic onset, left wrist |
| M08.239 | Juvenile rheumatoid arthritis with systemic onset, unspecified wrist |
| M08.241 | Juvenile rheumatoid arthritis with systemic onset, right hand |
| M08.242 | Juvenile rheumatoid arthritis with systemic onset, left hand |
| M08.249 | Juvenile rheumatoid arthritis with systemic onset, unspecified hand |
| M08.251 | Juvenile rheumatoid arthritis with systemic onset, right hip |
| M08.252 | Juvenile rheumatoid arthritis with systemic onset, left hip |
| M08.259 | Juvenile rheumatoid arthritis with systemic onset, unspecified hip |
| M08.261 | Juvenile rheumatoid arthritis with systemic onset, right knee |
| M08.262 | Juvenile rheumatoid arthritis with systemic onset, left knee |
| M08.269 | Juvenile rheumatoid arthritis with systemic onset, unspecified knee |
| M08.271 | Juvenile rheumatoid arthritis with systemic onset, right ankle and foot |
| M08.272 | Juvenile rheumatoid arthritis with systemic onset, left ankle and foot |
| M08.279 | Juvenile rheumatoid arthritis with systemic onset, Unspecified ankle and foot |
| M08.28 | Juvenile rheumatoid arthritis with systemic onset, vertebrae |
| M08.29 | Juvenile rheumatoid arthritis with systemic onset, multiple sites |
| M08.2A | Juvenile rheumatoid arthritis with systemic onset, other site |
| M08.3 | Juvenile polyarthritis (seronegative) |
| M08.40 | Pauciarticular juvenile rheumatoid arthritis of unspecified site |
| M08.411 | Pauciarticular juvenile rheumatoid arthritis, right shoulder |
| M08.412 | Pauciarticular juvenile rheumatoid arthritis, left shoulder |
| M08.419 | Pauciarticular juvenile rheumatoid arthritis, unspecified shoulder |
| M08.421 | Pauciarticular juvenile rheumatoid arthritis, right elbow |
| M08.422 | Pauciarticular juvenile rheumatoid arthritis, left elbow |
| M08.429 | Pauciarticular juvenile rheumatoid arthritis, unspecified elbow |
| M08.431 | Pauciarticular juvenile rheumatoid arthritis, right wrist |
| M08.432 | Pauciarticular juvenile rheumatoid arthritis, left wrist |
| M08.439 | Pauciarticular juvenile rheumatoid arthritis, unspecified wrist |
| M08.441 | Pauciarticular juvenile rheumatoid arthritis, right hand |
| M08.442 | Pauciarticular juvenile rheumatoid arthritis, left hand |
| M08.449 | Pauciarticular juvenile rheumatoid arthritis, unspecified hand |
| M08.451 | Pauciarticular juvenile rheumatoid arthritis, right hip |
| M08.452 | Pauciarticular juvenile rheumatoid arthritis, left hip |
| M08.459 | Pauciarticular juvenile rheumatoid arthritis, unspecified hip |
| M08.461 | Pauciarticular juvenile rheumatoid arthritis, right knee |
| M08.462 | Pauciarticular juvenile rheumatoid arthritis, left knee |
| M08.469 | Pauciarticular juvenile rheumatoid arthritis, unspecified knee |
| M08.471 | Pauciarticular juvenile rheumatoid arthritis, right ankle and foot |
| M08.472 | Pauciarticular juvenile rheumatoid arthritis, left ankle and foot |
| M08.479 | Pauciarticular juvenile rheumatoid arthritis, Unspecified ankle and foot |
| M08.48 | Pauciarticular juvenile rheumatoid arthritis, vertebrae |
| M08.4A | Pauciarticular juvenile rheumatoid arthritis, other site |
| M08.80 | Other juvenile arthritis of unspecified site |
| M08.811 | Other juvenile arthritis, right shoulder |
| M08.812 | Other juvenile arthritis, left shoulder |
| M08.819 | Other juvenile arthritis, unspecified shoulder |
| M08.821 | Other juvenile arthritis, right elbow |
| M08.822 | Other juvenile arthritis, left elbow |
| M08.829 | Other juvenile arthritis, unspecified elbow |
| M08.831 | Other juvenile arthritis, right wrist |
| M08.832 | Other juvenile arthritis, left wrist |
| M08.839 | Other juvenile arthritis, unspecified wrist |
| M08.841 | Other juvenile arthritis, right hand |
| M08.842 | Other juvenile arthritis, left hand |
| M08.849 | Other juvenile arthritis, unspecified hand |
| M08.851 | Other juvenile arthritis, right hip |
| M08.852 | Other juvenile arthritis, left hip |
| M08.859 | Other juvenile arthritis, unspecified hip |
| M08.861 | Other juvenile arthritis, right knee |
| M08.862 | Other juvenile arthritis, left knee |
| M08.869 | Other juvenile arthritis, unspecified knee |
| M08.871 | Other juvenile arthritis, right ankle and foot |
| M08.872 | Other juvenile arthritis, left ankle and foot |
| M08.879 | Other juvenile arthritis, Unspecified ankle and foot |
| M08.88 | Other juvenile arthritis, other specified site |
| M08.89 | Other juvenile arthritis, multiple sites |
| M08.90 | juvenile arthritis, unspecified of unspecified site |
| M08.911 | juvenile arthritis, unspecified, right shoulder |
| M08.912 | juvenile arthritis, unspecified, left shoulder |
| M08.919 | juvenile arthritis, unspecified, unspecified shoulder |
| M08.921 | juvenile arthritis, unspecified, right elbow |
| M08.922 | juvenile arthritis, unspecified, left elbow |
| M08.929 | juvenile arthritis, unspecified, unspecified elbow |
| M08.931 | juvenile arthritis, unspecified, right wrist |
| M08.932 | juvenile arthritis, unspecified, left wrist |
| M08.939 | juvenile arthritis, unspecified, unspecified wrist |
| M08.941 | juvenile arthritis, unspecified, right hand |
| M08.942 | juvenile arthritis, unspecified, left hand |
| M08.949 | juvenile arthritis, unspecified, unspecified hand |
| M08.951 | juvenile arthritis, unspecified, right hip |
| M08.952 | juvenile arthritis, unspecified, left hip |
| M08.959 | juvenile arthritis, unspecified, unspecified hip |
| M08.961 | juvenile arthritis, unspecified, right knee |
| M08.962 | juvenile arthritis, unspecified, left knee |
| M08.969 | juvenile arthritis, unspecified, unspecified knee |
| M08.971 | juvenile arthritis, unspecified, right ankle and foot |
| M08.972 | juvenile arthritis, unspecified, left ankle and foot |
| M08.979 | juvenile arthritis, unspecified, Unspecified ankle and foot |
| M08.98 | juvenile arthritis, unspecified, vertebrae |
| M08.99 | juvenile arthritis, unspecified, multiple sites |
| M08.9A | juvenile arthritis, unspecified, other specified site |
| M09.8 | Juvenile arthritis in other diseases classified elsewhere |
| M31.30 | Wegener's granulomatosis without renal involvement |
| M31.31 | Wegener's granulomatosis with renal involvement |
| M33.00 | Juvenile dermatomyositis, organ involvement unspecified |
| M33.01 | Juvenile dermatomyositis with respiratory involvement |
| M33.02 | Juvenile dermatomyositis with myopathy |
| M33.03 | Juvenile dermatomyositis without myopathy |
| M33.09 | Juvenile dermatomyositis with other organ involvement |
| M33.10 | Other dermatomyositis, organ involvement unspecified |
| M33.11 | Other dermatomyositis with respiratory involvement |
| M33.12 | Other dermatomyositis with myopathy |
| M33.13 | Other dermatomyositis without myopathy |
| M33.19 | Other dermatomyositis with other organ involvement |
| M33.20 | Polymyositis, organ involvement unspecified |
| M33.21 | Polymyositis with respiratory involvement |
| M33.22 | Polymyositis with myopathy |
| M33.29 | Polymyositis with other organ involvement |
| M33.90 | Dermatopolymyositis, unspecified, organ involvement unspecified |
| M33.91 | Dermatopolymyositis, unspecified with respiratory involvement |
| M33.92 | Dermatopolymyositis, unspecified with myopathy |
| M33.93 | Dermatopolymyositis, unspecified without myopathy |
| M33.99 | Dermatopolymyositis, unspecified with other organ involvement |
| M31.5 | Giant cell arteritis with polymyalgia rheumatica |
| M31.6 | Other giant cell arteritis |
| M35.3 | Polymyalgia rheumatica |
| G70.00 | Myasthenia gravis without (acute) exacerbation |
| G70.01 | Myasthenia gravis with (acute) exacerbation |
| M34.0 | progressive systemic sclerosis |
| M34.1 | CREST syndrome |
| M34.81 | Systemic sclerosis with lung involvement |
| M34.82 | Systemic sclerosis with myopathy |
| M34.83 | Systemic sclerosis with polyneuropathy |
| M34.89 | Other systemic sclerosis |
| M34.9 | Systemic sclerosis, unspecified |
| M32.10 | Systemic lupus erythematosus, organ or system involvement unspecified |
| M32.11 | Endocarditis in systemic lupus erythematosus |
| M32.12 | Pericarditis in systemic lupus erythematosus |
| M32.13 | Lung involvement in systemic lupus erythematosus |
| M32.14 | Glomerular disease in systemic lupus erythematosus |
| M32.15 | Tubulo-interstitial neuropath in systemic lupus erythematosus |
| M32.19 | Other organ or system involvement in systemic lupus erythematosus |
| M32.8 | Other forms of systemic lupus erythematosus |
| M32.9 | Systemic lupus erythematosus, unspecified |
| M35.00 | Sjögren syndrome, unspecified |
| M35.01 | Sjögren syndrome with keratoconjunctivitis |
| M35.02 | Sjögren syndrome with lung involvement |
| M35.03 | Sjögren syndrome with myopathy |
| M35.04 | Sjögren syndrome with tubulo-interstitial nephropathy |
| M35.05 | Sjögren syndrome with inflammatory arthritis |
| M35.06 | Sjögren syndrome with peripheral nervous system involvement |
| M35.07 | Sjögren syndrome with central nervous system involvement |
| M35.08 | Sjögren syndrome with gastrointestinal involvement |
| M35.09 | Sjögren syndrome with other organ involvement |
| M35.0A | Sjögren syndrome with glomerular disease |
| M35.0B | Sjögren syndrome with vasculitis |
| M35.0C | Sjögren syndrome with dental involvement |
| M45.0 | Ankylosing spondylitis of multiple sites in spine |
| M45.1 | Ankylosing spondylitis of occipito-atlanto-axial region |
| M45.2 | Ankylosing spondylitis of cervical region |
| M45.3 | Ankylosing spondylitis of cervicothoracic region |
| M45.4 | Ankylosing spondylitis of thoracic region |
| M45.5 | Ankylosing spondylitis of thoracolumbar region |
| M45.6 | Ankylosing spondylitis lumbar region |
| M45.7 | Ankylosing spondylitis of lumbosacral region |
| M45.8 | Ankylosing spondylitis sacral and sacrococcygeal region |
| M45.9 | Ankylosing spondylitis of unspecified sites in spine |
| M45.A0 | Non-radiographic axial spondyloarthritis of unspecified sites in spine |
| M45.A1 | Non-radiographic axial spondyloarthritis of occipito-atlanto-axial region |
| M45.A2 | Non-radiographic axial spondyloarthritis of cervical region |
| M45.A3 | Non-radiographic axial spondyloarthritis of cervicothoracic region |
| M45.A4 | Non-radiographic axial spondyloarthritis of thoracic region |
| M45.A5 | Non-radiographic axial spondyloarthritis of thoracolumbar region |
| M45.A6 | Non-radiographic axial spondyloarthritis of lumbar region |
| M45.A7 | Non-radiographic axial spondyloarthritis of lumbosacral region |
| M45.A8 | Non-radiographic axial spondyloarthritis of sacral and sacrococcygeal region |
| M45.AB | Non-radiographic axial spondyloarthritis of multiple sites in spine |
| M35.2 | Behcet's disease |
| D86.0 | Sarcoidosis of lung |
| D86.1 | Sarcoidosis of lymph nodes |
| D86.2 | Sarcoidosis of lung with sarcoidosis of lymph nodes |
| D86.3 | Sarcoidosis of skin |
| D86.81 | Sarcoid meningitis |
| D86.82 | Multiple cranial nerve palsies in sarcoidosis |
| D86.83 | Sarcoid iridocyclitis |
| D86.84 | Sarcoid pyelonephritis |
| D86.85 | Sarcoid myocarditis |
| D86.86 | Sarcoid arthropathy |
| D86.87 | Sarcoid myositis |
| D86.89 | Sarcoidosis of other sites |
| D86.9 | Sarcoidosis, unspecified |
| M63.3 | Myositis in sarcoidosis |
| G53.2 | Multiple cranial nerve palsies in sarcoidosis |

**Table S22.** IMID Diagnosis Codes. From Attauabi et al. [84].

| ICD-10 Code | Diagnosis Type |
| --- | --- |
| K50 | Inflammatory Bowel Disease (IBD) |
| K50.0 | Inflammatory Bowel Disease (IBD) |
| K50.1 | Inflammatory Bowel Disease (IBD) |
| K50.8 | Inflammatory Bowel Disease (IBD) |
| K50.9 | Inflammatory Bowel Disease (IBD) |
| K51 | Inflammatory Bowel Disease (IBD) |
| K51.0 | Inflammatory Bowel Disease (IBD) |
| K51.2 | Inflammatory Bowel Disease (IBD) |
| K51.3 | Inflammatory Bowel Disease (IBD) |
| K51.4 | Inflammatory Bowel Disease (IBD) |
| K51.5 | Inflammatory Bowel Disease (IBD) |
| K51.8 | Inflammatory Bowel Disease (IBD) |
| K51.9 | Inflammatory Bowel Disease (IBD) |
| M45 | Inflammatory Arthropathy |
| M46 | Inflammatory Arthropathy |
| M46.0 | Inflammatory Arthropathy |
| M46.2 | Inflammatory Arthropathy |
| M46.3 | Inflammatory Arthropathy |
| M46.4 | Inflammatory Arthropathy |
| M46.5 | Inflammatory Arthropathy |
| M46.8 | Inflammatory Arthropathy |
| M46.9 | Inflammatory Arthropathy |
| M05 | Seropositive Rheumatoid Arthritis |
| M05.0 | Seropositive Rheumatoid Arthritis |
| M05.1 | Seropositive Rheumatoid Arthritis |
| M05.2 | Seropositive Rheumatoid Arthritis |
| M05.3 | Seropositive Rheumatoid Arthritis |
| M05.8 | Seropositive Rheumatoid Arthritis |
| M05.9 | Seropositive Rheumatoid Arthritis |
| M06 | Other Rheumatoid Arthritis |
| M06.0 | Other Rheumatoid Arthritis |
| M06.1 | Other Rheumatoid Arthritis |
| M06.2 | Other Rheumatoid Arthritis |
| M06.3 | Other Rheumatoid Arthritis |
| M06.4 | Other Rheumatoid Arthritis |
| M06.8 | Other Rheumatoid Arthritis |
| M06.9 | Other Rheumatoid Arthritis |
| M07 | Psoriatic Arthritis |
| M07.0 | Psoriatic Arthritis |
| M07.1 | Psoriatic Arthritis |
| M07.2 | Psoriatic Arthritis |
| M07.3 | Psoriatic Arthritis |
| M07.4 | Psoriatic Arthritis |
| M07.5 | Psoriatic Arthritis |
| M07.6 | Psoriatic Arthritis |
| L40 | Psoriasis |
| L40.0 | Psoriasis |
| L40.1 | Psoriasis |
| L40.2 | Psoriasis |
| L40.3 | Psoriasis |
| L40.4 | Psoriasis |
| L40.5 | Psoriasis |
| L40.8 | Psoriasis |
| L40.9 | Psoriasis |

## List of members of the Danish COVID-19 Genome Consortium (DCGC)

Below is a list of members of The Danish COVID-19 Genome Consortium. Individuals may appear both as authors and as acknowledged DCGC members.

Anders Fomsgaard^1^, Morten Rasmussen^1^, Søren M. Karst^1^, Jannik Fonager^1^, Vithiagaran Gunalan^1^, Marc Bennedbæk^1^, Aleksander Ring^1^, Marc Stegger^1^, Raphael Sieber^1^, Kirsten Ellegaard^1^, Anna C. Ingham^1^, Thor B. Johannesen^1^, Gitte Nygaard Aasbjerg^1^, Berit Lilje^1^, Kim L. Ng^1^, Sofie M. Edslev^1^, Sharmin Baig^1^, Povilas Matusevicius^1^, Lars Bustamante Christoffersen^1^, Man-Hung Eric Tang^1^, Leandro Andrés Escobar-Herrera^1^, Casper Westergaard^1^, Christina Wiid Svarrer^1^, Nour Saad Al-Tamimi^1^, Marie Bækvad-Hansen^1^, Jonas Byberg-Grauholm^1^, Mette Theilgaard Christiansen^1^, Karen Marie Jørgensen^2^, Tobias Nikolaj Gress Hansen^2^, Esben Mørk Hartmann^2^, Nicolai Balle Larsen^2^, Arieh Cohen^2^, Kasper Holtz Uhre^3^, Christian Nielsen^3^, Anders Jensen^3^, Jakob Matias Melbye^3^, Bartlomiej Wilkowski^3^, Karina Meden Sørensen^3^, Mads Albertsen^4^, Thomas Y. Michaelsen^4^, Vang Le-Quy^4^, Mantas Sereika^4^, Rasmus H. Kirkegaard^4^, Kasper S. Andersen^4^, Martin H. Andersen^4^, Karsten K. Hansen^4^, Mads Boye^4^, Mads P. Bach^4^, Peter Dissing^4^, Anton Drastrup-Fjordbak^4^, Michael Collin^4^, Finn Büttner^4^, Jakob Brandt^4^, Simon Knuttson^4^, Emil A. Sørensen^4^, Thomas B. N. Jensen^4^, Trine Sørensen^4^, Celine Petersen^4^, Clarisse Chiche-Lapierre^4^, Frederik T. Hansen^4^, Emilio F. Collados^4^, Amalie Berg^4^, Susanne R. Bielidt^4^, Sebastian M. Dall^4^, Erika Dvarionaite^4^, Susan H. Hansen^4^, Vibeke R. Jørgensen^4^, Trine B. Nicolajsen^4^, Wagma Saei^4^, Stine K. Østergaard^4^, Martin Bøgsted^4^, Rasmus Brøndu^4^, Katja Hose^4^, Tomer Sagi^4^, Miroslav Pakanec^4^, Henrik Krarup^5^, David Fuglsang-Damgaard^5^, Mette Mølvadgaard^5^, Marc T. K. Nielsen^5^, Kristian Schønning^6^, Martin S. Pedersen^6^, Rasmus L. Marvig^6^, Nikolai Kirkby^6^, Uffe V. Schneider^7^, Jose A. S. Castruita^7^, Nana G. Jacobsen^7^, Christian Ø. Andersen^7^, Mette Christiansen^8^, Ole H. Larsen^8^, Kristian A. Skipper^8^, Søren Vang^8^, Kurt J. Handberg^8^, Carl M. Kobel^8^, Camilla Andersen^8^, Irene H. Tarpgaard^8^, Svend Ellermann-Eriksen^8^, Marianne Skov^9^, Thomas V. Sydenham^9^, Lene Nielsen^10^, Line L. Nilsson^10^, Martin B. Friis^10^,Thomas Sundelin^10^, Thomas A. Hansen^10^, Dorte T. Andersen^11^, John E. Coia^11^, Anders Jensen^12^, Ea S. Marmolin^12^, Xiaohui C. Nielsen^13^, Christian H. Schouw^13^, Simon Knutsson^4^, Marianne Kragh Thomsen^8^, Tine Sneibjerg Ebsen^8^, Morten Hoppe^10^, Asta Laugesen^13^, Laus Vejrum^13^, Rikke Johansen^13^, Josefine Møller^13^, Tina Madsen^13^

^1^Statens Serum Institut, Copenhagen, Denmark

^2^TestCenter Denmark, Copenhagen, Denmark

^3^Danish National Biobank, Copenhagen, Denmark

^4^Aalborg University, Aalborg, Denmark

^5^Aalborg University Hospital, Aalborg, Denmark

^6^Rigshospitalet, Copenhagen University Hospital, Copenhagen, Denmark

^7^Hvidovre Hospital, Hvidovre, Denmark

^8^Aarhus University Hospital, Aarhus, Denmark

^9^Odense University Hospital, Odense, Denmark

^10^Herlev Hospital, Herlev, Denmark

^11^Sydvestjysk Sygehus, Denmark

^12^Sygehus Lillebaelt, Denmark

^13^Zealand University Hospital, Denmark

## Notes

### Competing Interest Statement

The authors have declared no competing interest.

### Author Declarations

This study was conducted using data from national registers only. According to Danish law, ethics approval is not needed for this type of research. All data management and analyses were carried out on the secure research servers of Statistics Denmark. The study only contains aggregated results and no personal data.

## References

1. Riddell, A. C. & Cutino-Moguel, T. The origins of new SARS-COV-2 variants in immuncompromised individuals. Current Opinion in HIV and AIDS. ISSN: 1746-630X, 1746-6318. https://journals.lww.com/10.1097/COH.0000000000000794 (2023).

2. Wilkinson, S. A. J., et al. Recurrent SARS-CoV-2 mutations in immunodeficient patients. Virus Evolution 8, veac050. ISSN: 2057-1577. https://academic.oup.com/ve/article/doi/10.1093/ve/veac050/6661028 (2022).

3. Choi, B. et al. Persistence and Evolution of SARS-CoV-2 in an Immunocompromised Host. New England Journal of Medicine 383, 2291–2293. ISSN: 0028-4793, 1533-4406. https://www.nejm.org/doi/10.1056/NEJMc2031364 (2020).

4. Sepulcri, C. et al. The Longest Persistence of Viable SARS-CoV-2 With Recurrence of Viremia and Relapsing Symptomatic COVID-19 in an Immunocompromised Patient—A Case Study. Open Forum Infectious Diseases 8, ofab217. ISSN: 2328-8957. https://academic.oup.com/ofid/article/doi/10.1093/ofid/ofab217/6257145 (2021).

5. Birnie, E. et al. Development of Resistance-Associated Mutations After Sotrovimab Administration in High-risk Individuals Infected With the SARS-CoV-2 Omicron Variant. JAMA 328, 1104. ISSN: 0098-7484. https://jamanetwork.com/journals/jama/fullarticle/2794999 (2022).

6. Harari, S. et al. Drivers of adaptive evolution during chronic SARS-CoV-2 infections. Nature Medicine 28, 1501–1508 (2022).

7. Ghafari, M. et al. Prevalence of persistent SARS-CoV-2 in a large community surveillance study. Nature 626, 1094–1101. ISSN: 0028-0836, 1476-4687. https://www.nature.com/articles/s41586-024-07029-4 (2024).

8. Bendall, E. E. et al. Rapid transmission and tight bottlenecks constrain the evolution of highly transmissible SARS-CoV-2 variants. en. Nature Communications 14, 272. ISSN: 2041-1723. https://www.nature.com/articles/s41467-023-36001-5 (2023).

9. Carabelli, A. M. et al. SARS-CoV-2 variant biology: immune escape, transmission and fitness. Nature Reviews Microbiology (2023).

10. Schoefbaenker, M. et al. Characterisation of the antibody-mediated selective pressure driving intra-host evolution of SARS-CoV-2 in prolonged infection. PLOS Pathogens 20 (ed Gorbalenya, A. E.) e1012624. ISSN: 1553-7374. https://dx.plos.org/10.1371/journal.ppat.1012624 (2024).

11. Ip, J. D. et al. The significance of recurrent de novo amino acid substitutions that emerged during chronic SARS-CoV-2 infection: an observational study. eBioMedicine 107, 105273. https://linkinghub.elsevier.com/retrieve/pii/S2352396424003098 (2024).

12. Cao, Y. et al. Imprinted SARS-CoV-2 humoral immunity induces convergent Omicron RBD evolution. Nature. ISSN: 0028-0836, 1476-4687. https://www.nature.com/articles/s41586-022-05644-7 (2022).

13. Groenheit, R., et al. Rapid emergence of omicron sublineages expressing spike protein R346T. The Lancet Regional Health - Europe 24, 100564. https://linkinghub.elsevier.com/retrieve/pii/S2666776222002605 (2023).

14. Dong, J. et al. Genetic and structural basis for SARS-CoV-2 variant neutralization by a two-antibody cocktail. Nature Microbiology 6, 1233–1244. ISSN: 2058-5276. https://www.nature.com/articles/s41564-021-00972-2 (2021).

15. Rockett, R. et al. Resistance Mutations in SARS-CoV-2 Delta Variant after Sotrovimab Use. New England Journal of Medicine 386, 1477–1479. ISSN: 0028-4793, 1533-4406. http://www.nejm.org/doi/10.1056/NEJMc2120219 (2022).

16. Wang, Q. et al. Resistance of SARS-CoV-2 omicron subvariant BA.4.6 to antibody neutralisation. The Lancet Infectious Diseases 22, 1666–1668. https://linkinghub.elsevier.com/retrieve/pii/S1473309922006946 (2022).

17. Liu, Z. et al. Identification of SARS-CoV-2 spike mutations that attenuate monoclonal and serum antibody neutralization. Cell Host & Microbe 29, 477–488.e4. https://linkinghub.elsevier.com/retrieve/pii/S1931312821000445 (2021).

18. Ghafari, M. et al. Determinants of SARS-CoV-2 within-host evolutionary rates in persistently infected individuals 2024. http://medrxiv.org/lookup/doi/10.1101/2024.06.21.24309297.

19. Karthikeyan, S. et al. Wastewater sequencing reveals early cryptic SARS-CoV-2 variant transmission. Nature 609, 101–108. ISSN: 0028-0836, 1476-4687. https://www.nature.com/articles/s41586-022-05049-6 (2022).

20. Smyth, D. S., et al. Tracking cryptic SARS-CoV-2 lineages detected in NYC wastewater. Nature Communications 13, 635. ISSN: 2041-1723. https://www.nature.com/articles/s41467-022-28246-3 (2022).

21. Shafer, M. M. et al. Tracing the origin of SARS-CoV-2 omicron-like spike sequences detected in an urban sewershed: a targeted, longitudinal surveillance study of a cryptic wastewater lineage. The Lancet Microbe 5, e335–e344. https://linkinghub.elsevier.com/retrieve/pii/S2666524723003725 (2024).

22. Dennehy, J. J., Gupta, R. K., Hanage, W. P., Johnson, M. C. & Peacock, T. P. Where is the next SARS-CoV-2 variant of concern? The Lancet 399, 1938–1939. https://linkinghub.elsevier.com/retrieve/pii/S0140673622007437 (2022).

23. Otto, S. P. et al. The origins and potential future of SARS-CoV-2 variants of concern in the evolving COVID-19 pandemic. Current Biology 31, R918–R929. https://linkinghub.elsevier.com/retrieve/pii/S0960982221008782 (2021).

24. Ghafari, M., Liu, Q., Dhillon, A., Katzourakis, A. & Weissman, D. B. Investigating the evolutionary origins of the first three SARS-CoV-2 variants of concern. Frontiers in Virology 2, 942555. ISSN: 2673-818X. https://www.frontiersin.org/articles/10.3389/fviro.2022.942555/full (2022).

25. Hill, V., et al. The origins and molecular evolution of SARS-CoV-2 lineage B.1.1.7 in the UK. Virus Evolution 8, veac080. ISSN: 2057-1577. https://academic.oup.com/ve/article/doi/10.1093/ve/veac080/6677185 (2022).

26. Deng, X. et al. Genomic surveillance reveals multiple introductions of SARS-CoV-2 into Northern California. Science 369, 582–587 (2020).

27. Robishaw, J. D. et al. Genomic surveillance to combat COVID-19: challenges and opportunities. The Lancet Microbe 2, e481–e484 (2021).

28. Khurana, M. P., et al. High-resolution epidemiological landscape from ∼290,000 SARS-CoV-2 genomes from Denmark. Nature Communications 15, 7123. ISSN: 2041-1723. https://www.nature.com/articles/s41467-024-51371-0 (2024).

29. Stan Development Team. Stan modeling language users guide and reference manual, version 2.18.0 2018. http://mc-stan.org/.

30. Charlson, M. E., Pompei, P., Ales, K. L. & MacKenzie, C. A new method of classifying prognostic comorbidity in longitudinal studies: Development and validation. Journal of Chronic Diseases 40, 373–383. https://linkinghub.elsevier.com/retrieve/pii/0021968187901718 (1987).

31. Bloom, J. D. & Neher, R. A. Fitness effects of mutations to SARS-CoV-2 proteins. en. Virus Evolution 9, vead055. ISSN: 2057-1577. https://academic.oup.com/ve/article/doi/10.1093/ve/vead055/7265011 (2024).

32. Challen, R., Brooks-Pollock, E., Tsaneva-Atanasova, K. & Danon, L. Meta-analysis of the severe acute respiratory syndrome coronavirus 2 serial intervals and the impact of parameter uncertainty on the coronavirus disease 2019 reproduction number. en. Statistical Methods in Medical Research 31, 1686–1703. ISSN: 0962-2802, 1477-0334. https://journals.sagepub.com/doi/10.1177/09622802211065159 (2024) (Sept. 2022).

33. Bürkner, P.-C. brms: An R package for Bayesian multilevel models using Stan. Journal of Statistical Software 80, 1–28 (2017).

34. Tanino, Y. et al. Emergence of SARS-CoV-2 with Dual-Drug Resistant Mutations During a Long-Term Infection in a Kidney Transplant Recipient. Infection and Drug Resistance Volume 17, 531–541. ISSN: 1178-6973. https://www.dovepress.com/emergence-of-sars-cov-2-with-dual-drug-resistant-mutations-during-a-lo-peer-reviewed-fulltext-article-IDR (2024).

35. Wang, Q. et al. Key mutations in the spike protein of SARS-CoV-2 affecting neutralization resistance and viral internalization. Journal of Medical Virology 95, e28407. ISSN: 0146-6615, 1096-9071. https://onlinelibrary.wiley.com/doi/10.1002/jmv.28407 (2023).

36. Kaku, Y., et al. Virological characteristics of the SARS-CoV-2 KP.2 variant. The Lancet Infectious Diseases 24, e416. https://linkinghub.elsevier.com/retrieve/pii/S1473309924002986 (2024).

37. Carlin, A. F. et al. Virologic and Immunologic Characterization of Coronavirus Disease 2019 Recrudescence After Nirmatrelvir/Ritonavir Treatment. en. Clinical Infectious Diseases 76, e530–e532. ISSN: 1058-4838, 1537-6591. https://academic.oup.com/cid/article/76/3/e530/6611663 (2023).

38. Stevens, L. J. et al. Mutations in the SARS-CoV-2 RNA-dependent RNA polymerase confer resistance to remdesivir by distinct mechanisms. en. Science Translational Medicine 14, eabo0718. https://www.science.org/doi/10.1126/scitranslmed.abo0718 (Aug. 2022).

39. Igari, H., et al. Dynamic diversity of SARS-CoV-2 genetic mutations in a lung transplantation patient with persistent COVID-19. en. Nature Communications 15, 3604. ISSN: 2041-1723. https://www.nature.com/articles/s41467-024-47941-x (2024).

40. Chang, E. J. et al. 464. Whole Genome Sequencing and Mutation Analysis of SARS-CoV-2 in the Immunocompromised Patients with Persistent SARS-CoV-2 Shedding. en. Open Forum Infectious Diseases 10, ofad500.534. ISSN: 2328-8957. https://academic.oup.com/ofid/article/doi/10.1093/ofid/ofad500.534/7448443 (2023).

41. Sanderson, T. et al. A molnupiravir-associated mutational signature in global SARS-CoV-2 genomes. Nature 623, 594–600. ISSN: 0028-0836, 1476-4687. https://www.nature.com/articles/s41586-023-06649-6 (2023).

42. Picard, G. et al. Emergence, spread and characterisation of the SARS-CoV-2 variant B.1.640 circulating in France, October 2021 to February 2022. Eurosurveillance 28. ISSN: 1560-7917. https://www.eurosurveillance.org/content/10.2807/1560-7917.ES.2023.28.22.2200671 (2023).

43. Saifi, S., et al. SARS-CoV-2 VOCs, Mutational diversity and clinical outcome: Are they modulating drug efficacy by altered binding strength? Genomics 114, 110466. https://linkinghub.elsevier.com/retrieve/pii/S0888754322002117 (2022).

44. Focosi, D., et al. Monoclonal antibody therapies against SARS-CoV-2. The Lancet Infectious Diseases 22, e311–e326. https://linkinghub.elsevier.com/retrieve/pii/S1473309922003115 (2022).

45. Murakami, N. et al. Therapeutic advances in COVID-19. Nature Reviews Nephrology 19, 38–52. ISSN: 1759-5061, 1759-507X. https://www.nature.com/articles/s41581-022-00642-4 (2023).

46. Marques, B. D. C., et al. Impact of Vaccination on Intra-Host Genetic Diversity of Patients Infected with SARS-CoV-2 Gamma Lineage. en. Viruses 16, 1524. ISSN: 1999-4915. https://www.mdpi.com/1999-4915/16/10/1524 (2024).

47. Zhang, Y. et al. Vaccination Shapes Within-Host SARS-CoV-2 Diversity of Omicron BA.2.2 Breakthrough Infection. en. The Journal of Infectious Diseases 229, 1711–1721. ISSN: 0022-1899, 1537-6613. https://academic.oup.com/jid/article/229/6/1711/7502446 (2024).

48. Gu, H., et al. Within-host genetic diversity of SARS-CoV-2 lineages in unvaccinated and vaccinated individuals. en. Nature Communications 14, 1793. ISSN: 2041-1723. https://www.nature.com/articles/s41467-023-37468-y (2023).

49. Maponga, T. G., et al. Persistent SARS-CoV-2 Infection with Accumulation of Mutations in a Patient with Poorly Controlled HIV Infection. en. SSRN Electronic Journal. ISSN: 1556-5068. https://www.ssrn.com/abstract=4014499 (2022).

50. Karim, F. et al. Clearance of persistent SARS-CoV-2 associates with increased neutralizing antibodies in advanced HIV disease post-ART initiation. en. Nature Communications 15, 2360. ISSN: 2041-1723. https://www.nature.com/articles/s41467-024-46673-2 (2024).

51. Corey, L. et al. SARS-CoV-2 Variants in Patients with Immunosuppression. en. New England Journal of Medicine 385, 562–566. ISSN: 0028-4793, 1533-4406. http://www.nejm.org/doi/10.1056/NEJMsb2104756 (2021).

52. Cele, S. et al. SARS-CoV-2 prolonged infection during advanced HIV disease evolves extensive immune escape. en. Cell Host & Microbe 30, 154–162.e5. ISSN: 19313128. https://linkinghub.elsevier.com/retrieve/pii/S1931312822000415 (2022).

53. Lustig, G. et al. SARS-CoV-2 infection in immunosuppression evolves sub-lineages which independently accumulate neutralization escape mutations. en. Virus Evolution 10, vead075. ISSN: 2057-1577. https://academic.oup.com/ve/article/doi/10.1093/ve/vead075/7503539 (2024).

54. Baang, J. H. et al. Prolonged Severe Acute Respiratory Syndrome Coronavirus 2 Replication in an Immunocompromised Patient. en. The Journal of Infectious Diseases 223, 23–27. ISSN: 0022-1899, 1537-6613. https://academic.oup.com/jid/article/223/1/23/5934826 (Jan. 2021).

55. Tran-Kiem, C. & Bedford, T. Estimating the reproduction number and transmission heterogeneity from the size distribution of clusters of identical pathogen sequences. en. Proceedings of the National Academy of Sciences 121, e2305299121. ISSN: 0027-8424, 1091-6490. https://pnas.org/doi/10.1073/pnas.2305299121 (2024) (Apr. 2024).

56. Tran-Kiem, C. et al. Fine-scale spatial and social patterns of SARS-CoV-2 transmission from identical pathogen sequences en. May 2024. http://medrxiv.org/lookup/doi/10.1101/2024.05.24.24307811 (2024).

57. Quick, J. nCoV-2019 sequencing protocol v2 (GunIt) v2 2020. https://www.protocols.io/view/ncov-2019-sequencing-protocol-v2-bdp7i5rn.

58. Sorensen, E. A., Karst, S. M. & Knutsson, S. AAU-nCoV-2019_Tailed_Long_Amplicon_Sequncing V.2. https://www.protocols.io/view/aau-ncov-2019-tailed-long-amplicon-sequncing-bfc3jiyn (2020).

59. Martin, M. Cutadapt removes adapter sequences from high-throughput sequencing reads. EMBnet.journal 17, 10 (2011).

60. nanoporetech/medaka original-date: 2017-06-07. 2024. https://github.com/nanoporetech/medaka.

61. Quick, J. nCoV-2019 sequencing protocol v3 (LoCost) v3 2020. https://www.protocols.io/view/ncov-2019-sequencing-protocol-v3-locost-bh42j8ye.

62. Tang, M.-H. E. et al. Comparative subgenomic mRNA profiles of SARS-CoV-2 Alpha, Delta and Omicron BA.1, BA.2 and BA.5 sub-lineages using Danish COVID-19 genomic surveillance data. eBioMedicine 93, 104669 (2023).

63. Grubaugh, N. D. et al. An amplicon-based sequencing framework for accurately measuring intrahost virus diversity using PrimalSeq and iVar. Genome Biology 20, 8 (2019).

64. Nawrocki, E. P. Faster SARS-CoV-2 sequence validation and annotation for GenBank using VADR. NAR Genomics and Bioinformatics 5, lqad002 (2023).

65. Zhou, P. et al. A pneumonia outbreak associated with a new coronavirus of probable bat origin. Nature 579, 270–273 (2020).

66. Katoh, K., Rozewicki, J. & Yamada, K. D. MAFFT online service: multiple sequence alignment, interactive sequence choice and visualization. Briefings in bioinformatics 20, 1160–1166 (2019).

67. De Maio, N. et al. Maximum likelihood pandemic-scale phylogenetics. Nature Genetics, 1–7 (2023).

68. Hadfield, J. et al. Nextstrain: real-time tracking of pathogen evolution. Bioinformatics 34, 4121–4123 (2018).

69. Cevik, M. et al. SARS-CoV-2, SARS-CoV, and MERS-CoV viral load dynamics, duration of viral shedding, and infectiousness: a systematic review and meta-analysis. en. The Lancet Microbe 2, e13–e22. ISSN: 26665247. https://linkinghub.elsevier.com/retrieve/pii/S2666524720301725 (2021).

70. Jones, T. C., et al. Estimating infectiousness throughout SARS-CoV-2 infection course. en. Science 373, eabi5273. ISSN: 0036-8075, 1095-9203. https://www.science.org/doi/10.1126/science.abi5273 (2021).

71. Lyngse, F. P. et al. Effect of vaccination on household transmission of SARS-CoV-2 Delta variant of concern. Nature Communications 13, 3764 (2022).

72. Lynge, E., Sandegaard, J. L. & Rebolj, M. The Danish National Patient Register. Scandinavian Journal of Public Health 39, 30–33. ISSN: 1403-4948, 1651-1905. https://journals.sagepub.com/doi/10.1177/1403494811401482 (2011).

73. Rytgaard HC, Anders M, Torp-Pedersen C & Gerds T. R package “heaven” 2024. https://github.com/tagteam/heaven.

74. Quan, H. et al. Updating and Validating the Charlson Comorbidity Index and Score for Risk Adjustment in Hospital Discharge Abstracts Using Data From 6 Countries. American Journal of Epidemiology 173, 676–682. ISSN: 0002-9262, 1476-6256. https://academic.oup.com/aje/article-lookup/doi/10.1093/aje/kwq433 (2011).

75. Thygesen, S. K., Christiansen, C. F., Christensen, S., Lash, T. L. & Sørensen, H. T. The predictive value of ICD-10 diagnostic coding used to assess Charlson comorbidity index conditions in the population-based Danish National Registry of Patients. BMC Medical Research Methodology 11, 83. ISSN: 1471-2288. https://bmcmedresmethodol.biomedcentral.com/articles/10.1186/1471-2288-11-83 (2011).

76. Andrews, N. et al. Duration of Protection against Mild and Severe Disease by Covid-19 Vaccines. en. New England Journal of Medicine 386, 340–350. ISSN: 0028-4793, 1533-4406. http://www.nejm.org/doi/10.1056/NEJMoa2115481 (Jan. 2022).

77. Dietler, D., Kahn, F., Inghammar, M. & Björk, J. Waning protection after vaccination and prior infection against COVID-19-related mortality over 18 months. en. Clinical Microbiology and Infection 29, 1573–1580. ISSN: 1198743X. https://linkinghub.elsevier.com/retrieve/pii/S1198743X23003889 (Dec. 2023).

78. Srivastava, K. et al. SARS-CoV-2-infection- and vaccine-induced antibody responses are long lasting with an initial waning phase followed by a stabilization phase. en. Immunity 57, 587–599.e4. ISSN: 10747613. https://linkinghub.elsevier.com/retrieve/pii/S1074761324000438 (2024) (Mar. 2024).

79. Shapiro, S. S. & Wilk, M. B. An Analysis of Variance Test for Normality (Complete Samples). Biometrika 52, 591. https://www.jstor.org/stable/2333709?origin=crossref (1965).

80. Venables, W. N. & Ripley, B. D. Modern Applied Statistics with S 4th. ISBN: 0-387-95457-0. https://www.stats.ox.ac.uk/pub/MASS4/ (Springer, New York, 2002).

81. Mansfield, K. E., et al. Clinical Codelist - Immunosuppression - ICD-10 Codes 2019. http://datacompass.lshtm.ac.uk/id/eprint/1276.

82. Mansfield, K. E., et al. Clinical Codelist - Diabetes Mellitus Diagnostic - ICD-10 Codes 2019. http://datacompass.lshtm.ac.uk/id/eprint/1273.

83. Tunnicliffe, L. & Warren-Gash, C. Clinical codelist-autoimmune disease ICD10 codes 2022. https://datacompass.lshtm.ac.uk/id/eprint/2853.

84. Attauabi, M. et al. Coronavirus disease 2019, immune-mediated inflammatory diseases and immunosuppressive therapies – A Danish population-based cohort study. Journal of Autoimmunity 118, 102613. https://linkinghub.elsevier.com/retrieve/pii/S0896841121000214 (2021).

